# Evidence synthesis on sex differences in mortality after traumatic brain injury: Systematic review integrating meta-analysis and explainable machine learning

**DOI:** 10.64898/2026.10.02.26364454

**Authors:** Daniel Phan, Yupeng Yao, Urooba Shaikh, Nada Ahmad, Tatyana Mollayeva

## Abstract

Sex differences in mortality after traumatic brain injury (TBI) have been inconsistently reported, even after accounting for age, injury severity, and other injury related factors.

Understanding the sources of this heterogeneity is important for prognostic research and the development of data-driven clinical decision-support tools. We conducted a systematic review and meta-analysis of sex-specific mortality after adult TBI and used explainable machine learning (ML) to investigate potential modifiers of between-study effect estimates. We searched eight databases from inception to August 25, 2025, for studies reporting mortality estimates adjusted for at least age and injury severity. Risk of bias was assessed using the Quality in Prognosis Studies tool. Male-to-female mortality estimates were pooled using random-effects meta-analysis. Three explainable ML algorithms were used to identify important modifiers of study-level effect estimates. Among 9,418 records identified, 21 studies comprising 899,813 participants and 98,286 deaths met the inclusion criteria. Pooled analyses showed lower mortality among females than males (OR 0.88, 95% CI 0.80-0.98; HR 0.76, 95% CI 0.59-0.98). Lower mortality among females was also observed for in-hospital mortality (OR 0.86, 95% CI 0.76-0.97) and overall or unspecified mortality (OR 0.83, 95% CI 0.78-0.88). Substantial residual heterogeneity remained across analyses. Explainable ML identified mean age, study sex composition, and study quality as important modifiers of reported effect estimates. These findings were consistent across sensitivity analyses. Female sex was associated with lower mortality after adult TBI, although substantial between-study heterogeneity remained.

Explainable ML identified demographic and methodological factors that may contribute to variation in reported associations. These findings highlight the importance of population characteristics and study context when developing, validating, and interpreting explainable digital prognostic tools for TBI. PROSPERO: CRD420251252136.

**Author Summary:** Existing evidence syntheses on sex differences in mortality after traumatic brain injury (TBI) remains inconclusive, with some systematic reviews reporting female sex being protective to mortality, while others showing no differences. This raised a question on whether social or study methodology factors may be responsible for this heterogeneity.

To answer the question, we conducted a systematic review and integrated meta-analysis with explainable machine learning (ML). We extracted and organized data concerning social and cohort-related factors using the PROGRESS-Plus framework from 21 studies. We conducted a meta-analysis of the pooled effects of sex differences on mortality outcomes and explored between-study heterogeneity, using three ML algorithms to identify important modifiers.

Our meta-analysis confirmed substantial heterogeneity in the association between sex and mortality, with important variation in both the magnitude and direction of reported effects.

Explainable ML identified study-level age, sex composition, and methodological quality as the major drivers of heterogeneity, with consistent findings across several sensitivity analyses.

Our findings have implications for future digital health research. Explainable ML may be a useful tool for interpreting variability in existing evidence and informing the design and interpretation of future studies examining sex differences in TBI outcomes.

## Introduction

Traumatic brain injury (TBI), an injury to the brain caused by an external force to the head or body [1], is a major cause of death and disability, with an estimated 69 million cases and 4.48 million deaths occurring annually worldwide [2, 3]. Despite recent advances in acute trauma care, mortality remains substantial, particularly among people with moderate to severe TBI. Improving prognostic assessment therefore requires a clearer understanding of factors of the factors that contribute to variability in survival after TBI.

Mortality estimates following TBI vary widely across populations and are linked to demographic, clinical, and injury-related factors. Greater injury severity and polytrauma are consistently associated with substantially higher mortality risk compared with mild TBI (mTBI), and isolated injuries [2, 4, 5]. Age is another well-established prognostic factor, with older adults representing a particularly vulnerable population because of frailty, reduced physiological reserve, and a greater burden of comorbidities, that complicate acute management and recovery [6, 7]. The role sex as a prognostic factor in mortality following TBI remains uncertain [8, 9]. A recent systematic review and meta-analysis by Beijer et al. [10] found no statistically significant associations between sex and mortality on crude estimates, and meta-regression did not identify sex-specific differences in age as a source of variation. In contrast, findings from Ng et al. [11] reported less favorable outcomes among females, in both crude analyses and analyses accounting for age and injury severity. These conflicting findings suggest that differences in study populations, analytical approaches, covariate adjustment and methodological quality may contribute to the inconsistent findings across studies in sex-specific mortality estimates.

Sex differences in mortality between may also reflect complex interactions among biological, clinical, social, and environmental factors [5, 6, 12, 13, 14, 15, 16]. However, the extent to which these factors together contribute to variation in sex-specific mortality after TBI evidence remains poorly understood. Conventional meta-analytic approaches can quantify average effects and examine prespecified moderators but may be less suited to detecting complex or nonlinear relationships among multiple study-level characteristics. Exploratory machine learning (ML) may complement meta-analysis by identifying features that contribute most strongly to variation in effect estimates and thereby provide additional insight into sources of between-study heterogeneity [17]. Such approaches may be particularly relevant to digital health research, where understanding population heterogeneity and model interpretability is important for developing and validating generalizable prognostic tools.

We therefore conducted comprehensive meta-research consisted of systematic review and meta-analysis with the aim to quantify the association between sex and mortality among adults with TBI, with specific account for age and injury severity. We further applied an explainable ML framework to explore study-level characteristics associated with variation in sex-specific mortality estimates. By integrating evidence synthesis with explainable ML, we aimed to characterize the consistency of sex-specific mortality associations and identify factors that may contribute to heterogeneity across the existing evidence, with implications for the development and evaluation of data-driven prognostic approaches in TBI.

## Results

### Study Selection

Out of 9,418 records identified in database searches, 6,365 records remained after removing duplicates, and 170 studies were eligible for full-text review based on general parameters. Of these, 21 studies [11, 18, 19, 20, 21, 22, 23, 24, 25, 26, 27, 28, 29, 30, 31, 32, 33, 34, 35, 36, 37] published between 2006 and 2023 met our inclusion criteria for synthesis (Fig 1).

**Fig 1.**
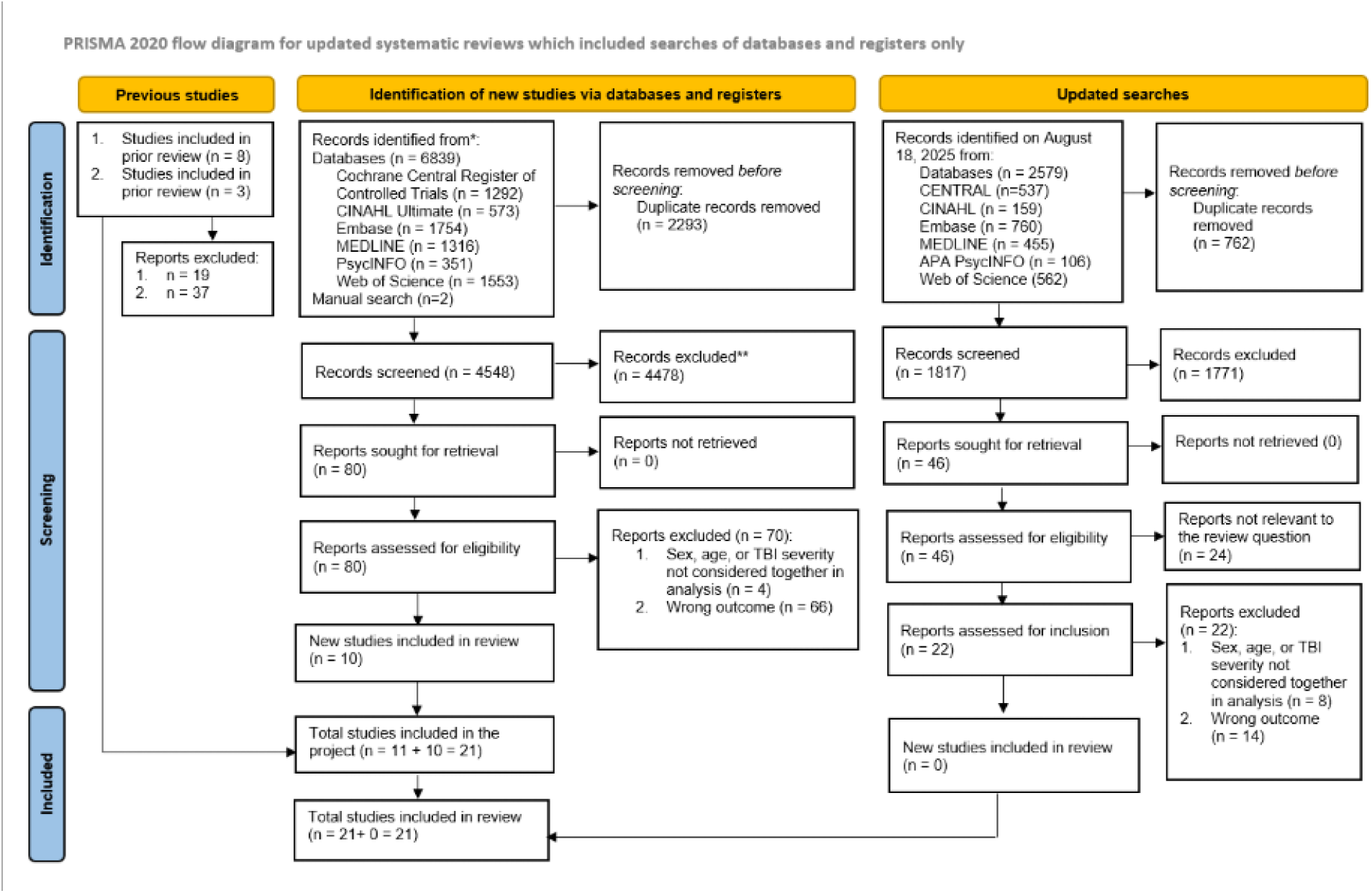
Flow chart for search strategy and study selection. Preferred Reporting Items for Systematic Reviews and Meta-Analyses (PRISMA) diagram.

### Analytical workflow

Fig 2 summarizes systematic review, data extraction and harmonization, meta-analysis, and explainable ML workflow. Detailed results for each analytical stage are presented in the sections below.

**Fig 2.**
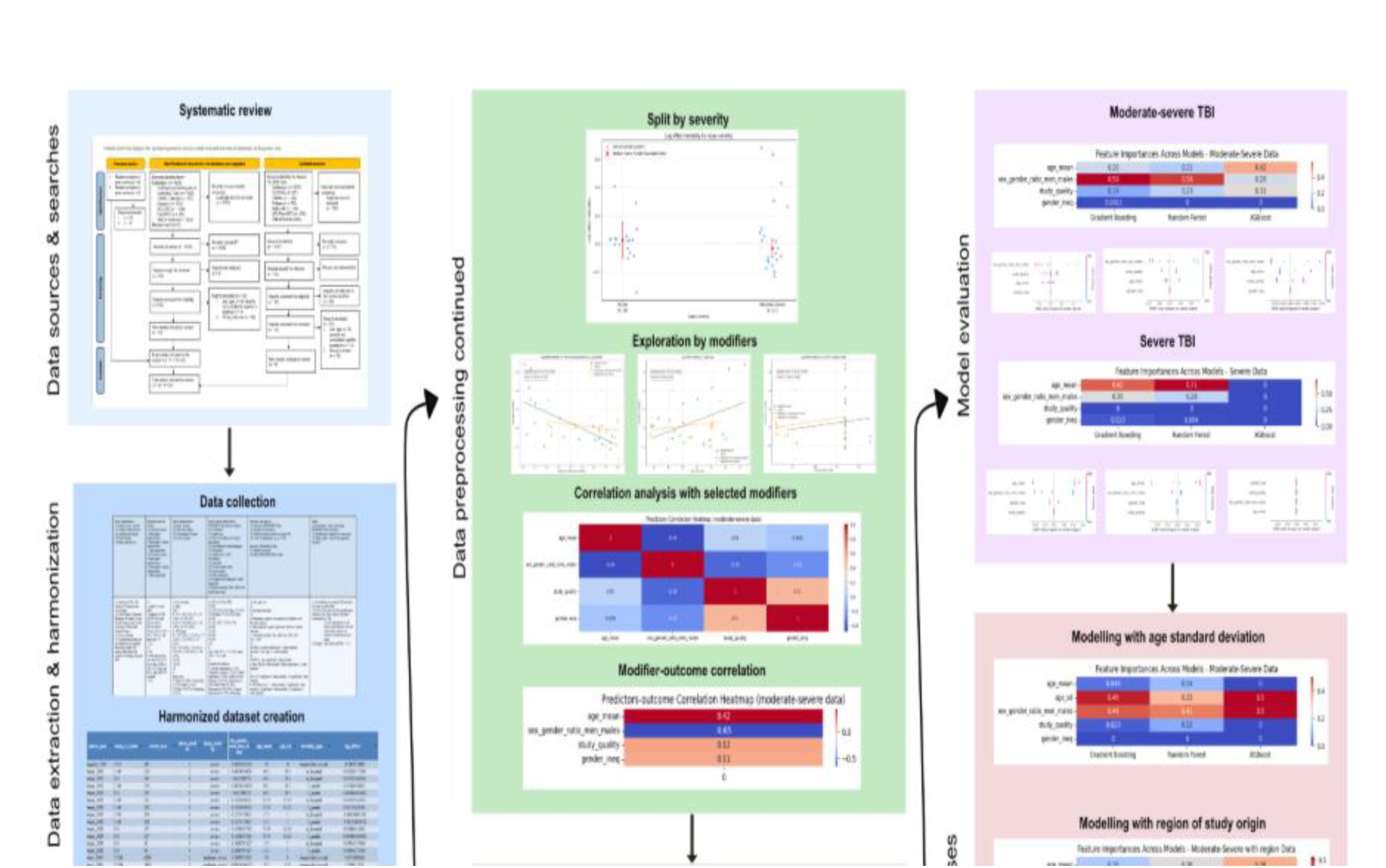
Overview of study design and analytical workflow. Schematic of the seven-stage analytical pipeline, beginning with data collection, extraction and harmonization, data narration and meta-analysis, severity stratification and modifier selection preprocessing, machine-learning modelling, model evaluation, and sensitivity analyses, with intermediate and final results displayed at each stage.

### Study characteristics

A summary of the studies included is presented in Table 1. The 21 studies that met the inclusion criteria consisted of data from 899,813 TBI patients, of whom 48.99% were males (n = 440,794). Among these, 11 (52.38%) were cross-sectional [18, 19, 21, 22, 25, 27, 28, 29, 31, 33, 35] and 10 (47.62%) employed longitudinal study designs [11, 20, 23, 24, 26, 30, 32, 34, 36, 37].

**Table 1.** Study and sample characteristics, including inclusion/exclusion criteria and consideration of PROGRESS-Plus parameters (n = 21)

| Study identification:<br>(1) Author (year):<br>Journal<br>(2) Country;<br>state/province; city;<br>location of research<br>(3) Study Design<br>(4) Study objective (s) | Inclusion/exclusion<br>criteria:<br>(1) Inclusion<br>criteria<br>a. Participants'<br>characteristics<br>b. Participants'<br>clinical<br>characteristics<br>c. Other<br>parameters<br>(2) Exclusion<br>criteria<br>a. Participants'<br>characteristics<br>b. Participants'<br>clinical<br>characteristics<br>c. Other<br>parameters | Injury characteristics:<br>(1) Injury severity<br>(2) Time since injury<br>(3) Circumstance of<br>injury<br>(4) Cause of injury | Study sample<br>characteristics,<br>PROGRESS-Plus factors,<br>country-level indicators:<br>(1) Sample size<br>(2) Place of residence of<br>research participants<br>(3) Race/ethnicity/culture/lang<br>uage<br>(4) Occupation<br>(5) Gender/Sex n (%M)<br>(6) Religion<br>(7) Education<br>(8) Socioeconomic status<br>(9) Social capital<br>(10) Plus parameters<br>(11) Country-level<br>indicators, where<br>applicable:<br>a. Gender Inequality Index<br>(GII) score (publication<br>year) | Measures and analysis:<br>(1) Measure of<br>PROGRESS-Plus<br>(2) Measure of<br>outcome(s)<br>(3) Statistical<br>analysis/analysis controls<br>for<br>(4) Level of significance<br>e.g. p < 0.05)<br>Results by PROGRESS-Plus:<br>(5) Gender/sex results<br>(6) Other PROGRESS-Plus results | Other:<br>(1) Researchers' notes<br>concerning<br>PROGRESS-Plus<br>outcomes<br>(2) Limitations as<br>reported by researchers<br>(3) Study quality / risk<br>of bias appraisal<br>(QUIPS) |
| --- | --- | --- | --- | --- | --- |
| (1) Albrecht (2016):<br>The Journal of Trauma<br>Acute Care Surgery<br>[23]<br>(2) United States;<br>Maryland; Baltimore; R<br>Adams Cowley Shock<br>Trauma Center of the<br>University of Maryland<br>Medical Center<br>(3) Cross-sectional<br>(4) To determine if<br>there are sex<br>differences in mortality<br>following isolated TBI<br>among older adults and<br>compare to findings<br>using all TBI | (1)<br>a. aged 65 y/o and<br>older<br>b. diagnosis of<br>TBI (ICD-9-CM<br>codes 800.xx,<br>801.xx, 803.xx,<br>804.xx, 850.xx-854.1x, 950.1-950.3, 959.01);<br>AIS head score >0<br>c. NA<br>(2)<br>a. NA<br>b. Neck injuries;<br>Pts who died w/in<br>12/24 hrs of adm,<br>all Pts w/ LOS<br><12/24 hrs, and<br>Pts w/ adm SBP<br><90 excluded<br>c. NA | (1) All severities<br>a. Adm<br>GCS:<br>F: 14-15: 418 (71%), 9-13: 83 (14%), 3-8: 89<br>(15%)<br>M 14-15: 496 (68%), 9-13: 103 (14%), 3-8: 131<br>(18%)<br>b. AIS (head):<br>F 1: 120 (20%), 2: 21<br>(4%), 3: 77 (13%), 4:<br>225 (38%), 5: 147 (25%)<br>M; 1: 143 (20%), 2: 16<br>(2%), 3: 70 (10%), 4:<br>305 (42%), 5: 196 (27%)<br>(2) NR<br>(3) NR<br>(4)<br>a.<br>Injury type:<br>F; Blunt 587 (99%);<br>Penetrating 1 (<1%);<br>Other 2 (<1%)<br>M; Blunt 709 (97%);<br>Penetrating 13 (2%)<br>Other 8 (1%)<br>b.<br>Cause of injury:<br>F; MVC 39 (7%);<br>MVC (driver) 18 (46%);<br>MVC (passenger) 12<br>(31%); MVC | (1) 4854 (1320 Isol TBI)<br>(2) NR<br>(3) 76%/76% (F/M) White,<br>17%/19% (F/M) Black,<br>7%/5% (F/M) Other<br>(4) NR<br>(5) Isol: 590 F, 730 M<br>(55%)<br>(6) NR<br>(7) NR<br>(8) NR<br>(9) NR<br>(10)<br>a.<br>Age: mean 78.9 ± 7.7 y/o<br>(F), mean 76.8 ± 7.5 y/o<br>(M)<br>b.<br>Comorbid Conditions:<br>F; Alcohol dependence 14<br>(2%); Alzheimer's disease<br>27 (5%); Cardiac<br>arrhythmia 37 (6%); COPD<br>26 (4%); Diabetes 129<br>(22%); Depression 45<br>(8%); Heart failure 36<br>(6%); Hypertension 346<br>(59%); Ischemic heart<br>disease 37 (6%);<br>Neurologic disorders 34<br>(6%); Stroke 49 (8%)<br>M; Alcohol dependence 47<br>(6%) | (1) Sex, age, race<br>(2)<br>a. In-hospital mortality<br>(3)<br>a. Preliminary analysis:<br>Chi-square test, Student's<br>t-test, Bivariate analysis<br>b. Main analysis:<br>Logistic regression,<br>Stepwise variable<br>selection<br>c. Controlled variables:<br>Sex, Adm year, GCS,<br>AIS<br>(4) p < 0.05<br>(5)<br>Isol TBI: F sex not<br>significant w/ ↓ odds<br>mortality<br>All cases: F sex sign w/ ↓<br>odds mortality<br>(6)<br>Isol TBI: a. Age<br>significant ↑ odds<br>mortality<br>b. Race: Black ↑ odds<br>mortality, Other<br>significant w/ ↓ odds<br>mortality<br>c.<br>GCS: 9-13 significant ↑<br>odds mortality, <9<br>significant ↑ odds | (1) No difference in<br>isolated TBI mortality<br>btw sexes in older adult<br>(2) ICD-9-CM codes for<br>TBI identification<br>instead of AIS codes<br>which could have<br>excluded Pts w/ TBI<br>- Lack of<br>information<br>on pre-<br>injury<br>medication<br>use; was<br>not used in<br>data<br>analysis; no<br>mention of<br>medications<br>post-injury<br>(3) Quality / risk of bias<br>(QUIPS): 7/11, + |
|  |  | (pedestrian) 8 (21%)<br>Falls 535 (91%); Falls On stairs 95 (18%); Falls Different level 75 (14%); Falls Same level 88 (16%); Falls Unspecified 277 (52%)<br>Assault 12 (2%); Other injuries/accident (women) 4 (1%)<br>M; MVC 66 (9%); MVC (driver) 43 (65%); MVC (passenger) 7 (11%); MVC (pedestrian) 8 (12%)<br>Falls 612 (84%); Falls On stairs 92 (15%); Falls Different level 98 (16%); Falls Same level 92 (15%); Falls Unspecified 330 (54%)<br>Assault 34 (5%); Other injuries/accident 18 (2%) | Alzheimer's disease 24 (3%); Cardiac arrhythmia 42 (6%); COPD 36 (5%); Diabetes 145 (20%); Depression 32 (4%); Heart failure 46 (6%); Hypertension 385 (53%); Ischemic heart disease 54 (7%); Neurologic disorders 58 (8%); Stroke 80 (11%)<br>c. Blood Alcohol (Concentration in g/dL): F; Negative 522 (88%); <0.1 8 (1%); 0.1 20 (3%); Missing 40 (7%)<br>M; Negative 614 (84%); <0.1 18 (2%); 0.1 55 (8%); Missing 43 (6%)<br>d. Adm systolic blood pressure (mmHg): F; <90 12 (2%); 90 to 120 41 (7%); 120 to <140 74 (13%); 140 to <160 139 (24%); 160 to <180 121 (21%); 180 203 (34%)<br>M; <90 12 (2%); 90 to 120 43 (6%); 120 to <140 113 (15%); 140 to <160 189 (25%); 160 to <180 171 (23%); 180 205 (28%)<br>e. Admission year: F; 1996-2000 118 (20%); 2001-2005 148 (25%); 2006-2010 216 (37%); 2011-2012 108 (18%)<br>M; 1996-2000 153 (21%); 2001-2005 177 (24%); 2006-2010 264 (36%); 2011-2012 136 (19%)<br>(11) Country-level indicators: a. GII score (publication year): 0.169 (2016) | mortality<br>d. AIS head score: 2 ↑ odds mortality, 3 significant ↑ odds mortality, 4 significant ↑ odds mortality, ≥5 significant ↑ odds mortality<br>All cases: a. Age significant ↑ odds mortality<br>b. Admission year significant w/ ↓ odds mortality<br>c. GCS: 9-13 significant ↑ odds mortality, <9 significant ↑ odds mortality<br>d. AIS head score: 2 ↑ odds mortality, 3 ↑ odds mortality, 4 ↑ odds mortality, ≥5 significant ↑ odds mortality<br>e. ISS: 10-15 significant ↑ odds mortality, 16-25 significant ↑ odds mortality, >25 significant ↑ odds mortality |  |
| (1) Baguley (2012): Medical Journal of Australia [29]<br>(2) Australia, New South Wales Brain Injury Rehabilitation Program<br>(3) Longitudinal<br>(4) To determine the long-term mortality pattern of adults with severe traumatic brain injury (TBI), and to identify the risk factors associated with death in this group. | (1)<br>a. aged ≥ 16-70y/o<br>b. severe TBI (GCS < 9 &/or duration of post-traumatic amnesia > 1 day)<br>c. NA<br>(2)<br>a. NR<br>b. Pts with adm for subsequent brain injuries and secondary adm for reviews & reassessments<br>c. NA | (1) Severe TBI 2545 (100%)<br>(2) NR<br>(3) NR<br>(4)<br>Cause of injury: Motor vehicle accident-related 1442 (58%); Fall/dive 524 (21%); Assault/non-accidental injury 355(14%); Sport/recreation-related 63(3%); Gunshot 15 (1%); Othertraumatic brain injury 65 (3%) | (1) 2545<br>(2) NR<br>(3) NR<br>(4) NR<br>(5) 485 F, 2060 M (81%)<br>(6) NR<br>(7) NR<br>(8) NR<br>(9) NR<br>(10)<br>a. Age: mean 35 ± 14 y/o<br>b. Median LOS for Rehabilitation (IQR): < 30 days 1099 (43%); 31–60 days 546 (22%); ≥ 61 days 900 (35%)<br>(11) Country-level indicators: a. GII score (publication year): 0.056 (2012) | (1) Sex, Age<br>(2) All-cause mortality long-term<br>(3)<br>a. NR<br>b. Main analysis: Standardized mortality ratios (SMRs), Cox proportional hazards regression, Univariate cox regression analysis<br>c. Controlled variables: Age, sex, follow-up-time, discharge functional independence measure (FIM)<br>(4) NR<br>(5) M associated w/ a ↑ mortality compared to F<br>(6)<br>Age is a risk factor; Adults ≥ 36 y/o are 2-3x more likely to die compared to younger adults | (1) M at all ages are associated w/ higher mortality rates compared to F<br>(2)<br>a. Primarily working-age adults with severe TBI,<br>b. Findings about cause of death reported in our study may underestimate true rates, because data on cause of death was undetermined for 18 deaths<br>(3) Quality / risk of bias (QUIPS): 8/14, + |
| (1) Beijer (2023): Journal of Clinical Medicine [30]<br>(2) Netherlands<br>(3) Cross-sectional<br>(4) To examine the association btw sex and | (1)<br>a. aged 16 y/o and older<br>b. severe TBI; admitted to ED w/ AIS ≥ 4 | (1) Severe TBI<br>a. AIS (≥ 3):<br>Total: Neck 17 (1.1%), Spine 90 (5.7%), Thorax 367 (23.4%), Abdomen 42 (2.7%), | (1) 1566 (831 Isol TBI)<br>(2) NR<br>(3) NR<br>(4) NR<br>(5) 520 F, 1046 M (67%) | (1) Sex, age<br>(2)<br>a. In-hospital mortality, 30-day mortality<br>b. GOS ≤ 3 at hospital discharge | (1) NA<br>(2)<br>a. F represented only 33.2% of study population |
| outcome among patients w/ severe TBI | c. NA<br>(2)<br>a. NR<br>b. Patients w/ drowning, asphyxia, or burns<br>c. NA | Lower extremities 110 (7%)<br>F: Neck 3 (0.6%), Spine 21 (4%), Thorax 90 (17.3%), Abdomen 13 (2.5%), Lower extremities 35 (6.7%)<br>M: Neck 14 (1.3%), Spine 69 (6.6%), Thorax 277 (26.5%), Abdomen 29 (2.8%), Lower extremities 75 (7.2%)<br>b. ISS median (IQR):<br>Total: 26 (16-75)<br>F: 25 (16-75)<br>M: 26 (16-75)<br>c. GCS ≤ 8:<br>Total: 829 (52.9%)<br>F: 264 (50.8%)<br>M: 565 (54%)<br>(2) NR<br>(3) NR<br>(4) Injury type:<br>Blunt 1526 (98)<br>Penetrating 31 (2) | (6) NR<br>(7) NR<br>(8) NR<br>(9) NR<br>(10)<br>a. Age: median 60 y/o; 65 (F) y/o, 56 (M) y/o; range 16-98 y/o<br>b. Comorbid Conditions:<br>Healthy or mild (ASA ≤ II): 1077 (68.7)<br>Severe comorbidity (ASA ≥ III): 289 (18.5) | c. ICU adm<br>d. 30-day mortality<br>e. ICU LOS<br>f. hospital LOS<br>g. MV during adm<br>h. MV duration<br>(3)<br>a. Preliminary analysis: Independent student's t-tests, Mann-Whitney U-test, Chi-square test or Fischer's Exact Test<br>b. Main analysis: Logical regression analysis<br>c. Controlled variables: Age, Sex, Mechanism of Injury, ISS, RTS, GCS, SI, Comorbidity, ICP, Craniotomy<br>(4) $p < 0.05$; $p < 0.0167$ for two-sided testing<br>(5)<br>a. No significant differences in in-hospital mortality and 30-day mortality in different sexes.<br>b. M had significantly higher ISS, lower RTS, higher SI, and lower GCS at ED adm<br>c. F were associated with ICU LOS $> 7$ days, but not w/ ICU adm or MV at ED adm<br>(6)<br>a. No significant difference in outcomes or in-hospital mortality btw patients aged 16-44 y/o<br>b. No significant differences in<br>c. More blunt injury mechanisms measure in F than males in isolated TBI | b. No data on reproductive cycles, hormonal status, hormone replacements, or oral contraceptives<br>c. Cause of injury was not explored<br>(3) Quality / risk of bias (QUIPS): 9/11, ++ |
| (1) Berry (2009): The Journal of TRAUMA Injury, Infection, and Critical Care [31]<br>(2) United States<br>(3) Cross-sectional<br>(4) To determine whether gender or menopausal status affects mortality in patients with moderate to severe TBI | (1)<br>a. age $\geq 14$ y/o<br>b. moderate to severe TBI; AIS $\geq 3$ head; AIS $< 3$ all other body regions<br>c. NA<br>(2)<br>a. age $\leq 13$ y/o<br>b. dead on arrival; dead w/ in 24 hrs; transferred out to another facility; admitted with a burn diagnosis; head AIS $> 5$; had any missing data including age, gender, and head AIS<br>c. NA | (1) Moderate to severe TBI<br>a.<br>GCS: Total GCS in ED: $11.1 \pm 4.9$<br>F (mean): $11.7 \pm 4.7$, M (mean): $10.9 \pm 5.0$<br>F (GCS $\leq 8$): 25.6%, M (GCS $\leq 8$): 32.4%<br>b. ISS<br>Total: $21.7 \pm 9.7$<br>F (mean): $21.3 \pm 9.7$, M (mean): $21.8 \pm 9.7$<br>F (ISS $\geq 16$): 76.5%, M (ISS $\geq 16$): 77.9%<br>c. Head AIS<br>Total: $3.8 \pm 0.7$<br>F (mean): $3.7 \pm 0.7$, M (mean): $3.7 \pm 0.7$<br>F (AIS $\geq 3$): 58.0%, M (AIS $\geq 3$): 59.5%<br>(2) NR<br>(3) NR<br>(4)<br>Injury type: Blunt injury: F 98.4%, M 96.2% | (1) 72294<br>(2) NR<br>(3) White 50783 (75.5%); Black 7115 (10.6%); Hispanic 6522 (9.7%); Asian 894 (1.3%); Native American 533 (0.8%); Others 1451 (2.2%)<br>(4) NR<br>(5) 22276 F, 50018 M (69.2%)<br>(6) NR<br>(7) NR<br>(8) NR<br>(9) NR<br>(10)<br>a.<br>Age: mean $44.6 \pm 21.8$ y/o; range 14-89 y/o<br>b. Hospital LOS excluding deaths (d): $11.6 \pm 15.1$<br>c.<br>Comorbid Conditions: Number of comorbidities $0.4 \pm 0.9$<br>Any comorbidity: F 28%, M 23.9% | (1) Sex, age<br>(2)<br>a. All-cause mortality<br>b. Hospital LOS<br>c. ICU LOS<br>d. Ventilated days<br>e. Total FIM score<br>f. Discharge disposition<br>(3)<br>a. Preliminary analysis: Two-sample Wilcoxon's test, Chi-square test<br>b. Main analysis: Binary logistic regression, Stepwise selection in regression, Stratified binary logistic regression<br>c. Controlled variables: Race, Age, Injury type, Systolic blood pressure, ISS, AIS<br>(4) $p < 0.05$; $p < 0.2$ to remain in model<br>(5)<br>a. No significant differences in mortality | (1) NA<br>(2)<br>a. Menopausal status estimated based on age<br>b. No data on hormone replacements or oral, and relationship to timing of injury<br>(3) Quality / risk of bias (QUIPS): 8/11, + |
|  |  |  | (11) Country-level indicators: a. GII score (publication year): 0.169 (2009) | in female patients.<br>b. When adjusting for risk factors by logistic regression, F were associated w/ ↓ mortality.<br>(6) The lowest mortality rate was among peri- and postmenopausal women. |  |
| (1) Clond (2011): The American Journal of Surgery [28]<br>(2) United States; California; Los Angeles<br>(3) Cross-sectional<br>(4) To explore the association btw M and F and SBP in trauma pts and establish how the difference btw M and F influenced outcomes in those w/ elevated SBP† | (1)<br>a. age ≥ 14 y/o<br>b. moderate to severe TBI; (head AIS ≥ 3)<br>c. NA<br>(2)<br>a. age ≤ 14 y/o<br>b. dead on arrival; AIS > 5 for any body region; missing data on age, gender, AIS, ISS < GCS, or blood alcohol status<br>c. NA | (1) Moderate to severe TBI<br>a.<br>GCS: F (mean): 11.5 ± 4.3<br>M (mean): 11.4 ± 4.4<br>F (GCS ≤ 8): 26.2%<br>M (GCS ≤ 8): 26.4%<br>b.<br>ISS: F (mean): 22.2 ± 11.6<br>M (mean): 21.3 ± 11.1<br>F (ISS ≥ 16): 70.4%<br>M (ISS ≥ 16): 73.6%<br>c.<br>AIS: AIS head: F (mean): 3.77 ± 0.3<br>M (mean): 3.76 ± 0.79<br>(2) NR<br>(3) NR<br>(4) NR | (1) 3025 TBI (24184 non-TBI)<br>(2) NR<br>(3) NR<br>(4) NR<br>(5) 607 F, 2418 M (79.9%)<br>(6) NR<br>(7) NR<br>(8) NR<br>(9) NR<br>(10)<br>Age: mean 40.7 ± 17.3 y/o (F), mean 44.8 ± 21.6 y/o (M)<br>(11) Country-level indicators: a. GII score (publication year): 0.169 (2011) | (1) Sex, age<br>(2)<br>a. In-hospital mortality<br>(3)<br>a. Preliminary analysis: Student's t test, Wilcoxon rank sum test<br>b. Main analysis: Multivariate linear models, Multivariate regression models<br>c. Controlled variables: Age, Sex, ISS, GCS, adm SBP<br>(4) p < 0.05<br>(5) No significant differences btw the mortality rates in the different sexes when pneumonia was unadjusted in both the TBI and non-TBI groups.<br>(6) Significant predictors of mortality in all groups included age, ISS 16, GCS 8, and SBP 90 mmHg; SBP 160mmHg was a significant predictor of ↑ mortality in M TBI pts but was not significant for F with TBI or non-TBI men. | (1) Elevated SBP was associated with ↑ pneumonia; increased SBP in TBI M ↑ mortality<br>(2) No direct cause-and-effect comparisons can be made for SBP mediated gender-related differences<br>(3) Quality / risk of bias (QUIPS): 8/11, + |
| (1) Davis (2006): Journal of Neurotrauma [32]<br>(2) United States<br>(3) Cross-sectional<br>(4) To explore the relationship btw M and F and outcome in adult patients with moderate-to-severe TBI using a large trauma registry | (1)<br>a. NR<br>b. moderate to severe TBI; (head/neck AIS ≥ 3)<br>c. NA<br>(2)<br>a. NR<br>b. Head/Neck AIS defined neck injuries<br>c. NA | (1) Moderate to severe TBI:<br>F: moderate to severe 3178, M: moderate to severe 10259<br>(2) NR<br>(3) NR<br>(4) NR | (1) 13437<br>(2) NR<br>(3) NR<br>(4) NR<br>(5) 3178 F, 10259 M (76.4%)<br>(6) NR<br>(7) NR<br>(8) NR<br>(9) NR<br>(10)<br>Age: range 15-49 y/o<br>(11) Country-level indicators: a. GII score (publication year): 0.169 (2006) | (1) Sex, age<br>(2) All-cause mortality<br>(3)<br>a. NR<br>b. Main analysis: Logistic regression, repeated logistic regression, Revised Trauma Score and Injury Severity Score (TRISS) methodology<br>c. Controlled variables: Age, mechanism of injury, pre-admission GCS, pre-admission hypotension (SBNP ≤ 90mmhg), head AIS, ISS<br>(4) p < 0.05<br>(5)<br>a. <50 y/o, no significant differences btw F or M mortality. F ≥ 50 years mortality is significantly ↓ than M.<br>b. Pre-menopausal F no significant difference in outcome compared to M<br>Post-menopausal F significant difference compared to M<br>c. F & M divergent differences shown btw 40-49 y/o and 50-59 y/o<br>(6) NR | (1) Improved outcomes in menopausal years of older F not explained by hormonal factors as w/ younger F<br>(2)<br>a. Logistic regression may not adjust for all factors<br>b. Hormone levels and menstrual history for F pts were undetermined<br>c. Unable to determine the use of oral contraceptives in younger F pts, hormone replacement therapy after menopause, and continuation/discontinuation<br>(3) Quality / risk of bias (QUIPS): 8/11, + |
| (1) de Guise (2014): International Scholarly Research Notices [24]<br>(2) Canada; Quebec; Montreal; McGill University Health Center<br>(3) Cross-sectional<br>(4) To examine gender differences in short-term acute care outcomes and compare demographic, clinical, and psychosocial characteristics between men and women with traumatic brain injury (TBI) | (1) Inclusion criteria<br>a. Pts admitted to TBI Program at MUHC-Montreal General Hospital between 2000-2011<br>b. Physician-diagnosed TBI<br>c. NA<br>(2) Exclusion criteria<br>a. NR<br>b. NR<br>c. NA | Injury Characteristics<br>(1) GCS Severity: F mild (13-15): 1277 (73.9%), moderate (9-12): 166 (9.6%), severe (3-8): 285 (16.5%)<br>M<br>mild (13-15): 2969 (68.9%), moderate (9-12): 395 (10.1%), severe (3-8): 823(21.0%)<br>(2) Acute hospitalization period (adm to discharge)<br>(3) Falls, Work accidents, Assaults, MVC<br>(4) Cause of injury:<br>F<br>Falls 963(55.7%), MVC 679 (39.3%), Work accidents 16 (0.9%), Assaults 70 (4.0%)<br>M<br>Falls 1948 (49.8%), MVC 1186 (30.3%), Work accidents 259 (6.6%), Assaults 521 (13.3%) | Characteristics of study sample by PROGRESS-Plus: (1) 5,642<br>(2) NR<br>(3) NR<br>(4)NR<br>(5) 1, 728 F, 3, 914 M (69.4%)<br>(6) NR<br>(7)<br>F: elementary (11.6%), high school (41.7%), college (16.1%), university (30.6%)<br>M: elementary (12.1%), high school (50.7%), college (13.7%), university (23.5%)<br>72.2% NR<br>(8) NR<br>(9) NR<br>(10)<br>a.<br>Age: F: 57.2 ± 23.6 y/o, M: 47.6 ± 20.8 y/o<br>b. Comorbid conditions: Positive brain CT, substance abuse, prior neurological conditions, intellectual impairments, severe behavioural issues, psychiatric diagnoses, dementia, prior suicide attempts, and homelessness (individual n (%) NR).<br>(11) Country-level indicators: a. GII score (publication year): 0.052 (2014) | (1) Sex, age, education<br>(2)<br>a. Hospital LOS<br>b. In hospital mortality<br>c. GOSE at discharge<br>d. FIM at discharge<br>e. discharge destinations<br>(3) a. Preliminary analysis: Kruskal-Wallis test, Chi-square test, Student's t-test<br>b. Main analysis: ordinal regression, logistic regression<br>c. controlled variables: age, year of accident, GCS, ISS, primary language, leaving against medical advice, mechanism of injury<br>(4) p < 0.05<br>(5) a. F ↑ rates of falls and MVC, M ↑ rates of assaults and work-related injuries<br>b. F ↓ injury severity than M<br>c. post-adjustment, F ↓ odds of unfavorable functional outcomes and likely to be discharged home<br>d. no significant sex differences observed for mortality<br>(6) a. ↑ age associated w/ poorer outcomes<br>b. higher injury severity (GCS/ISS) associated w/ worse outcomes | (1) F and M differed considerably in demographic and injury profiles, older age among F may partially explain outcome differences<br>(2) a. Retrospective single-center study design<br>b. Missing education data (72.2%)<br>c. Data obtained from registry and medical records may be incomplete<br>d. Limited generalizability beyond one Level 1 trauma center<br>e. Potential residual confounding from unmeasured variables<br>(3) Quality / risk of bias (QUIPS): 8/11, + |
| (1) Donohue (2007): The Journal of TRAUMA Injury, Infection, and Critical Care [33]<br>(2) United States<br>(3) Longitudinal<br>(4) The goal of this study was to determine whether advanced age affected the long-term survival of patients with a traumatic brain injury (TBI). | (1)<br>a. aged ≥ 65 y/o<br>b. Pts admitted for the first time w/ head injury (ICD-9 800 – 804 or 850 – 854)<br>c. NA<br>(2)<br>a. NR<br>b. NR<br>c. NA | (1) Severe TBI (AIS ≥ 3)<br>AISmax: AISmax 3 7,642 (36.31%); AISmax 4 12,207 (58.01%); AISmax 5 1,195 (5.68%)<br>(2) NR<br>(3) NR<br>(4) NR | (1) 30684; 21,044 pts w/ AISmax of at least 3 and survived past the first day<br>(2) NR<br>(3) NR<br>(4) NR<br>(5) 11,089 F, 9,955 M (47.31%)<br>(6) NR<br>(7) NR<br>(8) NR<br>(9) NR<br>(10)<br>a.<br>Age: range ≥65 y/o<br>b. Charlson Comorbidity Index: 0, 12414 (50.31%); 1, 5711 (22.56%); 2, 1968 (12.88%); ≥3, 951 (%)<br>(11) Country-level indicators: a. GII score (publication year): 0.169 (2007) | (1) Sex, age<br>(2) In-hospital, 12-month mortality; 24-month mortality<br>(3)<br>a. NR<br>b. Main analysis: Multivariate logistic regression<br>c. Controlled variables: Age, Sex, AIS, Loss of consciousness, Comorbidity factors (Charlson Comorbidity Index)<br>(4) p < 0.05<br>(5) Long-term mortality ↑ in M<br>(6) ↑ in age ↑ rate of long-term mortality, ↑ comorbidities, or AISmax | (1) ↑ risk of mortality much higher in older injured pts<br>(2) NR<br>(3) Quality / risk of bias (QUIPS): 9/14, + |
| (1) Fortuna (2008): Surgery [34]<br>(2) United States, Ohio, Cincinnati, Level 1 Trauma Center at the University of Cincinnati<br>(3) Cross-sectional<br>(4) To determine whether preinjury Clopidogrel/aspirin/wafarin use was an | (1)<br>a. age ≥ 50 y/o<br>b. blunt head trauma<br>c. NA<br>(2)<br>a. age ≤ 50 y/o<br>b. No hBI; non-blunt HBI (e.g. concussion & facial/skull | (1) All severities<br>a.<br>ISS: Clopidogrel 23.9 ± 2.7<br>Aspirin 24.5 ± 1.1<br>Clopidogrel/aspirin 18.2 ± 1.6<br>Warfarin 24.11 ± 1.1<br>Other 25.2 ± 2.7<br>None 0.7 ± 24.9<br>b. Head | (1) 416<br>(2) NR<br>(3) NR<br>(4) NR<br>(5) 163 F, 253 M (60.8%)<br>(6) NR<br>(7) NR<br>(8) NR<br>(9) NR<br>(10)<br>a. | (1) Sex, age<br>(2) In-hospital mortality<br>(3)<br>a. Preliminary analysis: Chi-square test, t-test, two-way anova, Tukey's post-hoc test<br>b. Main analysis: Multiple logistic regression<br>c. Controlled variables: | (1) Pts who were older were more likely to receive platelet therapy, higher injured severity, and greater aspirin or clopidogrel use history before injury<br>(2)<br>a. retrospective design<br>b. exclusion of pts w/ incomplete |
| important predictor of mortality in patients aged >50 y/o with blunt, hemorrhagic brain injury (HBI). | fractures); pts w/ incomplete documentation of outpatient clopidogrel, aspirin, or warfarin use<br>c. NA | AIS: Clopidogrel 4.0 ± 0.2<br>Aspirin 4.1 ± 0.1<br>Clopidogrel/aspirin 3.9 ± 0.2<br>Warfarin 4.4 ± 0.1<br>Other 4.5 ± 0.2<br>None 4.1 ± 0.04<br>c.<br>GCS: Clopidogrel 11.8 ± 1.3<br>Aspirin 12.5 ± 0.4<br>Clopidogrel/aspirin 13.3 ± 0.8<br>Warfarin 12.2 ± 0.9<br>Other 14.3 ± 0.6<br>None 12.0 ± 0.3<br>(2) NR<br>(3) NR<br>(4)<br>a.<br>Cause of injury: Fall from a height 245.44 (59%), MVC 95.68 (23%), Pedestrian struck 29.12 (7%)<br>b. Cause of injury by subgroup: 50-70 y/o<br>Clopidogrel/aspirin/warfarin: MVC 13 (25%), Fall 29 (56%), Assault 2 (4%), MCC 3 (6%), Pedestrian 2 (4%), Other 2 (4%), Unknown 1 (2%)<br>50-70 y/o<br>Clopidogrel/aspirin/warfarin: MVC 41 (27%), Fall 63 (42%), Assault 7 (5%), MCC 15 (10%), Pedestrian 13 (9%), Other 9 (6%), Unknown 2 (1%)<br>>70 y/o +<br>Clopidogrel/aspirin/warfarin: MVC 19 (17%), Fall 88 (77%), Assault 2 (2%), MCC 0 (0%); Pedestrian 4 (4%); Other 0 (0%); Unknown 1 (1%)<br>>70 y/o -<br>Clopidogrel/aspirin/warfarin: MVC 21 (21%); Fall 64 (64%); Assault 1 (1%); MCC 1 (1%); Pedestrian 11 (11%); Other 1 (1%); Unknown 0 (0%) | Age: mean 69 ± 1 y/o; range age ≥ 70 y/o (51%), range age 50-70 y/o (49%)<br>b. LOS (days): Clopidogrel 7.8 ± 1.6<br>Aspirin 7.6 ± 0.8<br>Clopidogrel/aspirin 4.1 ± 1.5<br>Warfarin 9.6 ± 1.7<br>Other 6.5 ± 1.9<br>None 7.9 ± 0.6<br>(11) Country-level indicators: a. GII score (publication year): 0.169 (2008) | Age, Sex, ISS, GCS, CAW (yes/no)<br>(4) p < 0.05<br>(5)<br>a. Mortality significantly ↑ for older CAW(-) pts; no significant differences btw older and younger CAW(+) pts in mortality.<br>b. No differences in mortality btw sexes<br>(6) Age was a significant predictor of mortality | documentation of clopidogrel, aspirin, or warfarin use<br>(3) Quality / risk of bias (QUIPS): 10/11, ++ |
| (1) Han (2017): Journal of Korean Neurosurgical Society [35]<br>(2) South Korea<br>(3) Longitudinal<br>(4) To evaluate the associations between 30-day mortality and various radiological and clinical factors in young patients with no deaths due to medical comorbidities or other organ problems. | (1)<br>a. Aged 15-65 y/o<br>b. Traumatic acute subdural hematoma<br>c. NA<br>(2)<br>a. Aged <15 y/o and > 65 y/o<br>b. Pts w/ surgery performed ≥ 48 hrs after head trauma excluded<br>c. NA | (1) All severities<br>GCS: 3-5, 41 (12.9%); 6-8, 159 (50.0%); 9-15, 118 (37.1%)<br>(2) NR<br>(3) NR<br>(4)<br>Cause of injury: Traffic accident 190 (59.7%), Fall 99 (31.1%), Assault or others 29 (9.1%) | (1) 318<br>(2) NR<br>(3) NR<br>(4) NR<br>(5) 78 F; 240 M (75.5%)<br>(6) NR<br>(7) NR<br>(8) NR<br>(9) NR<br>(10)<br>a.<br>Age: mean 47.8 ± 12.7 y/o<br>b.<br>Comorbid Conditions: Hypertension 109 (34.3%), Diabetes 48 (15.1 %), Congestive heart disease 5 (1.6%), Arrhythmia 14 (4.4%), Liver disease 12 (3.8%), Renal failure 3 | (1) Sex, Age<br>(2) 30-day mortality<br>(3)<br>a. Preliminary analysis: Student's t test, Chi-square test<br>b. Main analysis: Kaplan-Meier survival curves, Log-rank test, univariate cox proportional hazards regression, multivariate cox proportional hazards regression<br>c. Controlled variables: Age, Sex, radiologic findings, and patient clinical factors<br>(4) p < 0.05<br>(5) F had ↓ mortality risk than M but this was not | (1) No associations between 30-day mortality and patient age and gender<br>(2)<br>a. Pts older than 65 were excluded which may explain lower mortality rate in this study<br>b. inconsistencies in the duration between the immediate postoperative CT scan and the follow-up CT scan. Therefore, we could not accurately evaluate the cause of death.<br>(3) Quality / risk of bias (QUIPS): 7/14, + |
|  |  |  | (0.9%), Chronic obstructive pulmonary disease 4 (1.3%), History of stroke 8 (2.5%)<br>(11) Country-level indicators: a. GII score (publication year): 0.038 (2017) | significant<br>(6) ↑ in age ↑ risk of mortality but this was not significant |  |
| (1) Harrison-Felix (2012): Journal Of Head Trauma Rehabilitation [25]<br>(2) United States<br>(3) Longitudinal<br>(4) To characterize overall and cause-specific mortality and life expectancy among persons who have completed inpatient traumatic brain injury rehabilitation and to assess risk factors for mortality. | (1)<br>a. aged ≥ 16y/o<br>b. meet at least 1 of the following criteria for moderate to severe TBI: posttraumatic amnesia greater than 24 hrs, trauma-related intracranial neuroimaging abnormalities, loss of consciousness exceeding 30 mins, or a GCS score in the emergency department of <13 (unless due to intubation, sedation, or intoxication); present to the Model System's acute care hospital within 72 hrs of injury<br>c. provide informed consent to participate or have a proxy provide consent<br>(2)<br>a. NR<br>b. NR<br>c. NA | (1) All severities GCS at ED adm: severe, GCS 3-8 3094.85 (36.1%); moderate, GCS 9-12 1071.63 (12.5%); mild, GCS 13-15 2426.16 (28.3%); Sedated 1757.47 (20.5%) Missing 222.9 (2.6%)<br>(2) NR<br>(3) NR<br>(4)<br>Cause of injury: Vehicular 4672.29 (54.5%), Fall 1826.05(21.3%), Violence 1157.36 (13.5%), Pedestrian 617.27 (7.2%), Sports or recreational accidents 128.6 (1.5%), Other (including falling object) 145.74 (1.7%) | (1) 8573<br>(2) NR<br>(3) Non-Hispanic white 5709.62 (66.6%), Black 2400.44 (20.8%), Hispanic origin 728.71 (8.5%), Asian/Pacific Islander 231.47 (2.7%), Other (including Native American) 120.02 (1.4%)<br>(4) NR<br>(5) 2246 F, 6327 M (73.8%)<br>(6) NR<br>(7) NR<br>(8) NR<br>(9) NR<br>(10)<br>a. Age: mean 39.0 ± 18.4 y/o<br>b. Days from injury to death 3.5/2 (3.7)<br>(11) Country-level indicators: a. GII score (publication year): 0.169 (2012) | (1) Sex, Age, Race, Marital Status, Employment Status<br>(2) Overall and cause-specific mortality<br>(3)<br>a. Preliminary analysis: Student's t test, Chi-square test<br>b. Main analysis: Standardized mortality ratios (SMRs), Cumulative Conditional Probability of Survival, Life expectancy estimation, multivariate cox proportional hazards regression<br>c. Controlled variables: Age, Sex, race/ethnic group, level of education, marital status, employment status at injury, pre-injury drug use, GCS, PTA, unconsciousness duration, SCI, cause of injury, LOS< payer sources, discharge destination, FIM, DRS<br>(4) p < 0.05<br>(5) F ↓ in mortality risk than M<br>(6) a. Age significant ↑ mortality risk<br>b. Race: Hispanic and Other (including Native American) ↓ mortality risk, Black and Asian ↓ mortality risk<br>c. Marital status at injury: Divorced/widowed significant ↑ mortality risk<br>Never married ↓ mortality risk<br>d. Employment status at injury: Unemployed, Retires, and Other (homemaker/volunteer/other) significant ↑ mortality risk; Student (part-time/full/special education) ↓ mortality risk | (1) Moderate and severe TBI ↑ mortality risk for all ages, both sexes, all race/ethnic groups (except Native Americans), and all injury severity groups<br>(2)<br>a. the outcomes may not be generalizable to individuals with TBI receiving inpatient rehabilitation treatment elsewhere.<br>b. This study did not include those who did not require hospitalization or did not receive inpatient rehabilitation.<br>c. Fifteen percent of the deaths identified in this study were missing the cause of death information, which could result in an underestimation of the prevalence of some causes<br>(3) Quality / risk of bias (QUIPS): 10/14, + |
| (1) Hong (2022): European Journal of Trauma and Emergency Surgery [38]<br>(2) United States of America; National Trauma Data Bank<br>(3) Cross-sectional<br>(4) To investigate the combined effects of age and sex on outcomes following moderate to severe TBI and to determine whether premenopausal females have better outcomes than males | (1)<br>a. Adults > 16 y/o<br>b. Isolated moderate to severe blunt TBI, head AIS > 2<br>c. NTDB records from 2007 – 2016<br>(2) Exclusion criteria<br>a. Age ≤ 16 y/o<br>b. Mild TBI (head AIS ≤ 2), Polytrauma (AIS > 2 in other brain regions)<br>c. Non-isolated TBI | (1) Moderate to severe TBI, Head AIS > 2<br>(2) Acute hospitalization period<br>(3) Blunt trauma<br>(4) NR | (1) 686, 549<br>(2) United States of America<br>(3)<br>Aged < 45 y/o<br>F: White 72.2%, Black 10.6%, Asian 2.2%, American Indian 1.3%; M: White 64.7%, Black 12.8%, Asian 1.8%, American Indian 1.4%<br>Aged 45- 55 y/o<br>F: white (77.4%); M: white (69.0%)<br>Aged >55 y/o<br>F: white (84.2%); M: white (79.8%)<br>(4) NR<br>(5) 134 F, 436, 058 M (63.4%)<br>(6) NR<br>(7) NR<br>(8) NR<br>(9) NR<br>(10)<br>a. Age groups:<br>Premenopausal: < 45 y/o, Perimenopausal: 45-55 y/o, Postmenopausal: 55 y/o<br>b.<br>Comorbid Conditions: Smoking, CKD, Diabetes, CAD, Hypertension, Obesity, Respiratory disease, Cirrhosis<br>(11) Country-level indicators: a. GII score (publication year): 0.169 (2022) | (1) Sex, Age, Race, Comorbidities<br>(2)<br>Primary outcomes: mortality<br>Secondary outcomes: Complications, Hospital LOS, ICU LOS, Mechanical ventilation days<br>(3) Statistical tests: Chi-square test, Mann-Whitney U test, Multivariate logistic regression<br>Controlled variables: ISS, Race, Mechanism of injury, Smoking, CKD, Diabetes, CAD, Hypertension, Obesity, Respiratory disease, Cirrhosis<br>(4) p < 0.05<br>(5)<br>< 45 years: mortality similar between sexes, females had fewer complications, shorter hospital, ventilation and ICU stays<br>45-55 y/o: Mortality similar in unadjusted analysis, after adjustment, F had slightly ↑ mortality risk > 55 y/o: F had ↓ mortality than M<br>(6)<br>a. ↑ age associated with increased mortality<br>b. Smoking and obesity were associated with ↓ mortality risk<br>c. F > 65 y/o had greatest survival advantage | (1) Postmenopausal F had better outcomes than M, Premenopausal F did not demonstrate a protective effect, Findings do not support endogenous female sex hormones as the explanation for improved outcomes<br>(2)<br>a. Menopausal status was estimated using age categories<br>b. Oral contraceptive, serum hormone and hormone replacement therapy<br>c. Projective studies needed<br>(3) Quality / risk of bias (QUIPS): 9/11, ++ |
| (1) Hosomi (2021): BMC Neurology [36]<br>(2) Japan; Japanese Trauma Data Bank (JTDB)<br>(3) Cross-sectional<br>(4) To investigate sex differences in mortality and complications following TBI, to examine how age modifies the association between sex and TBI outcomes in Japan | (1)<br>a. Pts with TBI registered in JTDB<br>b. TBI requiring trauma centre care<br>c. JTDB study period (2013-2018)<br>(2)<br>a.<br>Age < 10 y/o<br>b.<br>Cardiopulmonary arrest on arrival<br>c.<br>Duplicate/incomplete records | (1) All TBI severities included, severities adjusted using AIS and GCS<br>(2) Acute hospitalization period<br>(3) NR<br>(4) Falls, Motor vehicle collisions, other trauma mechanisms | (1) 51, 726<br>(2) Japan<br>(3) NR<br>(4) NR<br>(5) 18, 756 F, 32, 907 M (63.7%)<br>(6) NR<br>(7) NR<br>(8) NR<br>(9) NR<br>(10)<br>a. Medication variables: collected and adjusted for anticoagulants and antiplatelet agents<br>b. Injury severity variables: AIS, GCS<br>(11) Country-level indicators: a. GII score (publication year): 0.059 (2021) | (1) Sex, Age, Medication Use, Injury severity<br>(2) Primary outcome: In-hospital mortality, Secondary outcome: neurological complications, non-neurological complications<br>(3)<br>a. Statistical analyses: Chi-square tests, logistic regression<br>b. Controlled variables: Age, Injury severity, medication use,<br>(4) p < 0.05<br>(5)<br>a. M had ↑ mortality than F<br>b. Mortality differences greatest in: 10-19 y/o, 60-69 y/o, 70-79 y/o, 80-89 y/o, 90-99 y/o<br>(6)<br>a. Elderly M experienced more complications<br>b. M had greater odds of neurologic complications (OR 1.29) and non-neurologic complications (OR 1.14) | (1)<br>a. Sex disparity was age-dependent<br>b. Elderly M experienced poorest outcomes<br>c. Findings suggest sex differences extend beyond reproductive age<br>(2)<br>a. Retrospective registry design<br>b. Residual confounding possible<br>c. Menopausal status, hormone levels, long-term outcomes unavailable<br>d. Data limited to hospitalized patients<br>(3) Quality / risk of bias (QUIPS): 8/11, + |
| (1) Mikolic (2020): Journal of Neurotrauma [37]<br>(2) European Centers<br>(3) Longitudinal<br>(4) To investigate the associations of sex/gender and functional and mental outcomes as well as HRQoL after TBI, Research concerns sex differences | (1)<br>a. Adults<br>b. Isol TBI; moderate to severe TBI; no additional injury AIS > 2; survived at least until admission to ICU<br>c. NA<br>(2)<br>a. NR<br>b. NR<br>c. NA | (1) All severities<br>(2) NR<br>(3) NR<br>(4)<br>a.<br>Cause of injury: mTBI: F; Incidental fall 570 (56.4%); MVA 330 (32.7%); Other 70 (6.9%); Violence 40 (4%)<br>M; Incidental fall 824 (45.7%); MVA 647 (35.8%); Other 173 (9.6%); Violence 161 (8.9%);<br>Moderate/severe TBI: F; Incidental fall 151 (44.3%)<br>MVA 159 (46.6%); Other 17 (5%); Violence 14 (4.1%)<br>M; Incidental fall 358 (37.9%); MVA 436 (46.2%); Other 85 (9%); Violence 65 (6.9%) | (1) 4195<br>(2) NR<br>(3) NR<br>(4) Change in Employment Status: a. mTBI: F; Job Change 26 (4.2%); Not Returned 110 (17.8%); Retired 206 (33.3%); Returned (Women) 277 (44.7%)<br>M; Job Change 48 (4.3%); Not Returned 203 (18.1%); Retired (Men) 260 (23.2%); Returned 610 (54.4%)<br>b. Moderate to Severe TBI: F; Job Change 5 (2.9%); Not Returned 96 (55.5%); Retired 37 (21.4%); Returned 35 (20.2%)<br>M; Job Change 26 (5.4%); Not Returned 295 (61.7%); Retired 52 (10.9%); Returned 105 (22%)<br>(5) mTBI: 1020 F, 1842 M (64%); moderate to severe TBI: 353 F, 980 M (74%)<br>(6) NR<br>(7) NR<br>(8) NR<br>(9) NR<br>(10)<br>Age: mTBI median 58 (F), 50 (M); moderate to severe TBI median 53 (F), 47 (M)<br>(11) Country-level indicators: a. GII score (publication year): 0.226 (2020) | (1) Sex, age, occupation<br>(2) In-hospital mortality; 6-month mortality<br>(3)<br>a. Preliminary analysis: Chi-square test, Mann-Whitney U test<br>b. Main analysis: Univariable and Multi-variable mixed effects regression, logistic mixed effects regression models, ordinal mixed effects regression, linear regression, complete-case analysis and multiple imputation<br>c. Controlled variables: Age, Baseline GCS, Pupillary reactivity, Hypotension (pre-admission or at admission), Hypoxia (pre-admission or at admission), CT abnormalities, ISS< Pre-injury medical situation, Pre-injury psychiatric disorder, Cause of injury<br>(4) $p < 0.05$<br>(5)<br>a. F have ↓ associated risk for in-hospital associated mortality and at 6 months.<br>b. F ↑ rates of falls, more moderate/severe TBI due to falls<br>c. M ↑ rates of moderate/severe TBI due to violence & other reasons<br>d. F <5 and >65 y/o had worse outcomes compared to M<br>(6)<br>a. Most common cause of injury was falls<br>b. Pts <45 y/o had largest sex difference in not returning to work | (1) M and F differ in care pathway and outcomes, depending on injury severity and age group<br>(2)<br>a. High proportion of missing 6-month outcomes values, especially for moderate-severe group & M<br>b. Potential ↑ in probability of false-positives due to testing for multiple end-points<br>c. Adjustment could miss important confounders<br>d. Intersex & cisgender persons not captured in detail<br>(3) Quality / risk of bias (QUIPS): 10/14, + |
| (1) Ng (2006): British Journal of Neurosurgery [11]<br>(2) NR<br>(3) Longitudinal<br>(4) The objective of this study was to investigate if there are possible gender differences in relation to outcome following closed severe traumatic brain injury (TBI) in a predominantly Asian population. | (1)<br>a. NR<br>b. severe TBI (GCS <8); included pts who initially admitted w/ GCS >8 but decreased once at neurosurgical ICU<br>c. NA<br>(2)<br>a. NR<br>b. NR<br>c. NA | (1) Severe TBI (GCS <8) 672 (100%)<br>(2) NR<br>(3) NR<br>(4)<br>Cause of injury: Road traffic accidents 308 (45.8%); Fell from height 108 (16.1%); Sustained simple falls 184 (27.4%); Assaults 20 (3%); Other injuries 41 (6.1%); Unknown causes 5 (0.7%); Sports or recreational accidents 6 (0.9%) | (1) 672<br>(2) NR<br>(3) NR<br>(4) NR<br>(5) 147 F, 525 M (78.1%)<br>(6) NR<br>(7) NR<br>(8) NR<br>(9) NR<br>(10)<br>Age: mean $53.7 \pm 22.8$ y/o (F), $43 \pm 18.6$ y/o (M); median 58 y/o (F), 40 y/o (M)<br>(11) Country-level indicators: a. GII score (publication year): 0.031 (2006) | (1) Sex<br>(2) 6-month mortality<br>(3)<br>a. Preliminary analysis: Chi-square test, two-sample test<br>b. Main analysis: Logistic regression<br>c. Controlled variables: Age, Multiple injuries, Post-resuscitation pupil abnormalities, GCS<br>(4) $p < 0.05$<br>(5)<br>a. F ↑ likelihood of mortality after severe TBI compared to M for crude model<br>b. No significant difference in mortality btw M & F after adjusting for age, post-resuscitation GCS, pupil abnormality & multiple injuries<br>(6) No significant<br>differences for results<br>btw ages <60 & >60 y/o<br>btw sexes | (1) No significant differences in poorer outcomes or higher mortality in F after adjustments for age, post-resuscitation GCS, pupil abnormality & multiple injuries<br>(2) Use of age of 60 as cut-off to separate females into pre-and postmenopausal groups is arbitrary<br>(3) Quality / risk of bias (QUIPS): 9/14, + |
| (1) Ottochian (2008):<br>The American Journal<br>of Surgery [38]<br>(2) United States,<br>California, Los Angeles<br>County and University<br>of Southern California<br>Medical Center<br>(3) Cross-sectional<br>(4) To examine<br>outcomes btw the sexes<br>after blunt TBI. | (1)<br>a. younger than 14<br>years, 14 to 44<br>years, 45 to 54<br>years, and 55<br>years and older<br>b. Isol severe TBI<br>(AIS > 3, w/ AIS<br>≤ 3 for all other<br>body regions<br>(chest, abdomen,<br>& extremity)<br>c. NA<br>(2)<br>a. NR<br>b. Pts w/ non-<br>survivable head<br>injuries (head<br>AIS, 6)<br>c. Pts transferred<br>to another acute<br>care facility<br>before 1 week | (1) Severe TBI AIS <3<br>1807 (100%)<br>(2) NR<br>(3) NR<br>(4) NR | (1) 1807 Isol TBI<br>(2) NR<br>(3) NR<br>(4) NR<br>(5) 406 F, 1401 M<br>(77.53%)<br>(6) NR<br>(7) NR<br>(8) NR<br>(9) NR<br>(10)<br>Age: mean 45.3 ± 22.1 y/o<br>(11) Country-level<br>indicators: a. GII score<br>(publication year): 0.169<br>(2008) | (1) Sex, age<br>(2) 6-month mortality<br>(3)<br>a. Preliminary analysis:<br>Pearson chi-square test,<br>Fisher's exact test (2-sided), Independent-<br>sample Student's t-test,<br>Kruskal-Wallis test,<br>Mann-Whitney U test<br>b. Main analysis: Binary<br>logistic regression<br>c. Controlled variables:<br>Age<br>(4) p < 0.05<br>(5)<br>a. Higher overall<br>mortality in F compared<br>to M<br>b. Only significant<br>mortality differences btw<br>M & F in age group of ≥<br>55 y/o<br>(6) NA | (1) F sex associated w/<br>higher risk of mortality<br>after Isol severe TBI,<br>especially in F ≥ 55 y/o<br>(2)<br>a. Pts true hormonal<br>status, hormone<br>replacement, or oral<br>contraceptive status<br>could not be determined<br>b. Mechanism of injury<br>was not accounted for in<br>assessment for blunt<br>mechanism<br>c. Smaller populations<br>from multiple age<br>stratifications<br>(3) Quality / risk of bias<br>(QUIPS): 9/11, + |
| (1) Ponsford (2008):<br>Injury, International<br>Journal of the Care of<br>the Injured [40]<br>(2) Australia,<br>Melbourne hospitals<br>(3) Longitudinal<br>(4) This study<br>examined pts w/ severe<br>TBI and hypotension to<br>compare 6-month<br>survival and functional<br>outcomes of M & F. | (1)<br>a. age ≥ 18 y/o<br>b. Severe TBI;<br>blunt head trauma<br>GCS < 9;<br>hypotension (SBP<br>< 100 mmHg); pts<br>w/ multisystem<br>trauma<br>c. NA<br>(2)<br>a. NR<br>b. Pts w/<br>penetrating<br>trauma; were<br>pregnant; had no<br>intravenous<br>access; serious<br>premorbid disease<br>identified on<br>medical bracelet;<br>peripheral<br>oedema; absent<br>sinus rhythm; or<br>cardiac arrest<br>c. Pts were in<br>close proximity to<br>the receiving<br>hospital | (1) Severe TBI 229<br>(100%)<br>(2) NR<br>(3) Traffic accident<br>circumstance: F: Driver<br>28 (35.9%), Passenger 19<br>(24.4%),<br>Motorcycle/bike 3<br>(3.8%), Pedestrian 26<br>(33.3%), Other 2 (2.6%)<br>M: Driver 56 (37.1%),<br>Passenger 22 (14.6%),<br>Motorcycle/bike 22<br>(14.6%), Pedestrian 35<br>(23.2%), Other 16<br>(10.6%)<br>(4) NR | (1) 229<br>(2) NR<br>(3) NR<br>(4)<br>a. Job Type: F: Skilled 7<br>(8.9%), Semiskilled 11<br>(14.1%), Unskilled 1<br>(1.3%), Student 8 (10.3%),<br>Retired 21 (26.9%), Other<br>30 (38.5%)<br>M: Skilled 27 (17.9%),<br>Semiskilled (male) 8<br>(5.3%), Unskilled 14<br>(9.3%), Student 17<br>(11.3%), Retired 18 (12%),<br>Other 67 (44.3%)<br>b. Change in Employment<br>Status: F: Same Job 1<br>(3.8%), Modified Job 3<br>(12%), Not working 22<br>(85%)<br>M: Same Job 12 (16%),<br>Modified Job 10 (14%),<br>Not working 51 (70%)<br>(5) 78 F, 151 M (65.9%)<br>(6) NR<br>(7) NR<br>(8) NR<br>(9) NR<br>(10)<br>a. Age: mean 43.3 ± 23.1<br>y/o (F); 33.8 ± 16.3 y/o<br>(M); range 16-89 y/o<br>b. Weight (kg): F: 63.5 ± | (1) Sex, age<br>(2) 6-month rate of<br>survival<br>(3)<br>a. Preliminary analysis:<br>Student's t-test, Mann-Whitney U test<br>b. Main analysis:<br>Multivariate analysis,<br>Logistic regression<br>c. Controlled variables:<br>Age, GCS, original study<br>group allocation (HTS or<br>control)<br>(4) p < 0.05<br>(5)<br>a. ↓ rate of 6-month<br>survival post-injury in F<br>b. F ≥ 60 y/o had a<br>significantly ↓ rate of<br>survival<br>(6)<br>a. M had ↑ likelihood to<br>be injured by motorcycle<br>or bike accidents<br>b. F more likely to be<br>injured as passenger or<br>pedestrian | (1) F w/ severe TBI and<br>hypotension associated<br>w/ worse survival<br>compared to age-<br>matched M<br>(2)<br>a. Severe TBI based on<br>GCS < 9 and prehospital<br>hypotension SBP < 100<br>mmHg; sample selected<br>more severe end of<br>severity<br>b. Age distribution and<br>causes of injury<br>differences btw sexes<br>(3) Quality / risk of bias<br>(QUIPS): 10/14, + |
|  |  |  | 11.5 ; M: 78.8 ± 15.8<br>c. Arrival of paramedics (min): F: 18.2 ± 13.7; M: 24.8 ± 50.1<br>d. Time to hospital (min): F: 70.4 ± 33.1 ; M: 77.7 ± 61.5<br>(11) Country-level indicators: a. GII score (publication year): 0.056 (2008) |  |  |
| (1) Selassie (2011): The Journal of TRAUMA Injury, Infection, and Critical Care [41]<br>(2) United States; South Carolina<br>(3) Cross-sectional<br>(4) The purpose of this study was to assess the risk factors of in-hospital death after acquiring TBI using time-to-event analyses and analyze the extent to which sepsis occurring in the hospital environment contributes to death. | (1)<br>a. aged ≥ 65 y/o<br>b. TBI ICD-9-CM 16 diagnosis codes 800 to 801, 803 to 804, 850 to 854, and 959.01<br>c. Death in an acute care facility w/ in 120 days after acquiring TBI<br>(2)<br>a. NR<br>b. Pts excluded coded as late effects of TBI (907.0) and repeat admissions for the same event.<br>c. NA | (1) All severities<br>a.<br>AIS: Severe AIS 4-6 16420 (39.7%), moderate AIS 3 7890 (19.1%), mild AIS 2 17085 (41.3%)<br>b.<br>ISS: Severe ISS 25-75 23955 (57.9%), moderate ISS 16-24 8059 (19.5%), mild ISS <16 9381 (22.7%)<br>(2) NR<br>(3) NR<br>(4) NR | (1) 41395<br>(2) Place of Residence: Rural 17545 (42.4%), Urban 23850 (57.6%)<br>(3) Race: Black 12319 (29.8%), White 29076 (70.2%)<br>(4) NR<br>(5) 14728 F; 26667 M (64.4%)<br>(6) NR<br>(7) NR<br>(8) NR<br>(9) NR<br>(10)<br>a.<br>Age: mean 43.7 ± 25.4 y/o<br>b. Insurance Status: Uninsured 6560 (15.6%), Indigent care/Medicaid 7905 (19.1%), Medicare 10576 (25.6%), Commercial 16354 (39.5%)<br>c.<br>Comorbid Conditions: Elixhauser's list- Liver-renal 431 (1.1) 70 (16.2) 361 (83.8), Neurological-stroke 1,217 (2.9) 227 (18.7) 990 (81.4) , Diabetes-metabolic 4,291 (10.4) 626 (14.6) 3,665 (85.4), Heart diseases 4,394 (10.6) 707 (16.1) 3,687 (83.9), Other comorbid diseases 10,743 (25.9) 687 (6.4) 10,056 (93.6) , No comorbid disease 20,319 (49.1)<br>(11) Country-level indicators: a. GII score (publication year): 0.169 (2011) | (1) Sex, Age, Race, Insurance Status, Place of Residence<br>(2) In-hospital mortality<br>(3)<br>a. Preliminary analysis: t-tests, Chi-square test, Fisher's exact tests, Kaplan-Meier survival curves, Log-Rank test<br>b. Main analysis: Cox proportional hazards regression<br>c. Controlled variables: Age, Sex, injury severity<br>(4) p < 0.05<br>(5) F ↓ in mortality risk than M<br>(6)<br>a. Age significant ↑ in-hospital mortality risk<br>b. Race: White ↑ mortality risk compared to Black<br>c. Insurance Status: Uninsured significant ↑ in-hospital mortality risk; Indigent care/Medicaid significant ↓ in-hospital mortality risk; Medicare ↑ in-hospital mortality risk<br>d. Place of Residence: Rural ↑ in-hospital mortality risk compared to Urban | (1) TBI severity is the strongest risk factor for death; pts ≥ 65 y/o and those with comorbid conditions at higher risk of mortality<br>(2)<br>a. Case ascertainment relied on administrative/billing data<br>b. Variable skillset & diagnostic resources across hospitals may affect accuracy of sepsis and TBI diagnosis codes<br>c. AIS/ISS derived from ICD-9-CM codes (accuracy coding-dependent); GCS unavailable for all patients<br>d. Residual confounders unavailable in dataset<br>(3) Quality / risk of bias (QUIPS): 8/11, + |
| (1) Selassie (2014): Journal of Head Trauma Rehabilitation [26]<br>(2) United States, California, Acute care hospital in South California<br>(3) Longitudinal<br>(4) This study examined the effects of preexisting heart, liver, kidney, cancer, stroke, and mental health problems and low socioeconomic status on mortality for pts w/ TBI after discharge from acute care facilities. | (1)<br>a. NR<br>b. mTBI (AIS 2); moderate TBI (AIS 3); severe TBI (AIS 4-6)<br>ICD-9-CM diagnosis codes 800-801, 803-804, 850-854, and 959.01; pts w/ multiple adms for repetitive TBI had the most severe case selected<br>c. NA<br>(2)<br>a. NR<br>b. Pts w/ ICD-9-CM codes 907.0 for late effects of intracranial injury & repeat encounters w/ the same diagnosis<br>c. NA | (1) All severities<br>Injury severity: AIS 2 15400 (45.7%); AIS 3 6614 (19.6%); AIS 4-6 11681 (34.7%)<br>(2) NR<br>(3) NR<br>(4)<br>Cause of injury: Fall 9488 (28.2%); Acts of violence 2596 (7.7%); Motor vehicle/transport 16406 (48.7%); All other causes 4413 (13.1%); Struck by (intentional) 792 (2.4%) | (1) 33695<br>(2) NR<br>(3) Race: Non-Hispanic white 23343 (69.3%); Non-Hispanic black 8498 (25.2%); Other 1854 (5.5%)<br>(4) NR<br>(5) 12108 F, 21587 M (64.1%)<br>(6) NR<br>(7) NR<br>(8) NR<br>(9) NR<br>(10)<br>a.<br>Age: mean $42.8 \pm 25.3$ y/o<br>b. Insurance Status: Uninsured 5387 (16%), Indigent care/Medicaid 6804 (20.2%), Medicare 8291 (24.6%); Commercial 13210 (39.2%)<br>c.<br>Comorbid Conditions: Heart diseases 3307 (9.8%), Liver-renal diseases 326 (1.0%), Cancer 422 (1.3%), Hypertensions 4992 (14.8%), Diabetes and metabolic illnesses 3236 (9.6%), Neurological diseases and stroke 859 (2.6%), Mental health problems 2032 (6.0%), All other comorbid conditions 1540 (4.6%), No comorbid condition 16981 (50.4)<br>(11) Country-level indicators: a. GII score (publication year): 0.169 (2014) | (1) Sex, age<br>(2) All-cause mortality post-discharge<br>(3)<br>a. Preliminary analysis: Chi-square test, t-test<br>b. Main analysis: Cox proportional hazards regression, log-log Kaplan-Meier plots, Kaplan-Meier survival curves, Log-rank test<br>c. Controlled variables: Age, Sex, Race<br>(4) $p < 0.05$<br>(5) F associated w/ a ↓ risk of death compared to M<br>(6)<br>a. Risk of death ↑ by 5% for each year of age<br>b. Pts w/ indigent insurance and Medicare insurance had ↑ risk of death compared to those w/ private insurance<br>c. High risk of death associated w/ pts w/ liver-renal diseases, cancer, heart diseases, stroke and neurological disorders<br>d. Diabetes. Mental health problems, hypertension, and diseases listed as all other had moderately strong associated risks of death | (1) Preexisting diseases, low socioeconomic status, age, sex, and injury severity associated with elevated risks of mortality after TBI<br>(2)<br>a. Coding errors and duplicates observations possible in administrative data systems<br>b. Exclusion of comorbid conditions that may not be helpful to maximize reimbursement<br>c. Pts diagnosed w/ TBI who died out of state after discharge not included by system<br>(3) Quality / risk of bias (QUIPS): 8/14, + |
| (1) Shandro (2008): The Journal of TRAUMA Injury, Infection, and Critical Care [27]<br>(2) United States<br>(3) Longitudinal<br>(4) The purpose of this study is to use data from the National Study on Costs and Outcome from Trauma (NSCOT) to examine the association of alcohol with outcome from TBI. | (1)<br>a. aged $\geq 18$ to 84y/o<br>b. ICD-9-CM codes 800-959 (excluding burns); a head injury of AIS $> 3$, regardless of other injuries<br>c. NA<br>(2)<br>a. Pts that spoke neither English nor Spanish, or were non-U.S. residents were excluded<br>b. Pts presented with no vital signs and were pronounced dead within 30 minutes of arrival, delayed seeking treatment for more than 24 hours, were 65 years of age or older with a first listed diagnosis of hip | (1) Moderate and severe TBI<br>AIS: AIS 4 820 (53.63%); AIS 5-6 709 (46.37%)<br>Head<br>AIS: Head AIS 4 842 (55.07%); Head AIS 5-6 687 (44.93%)<br>NISS: $37.3 \pm 21.0$ (BAC 0); $41.4 \pm 20.1$ (BAC 1-100); $38.8 \pm 19.8$ (BAC 101-230); $40.7 \pm 24.0$ (BAC $> 230$)<br>GCS (first ED motor score): 6: 634 (41.46%); 4-5: 220 (14.39%); 2-3: 79 (5.17%); 1, not paralyzed: 291 (19.03%); 1, paralyzed: 305 (19.95%)<br>Midline shift on CT: Yes 495 (32.37%); No 1035 (67.69%)<br>Shock: ED shock 86 (5.62%); Prehospital shock 202 (13.21%)<br>(2) NR<br>(3) NR<br>(4) Injury Type: Blunt 1371 (89.67%); Penetrating 158 | (1) 1529<br>(2) NR<br>(3) Race: Hispanic 252 (16.48%); Non-Hispanic, white 995 (65.08%); Non-Hispanic, non-white 283 (18.51%)<br>(4) NR<br>(5) 456 F, 1073 M (70.18%)<br>(6) NR<br>(7) NR<br>(8) NR<br>(9) NR<br>(10)<br>a.<br>Age: Mean: $47.8 \pm 32.3$ y/o (BAC 0), $37.8 \pm 26.6$ y/o (BAC 1-100), $40.3 \pm 28.7$ y/o (BAC 101-230), $44.8 \pm 25.4$ y/o (BAC $> 230$); Range: 18-84 y/o<br>b. Insurance Status: No insurance 312 (20.41%), Private + Medicare 339 (22.17%), Private only 213 (13.93%), Medicare only 442 (28.91%), Medicaid 178 (11.64%), Other 45 (2.94%)<br>c. Comorbidities (Charlson Index): 0: 973 (63.64%); 1: | (1) Sex, Age, Race, Insurance<br>(2) In-hospital, 90-day, and 365-day mortality<br>(3)<br>a. Preliminary analysis: Comparison of patient characteristics across BAC categories (unadjusted); multiple imputation (10 datasets) for missing BAC values<br>b. Main analysis: Multivariate Weighted Logistic Regression<br>c. Controlled variables: Age, Sex, New ISS, Insurance status, injury mechanism, midline shift, ED shock, GCS motor score, Comorbidities, Maximum AIS, Head maximum AIS<br>(4) $p < 0.05$<br>(5) F ↓ in mortality risk than M<br>(6)<br>a. Age significant ↑ in-hospital mortality risk<br>b. Race: Hispanic ↑ mortality risk compared | (1) No significant difference in in-hospital, 90-day, or 365-day mortality by BAC after adjustment for injury severity and confounders; Nonsignificant trend toward lower mortality at higher BAC levels, warranting further investigation<br>(2)<br>a. Starting time bias: BAC drawn in ED, not at time of injury (74% came directly to study hospitals; referring-hospital BAC available for 36% of transfers)<br>b. Limited to acute alcohol intoxication; no data on preinjury alcohol dependence<br>(3) Quality / risk of bias (QUIPS): 7/14, + |
|  | fracture, had major burns, c. . Incarcerated and homeless patients were excluded | (10.33%) | 220 (14.39%); 2: 136 (8.90%); ≥3: 200 (13.08%)<br>(11) Country-level indicators: a. GII score (publication year): 0.169 (2008) | to Non-Hispanic white and black pts<br>c. Insurance Status: Private + Medicare, Private only, and Medicaid significant ↑ in-hospital mortality risk; Other significant ↓ in-hospital mortality risk; Medicare ↑ in-hospital mortality risk |  |
| (1) Shibahashi (2017): World Neurosurgery [42]<br>(2) Japan<br>(3) Longitudinal<br>(4) In this study, we aimed to investigate the incidence of, and risk factors for, in-hospital mortality of patients who talked after TBI. | (1)<br>a. aged ≥ 16 y/o<br>b. TBI was defined as AIS 3-5 for the head. Talking after an injury was defined as a score of ≥3 on the verbal component of the GCS on adm<br>c. NA<br>(2)<br>a. aged < 16 y/o<br>b. SBP <40 mmHg, and 3) presence of severe injury (AIS score ≥3) on body regions other than the head<br>c. NA | (1) Severe TBI<br>ISS: 16 (survived), 20 (death)<br>GCS: 14 (survived), 13 (death)<br>(2) NR<br>(3) NR<br>(4) Causes of Injury: Accident (survivors), 22637, 95<br>Suicide (survivors) 245, 1<br>Others (survivors), 175, 1<br>Unknown (survivors), 820, 3<br>Accident (Deaths), 891, 93<br>Suicide (Deaths), 8, 1<br>Others (Deaths), 11, 1<br>Unknown (Deaths), 46, 5 | (1) 24833<br>(2) NR<br>(3) NR<br>(4) NR<br>(5) Survivors: 7928 F, 15949 M (67%)<br>Deaths: 274F, 682 M (71%)<br>(6) NR<br>(7) NR<br>(8) NR<br>(9) NR<br>(10) a.<br>Age: mean (range): Survivors: 66 (48-78) y/o, Deaths: 77 (67-84) y/o<br>(11) Country-level indicators: a. GII score (publication year): 0.059 (2017) | (1) Sex, Age<br>(2) In-hospital mortality<br>(3) a. Preliminary analysis: Chi-square test, Fisher's exact tests, Mann-Whitney U test<br>b. Main analysis: Multivariable Logistics Regression<br>c. Controlled variables: Age, Sex, Year of admittance, GCS score on arrival, comorbidities, hypotension (<90 mm Hg systolic) on arrival, RTS, ISS, whether head CT was performed for initial surveying, and the nature of the head injury<br>(4) p < 0.05<br>(5) F significant ↓ in mortality risk than M<br>(6) Age significant ↑ in-hospital mortality risk | (1) Worse outcomes in M considered biologically plausible (possible neuroprotective role of gonadal steroids); mechanism not addressed<br>(2) a. Retrospective design prone to bias; JTDB lacked detailed CT findings (e.g., midline shift) and blood test results (e.g., coagulopathy, glucose, hemoglobin)<br>b. Patients older than in prior studies (aging Japanese population); "talk and die" incidence may be lower in younger patients; results warrant external validation<br>(3) Quality / risk of bias (QUIPS): 9/14, + |

\*Parameters considered in analysis of sex diff; † Study uses the terms men and women (gender) but is referring to sex; ‡ Study indicated the recruitment took place in the local population. Abbreviations: Adm, admission; AIS, abbreviated injury scale score; BAC, blood alcohol content; btw, between; COPD, chronic obstructive pulmonary disease; diff, difference; ED, Emergency department; F, female; GCS, Glasgow Coma Scale; HBI, hemorrhagic brain injury; HR, hazard ratio; hrs, hours; ICD-9-CM, International Classification of Disease version 9

Studies were conducted across multiple countries, including the United States (n = 12) [19, 21, 22, 23, 24, 25, 29, 30, 31, 32, 36, 38], Canada (n = 1) [19], Australia (n = 2) [26, 36], South Korea [32], Japan [33, 37], Singapore [11], the Netherlands [27], and multiple European centers [34]. Overall, studies were conducted primarily in North America (n = 13), followed by Asia (n = 4), Europe (n = 2), and Oceania (n = 2).

Clinical Modification; ICD-10, International Classification of Disease version 10; ICP, Intracranial pressure; ICU, intensive care unit; Isol, isolated; ISS, injury severity score; IQR: Interquartile range; LOS, length of stay; M, male; MCC, motorcycle collision; mTBI, mild traumatic brain injury; MV, mechanical ventilation; MVA, MVC, motor vehicle collision; NA, not applicable; NR, not reported; OSHPD, Office of Statewide Health Planning and Development; RBC, red blood cell; Pts, patients; RTS, Revised Trauma Score; SBP, systolic blood pressure; SI, Shock Index; SNF, skilled nursing facilities; TBI, traumatic brain injury; w/, with; w/in, within; y/o, years old.

Age eligibility criteria investigated varied across study samples, consistent with our predefined inclusion criteria of participants aged ≥18 years or study samples with a reported mean age >18 years. Seven studies enrolled participants aged ≥16 years [20, 21, 24, 26, 27, 36, 37], and three allowed enrollment from age 13 years [25, 28, 32]; all were retained because mean participant age exceeded 18 years. Four studies included populations aged ≥50 years [18, 22, 30, 31], and seven studies did not explicitly apply age restrictions, but reported mean or median ages of participants older than 18 years [11, 19, 23, 29, 33, 34, 35].

Fig 3 summarizes the reporting of study characteristics and PROGRESS-Plus variables across included studies, organized by mortality ascertainment period. The median sample size was 4,854 (IQR, 1,250 - 30,368) and was used as the threshold to classify studies as having smaller or larger sample sizes.

**Fig 3.**
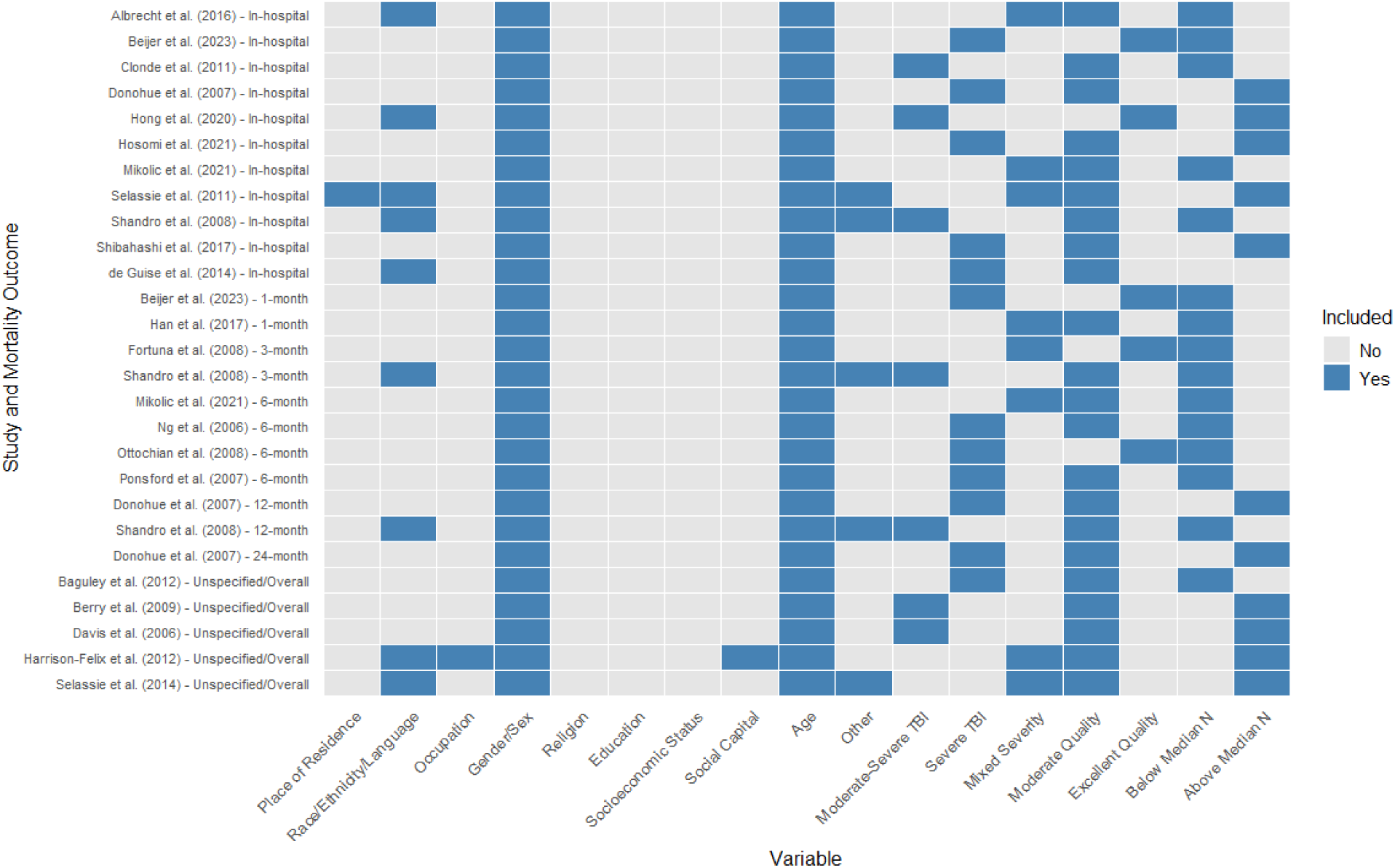
Heatmap of study characteristics and PROGRESS-Plus parameters included across 21 studies (27 cohorts) assessed. Blue cells indicate that the variable was reported or considered in the analyses in the study, while grey cells indicate that the variable was not reported. Studies are organized by mortality outcomes, including in-hospital, 1-month, 3-month, 6-month, 12-month, 24-month, and unspecified/overall mortality. The median sample size was 4854 (IQR, 1,250 - 30,368) with studies classified as below or above the median sample size. Plus parameters (Other) included insurance status.

Nine studies (42.86%) [23, 24, 25, 26, 34, 35, 36, 37, 41] included participants with mixed severity TBI, five (23.81%) [27, 28, 31, 32, 38] included participants with moderate to severe TBI, and seven (33.33%) [11, 29, 30, 33, 39, 40, 42] included participants with severe TBI. Although some mixed-severity cohorts included participants with mild TBI, mild TBI was not analyzed as a separate ML stratum because insufficient study-level data were available.

PROGRESS-Plus social parameters were considered in analyses in a total of seven studies (33.33%) [23, 24, 25, 26, 27, 38, 41]: seven studies reported on race/ethnicity or language, one each (4.76%) on place of residence/recruitment location [41], occupation [25], and social capital [25]. Plus, parameters, which only included insurance status, were considered in three studies (14.29%) [26, 27, 41]. Detailed findings are presented in the S5 Appendix. Overall, PROGRESS-Plus factors were operationalized using heterogeneous definitions, reference groups, and effect measures, and no studies presented these associations stratified by sex, limiting direct comparison and precluding quantitative synthesis of sex-specific social determinants of mortality.

Mortality types varied across included studies (Fig 3). Eleven studies [18, 19, 21, 22, 24, 25, 27, 31, 33, 34, 37] reported in-hospital mortality. Mortality at one, three, six, 12, and 24 months was reported in two [27, 32], two [24, 31], four [11, 34, 35, 36], two [24, 30], one study [30], respectively. Five studies reported overall or unspecified mortality [20, 23, 26, 28, 29] (Fig 3).

### Risk bias in included studies

Four studies (19%) were rated as high quality [11, 34, 38, 39 and the remaining 17 studies [18, 19, 21, 22, 24, 25, 27, 31, 33, 34, 37] (81%) were rated as moderate quality. No studies were rated as low quality. Methodological limitations were concentrated in participant selection, sample size, statistical power, and adjustment for potential confounders, indicating potential risks of selection bias, imprecision, and limited control of confounding. Detailed quality-appraisal findings are presented in S1 Table 2.

### Meta-analysis

Sixteen studies [11, 23, 24, 27, 28, 30, 31, 32, 33, 34, 36, 37, 38, 39, 40, 42] (76.2%) reported adjusted ORs, four [26, 29, 35, 41] (19.0%) reported HRs, and one each (4.8%) reported SMRs [25] and RRs [25] comparing mortality between females and males following TBI.

Random-effects meta-analysis of pooled ORs showed significantly lower odds of TBI mortality in females than males (OR = 0.88, 95% CI: 0.80-0.98, *p* = 0.017) (Fig 4a). When stratified by mortality ascertainment period, for in-hospital, one-month, six-month, 12-month, and unspecified/overall mortality, significant results were driven by lower odds of in-hospital mortality (OR = 0.86, 95% CI: 0.76-0.97, *I^2^* = 90.8%, *p* = 0.015) and unspecified/overall mortality (OR = 0.83, 95% CI: 0.78-0.88, *I^2^*= 0%, *p* <0.001) (Fig 4a). No significant associations were found for mortality at six months (OR = 1.15, 95% CI: 0.67-1.97, I^2^ = 82.2%, *p* = 0.618). Egger’s test showed no evidence of funnel-plot asymmetry (*p =* 0.383; S2 Fig 1).

**Fig 4.**
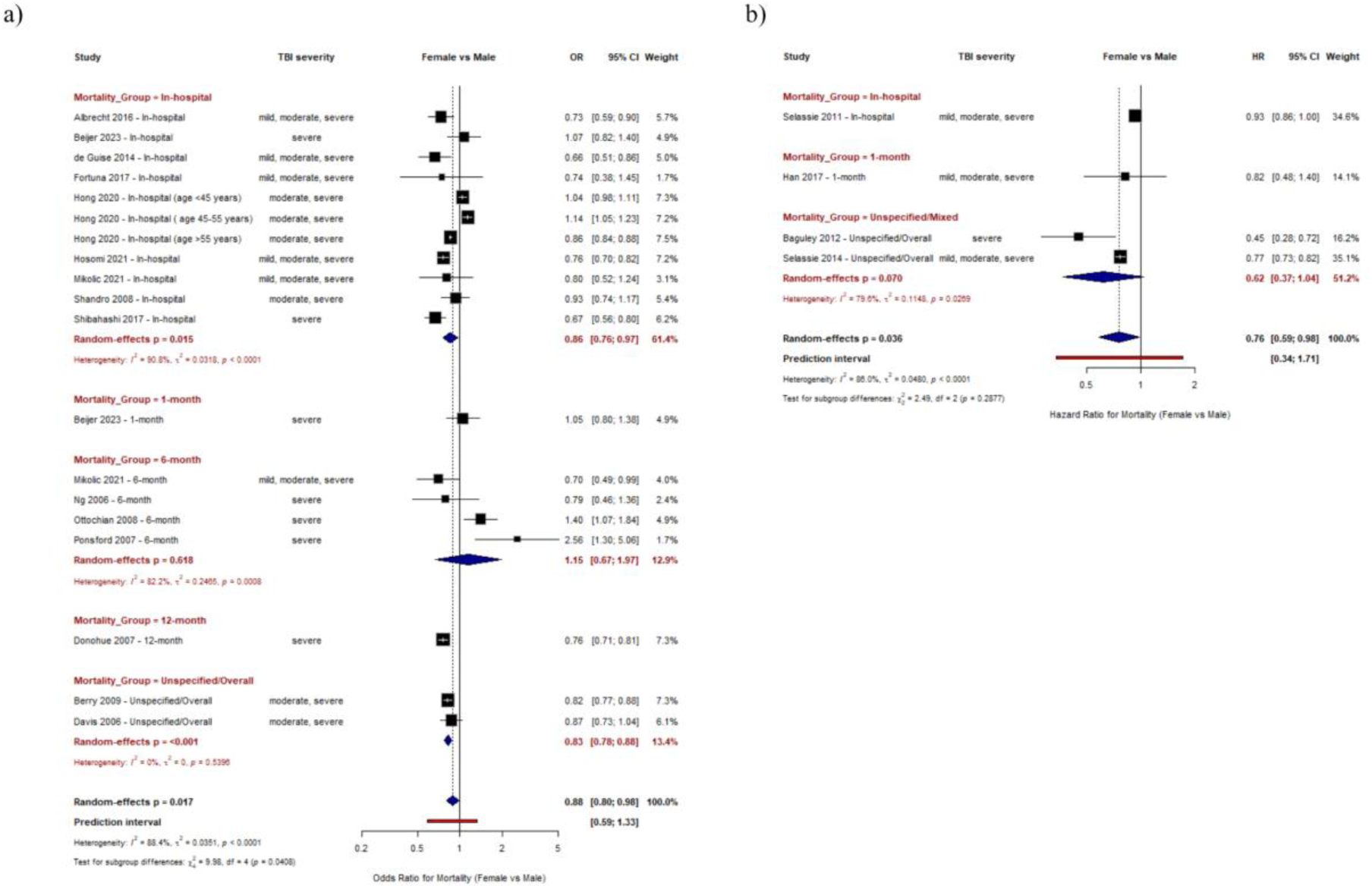
Forest plot of random effects meta-analysis of individual studies and pooled results of adjusted odds ratios (a) and hazard ratios (b) for mortality after TBI by mortality ascertainment period, comparing females versus males.

Results of pooled HR were statistically significant and supported the direction of associations (HR = 0.76, 95% CI: 0.59-0.98, *p* = 0.036). In subgroup analysis, the unspecified/mixed mortality subgroup suggested females having lower mortality risk compared with males, however, this was not statistically significant (HR = 0.62, 95% CI: 0.37-1.04, I^2^ = 79.6%, *p* = 0.070) (Fig 4b). Egger’s test showed no evidence of funnel-plot asymmetry (*p =* 0.214; S2 Fig 2).

Heterogeneity was substantial for both the overall pooled OR (*I²* = 88.4.0%, τ² = 0.0351, *p* < 0.0001) and HR (*I²* = 86.0%, τ² = 0.0480, *p* < 0.0001) results. The test for subgroup differences by mortality ascertainment period were statistically significant (χ² = 9.98, df = 4, p = 0.0408) in the OR meta-analysis but not the HR meta-analysis (χ² = 2.49, df = 2, p = 0.2877) (Figs 4a, b).

Clond et al. [28] reported sex-stratified adjusted ORs for mortality predictors. Increasing age per year was associated with mortality in females (OR = 1.04, 95% CI: 1.02-1.05, *p* = <0.0001) and males (OR = 1.04, 95% CI: 1.03-1.05, *p* <0.0001). For Injury Severity Scores (ISS) ≥ 16, the association with mortality was stronger in males (OR = 23.58, 95% CI: 7.42-74.97, *p =* <0.0001) compared to females (OR = 4.51, 95% CI:1.33-15.31, *p = 0*.016); and for Glasgow Coma Scales scores (GCS) ≤ 8, effect estimates were elevated in males (OR 8.32, 95% CI 6.01-11.50) and females (OR 9.74, 95% CI 5.32-17.84). Harrison-Felix et al. [25] examined overall mortality in a mixed-severity TBI cohort. Females had a significantly lower mortality risk than males (RR 0.59, 95% CI 0.48-0.72). Standardized mortality ratios were elevated in both sexes but were higher in males (SMR 2.31, 95% CI 2.13-2.49) than females (SMR 2.08, 95% CI 1.79-2.36).

### Meta-analysis sensitivity analyses

Sensitivity analysis by injury severity results were significant for mixed injury severity only, which showed that females were associated with lower mortality odds (OR = 0.75, 95% CI: 0.79-0.80, *I^2^* = 0%, *p* < 0.001). No significant associations were found in the moderate-severe (OR = 0.94, 95% CI: 0.84-1.05, *p* = 0.274) or severe TBI groups (OR = 1.01, 95% CI: 0.77-1.33, *p* = 0.944) (Fig 5). The test for subgroup differences was statistically significant (χ² = 14.26, df = 2, *p* = 0.0008).

**Fig 5.**
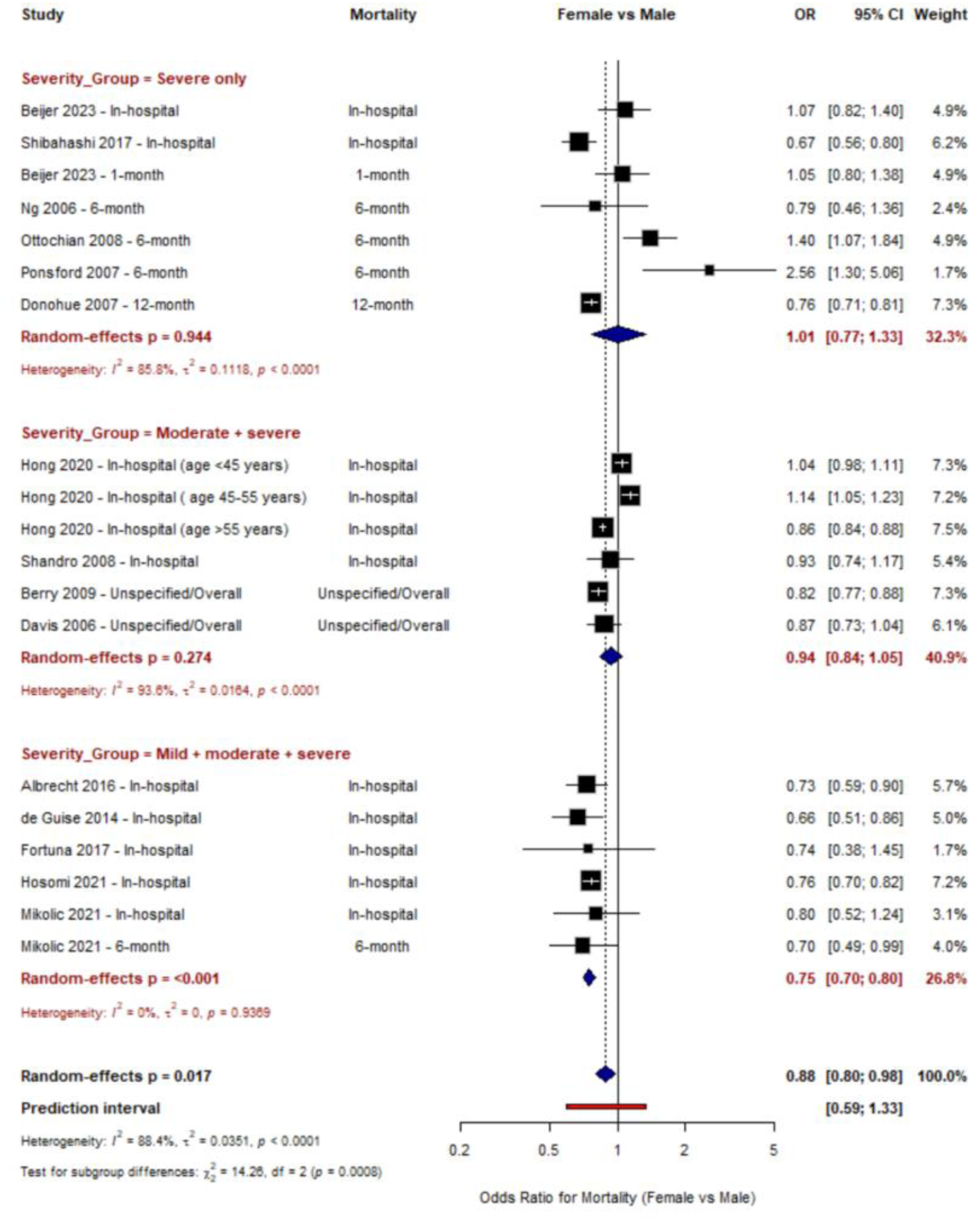
Forest plot of random effects meta-analysis of individual studies and pooled results of adjusted odds ratios for mortality after TBI by injury severity, comparing females versus males.

In the OR meta-analysis stratified by study quality, moderate quality studies were associated with significantly lower odds of mortality for females compared to males (OR = 0.78, 95% CI: 0.74-0.82, *p* < 0.001), but not among high-quality studies (OR = 1.04, 95% CI: 0.92-1.18, *p* = 0.510). The test for subgroup differences was not statistically significant (χ² = 14.26, df = 2, *p* = 0.1186) (S2 Fig 3a). All the studies for the HR analysis were of moderate quality (S2 Fig 3b).

Stratification by median sample size showed no significant sex difference in mortality among studies below the median (OR 1.01, 95% CI 0.83-1.22; p = 0.944). Studies above the median showed lower odds of mortality in females than males (OR 0.84, 95% CI 0.75-0.94; p = 0.004). The test for subgroup differences was not statistically significant (χ² = 5.10, df = 2, *p* = 0.078) (S2 Fig 4A).

For adjusted HRs, neither studies below the median sample size (HR = 0.60, 95% CI: 0.33-1.08, *p* = 0.088), nor above the median (HR 0.84, 95% CI 0.70-1.02; p = 0.074) showed a statistically significant sex difference in mortality, although both estimates favored lower mortality hazards in females. The test for subgroup differences was not statistically significant (χ² = 1.19, df = 1, *p* = 0.2753) (S2 Fig 4B).

### Exploratory explainable ML analysis

#### Data preprocessing

The complete list of extracted variables and their data coverage is provided in Fig 3. The availability of study-level moderators varied substantially across included studies. Variables with the greatest proportion of missing data were race/ethnicity/language (91.9%), occupation (96.5%), and mean age of the sample (75.5%), education, socioeconomic status, and social capital (75.5-100% missing); they were excluded from ML modelling which require complete feature matrices.

We created heatmaps with correlation coefficient matrices for moderate-severe and severe TBI to identify associations among potential modifiers in mortality (Fig 6). We included mean ages of samples, sex and gender ratios of males, and study quality, and GII as contextual measures and effect modifiers based on the country of study origin as primary predictors. The assigned GII values ranged from 0.013 to 0.226, and the country-level structural indicators contributed to measurable heterogeneity across moderate-severe and severe TBI cohorts in several key predictors. For moderate-severe TBI, the proportion of male participants had the strongest positive correlation with mortality effect estimates (r = 0.42), followed by study quality (r = 0.12) and GII (r = 0.11) which showed weak positive correlations (Fig 6a). For severe TBI, study quality showed the strongest positive correlation with mortality effect estimates (r = 0.35), followed by GII (r = 0.29) and mean age (r = 0.23). The proportion of male participants was negatively correlated with mortality effect estimates (r = - 0.24) (Fig 6b).

**Fig 6.**
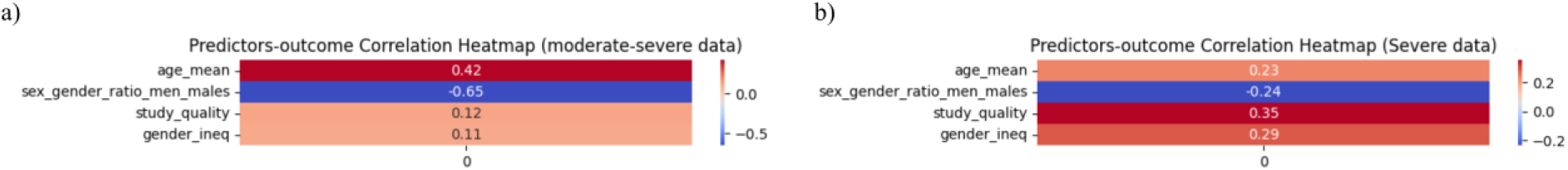
Heatmaps of the correlation coefficient matrix, by injury severity (a, moderate-severe TBI; b, severe TBI). Heatmaps of the correlation coefficient matrix for each of the parameters’ association with the outcome, mortality (bivariate associations). Positive correlations are represented as red, while negative correlations are indicated as blue. Colour intensity signify the correlation coefficient magnitude, where stronger correlations are represented as darker shades; shades of red represent coefficients approaching 1 and blue shades represent coefficients approaching − 1.

#### Machine learning modeling

Figs 7 and 8 present the results of three supervised ML algorithms, Random Forest (RF), Gradient Boosting (GB), and extreme GB (XGBoost), including the importance of mortality effect modifiers, for moderate-severe and severe TBI, respectively.

**Fig 7.**
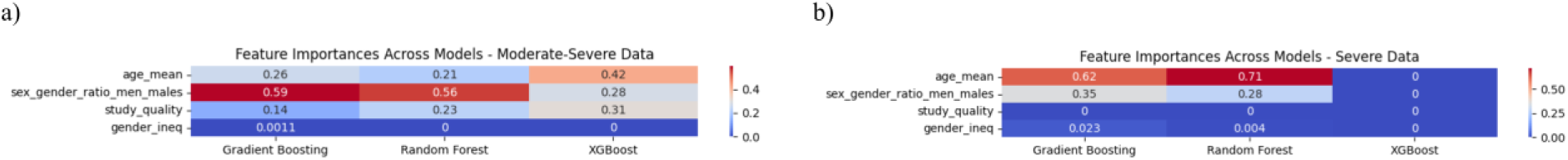
Feature contributions across machine-learning models (a, moderate-severe TBI; b, severe TBI). Ranking of feature contributions for prediction of mortality in Random Forest, Gradient Boosting, and XGBoost models. The strength of positive association is represented by the colour scale, where darker red signifies the strongest positive correlations and darker blue signfiies the weakest positive correlations. The magnitude of the correlation coefficient is represented by colour intensity and gradients in effect size are shown as intermediate shades.

**Fig 8.**
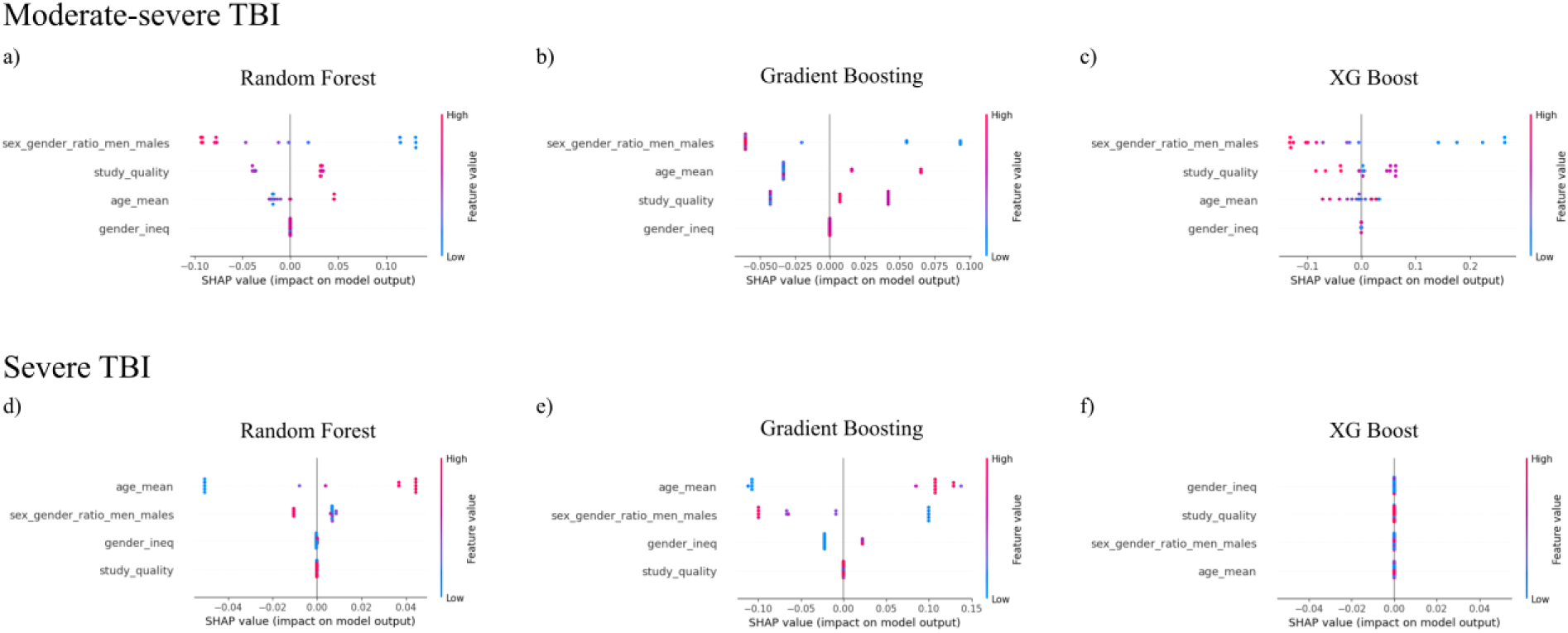
SHAP summary of feature impact on predictive models’ output (a-c moderate-severe TBI; d-f severe TBI). SHAP summary of feature impact on predictive models’ output (a-c mild TBI; d-f moderate-severe TBI). SHAP summary plots showing the influence of input variables on model predictions for Random Forest (a, d), Gradient Boosting (b, e), and XGBoost (c, f) models. The colour of each point indicates the value of the feature, with red corresponding to higher values and blue to lower values. The position along the x-axis reflects whether the feature increases or decreases the prediction in the model output. Features are ranked on the y-axis based on their overall importance, with those contributing most to the model’s predictions appearing at the top.

Feature importance heatmaps of each of the models for moderate-severe and severe TBI, are shown in Figs 7a and b. For moderate-severe TBI, across the three models, the top feature was sex ratio of males (GB 0.59, RF 0.56, XGBoost 0.28), followed by mean age (GB 0.26, RF 0.21, XGBoost 0.42), study quality (GB 0.14, RF 0.23, XGBoost 0.31), and GII (GB <0.01, RF 0, XGBoost 0). For severe TBI cohorts, the top feature was mean age (GB 0.62, RF 0.71, XGBoost 0), followed by sex ratio of males (GB 0.35, RF 0.28, XGBoost 0), GII (GB 0.02, RF< 0.01, XGBoost 0) and study quality (GB 0, RF 0, XGBoost 0).

#### Model evaluation and interpretation

Predictions were displayed using the SHAP rankings. For moderate-severe TBI cohorts, the sex ratio of males emerged as the strongest predictor across all models, followed by study quality in RF and XGBoost, and mean age in GB (Fig 8). GII had the weakest importance amongst the predictors in all models (Fig 8). For severe TBI, mean age emerged as the strongest predictors in RF and GB, followed by the sex ratio of males and GII (Fig 8). The XGBoost model generated zero importance for all predictors in the severe TBI cohort (Fig 8).

Predictive accuracy was comparable across ML algorithms but varied by injury severity.

Gradient Boosting showed consistent MAE and RMSE across severity levels, with Random Forest and XGBoost having slightly lower MAE and RMSE than Gradient Boosting in severe TBI cohorts. Model performance metrics are summarized in S3 Table.

#### Explanatory ML sensitivity analyses

Three sensitivity analyses were conducted by adding modifiers to the main ML models. First, we added mean age SD (S4 Figs 4 and 5). The sex ratio of males remained as the strongest importance parameter, followed by age SD, study quality, mean age, and GII in the RF model for moderate-severe TBI cohorts. Gradient Boosting and XGBoost models revealed age SD as the predictor of strongest importance followed by the sex ratio of males, reducing the value of study quality and mean age in SHAP analysis. In the severe TBI cohort, GII led as the strongest predictor in the RF and XGBoost models, followed by sex ratio of males and study quality, and study quality and sex ratio of males, respectively. Gradient Boosting showed age SD as the strongest importance, followed by mean age, study quality, and GII, and reduced the value of sex ratio of males (S4 Figs 4 and 5).

Second, we added region/country of study origin to the main models, which showed minimal effect on the importance of parameters in the main analysis among the moderate-severe cohort. For severe TBI cohorts, mean age and sex ratio of males remained as strong predictors in RF, while age SD and mean age were of importance in GB (S4 Figs 6 and 7).

Third, we added sample cohort size to the main models. For moderate-severe TBI, we observed that sex ratio of males remained as the strongest predictor, followed by mean age across models. In GB, sample cohort size weakened the importance of mean age and GII. In severe TBI cohorts, GII emerged as the strongest predictor across all models (S4 Figs 8 and 9).

## Discussion

In this systematic review and meta-analysis of 21 studies, encompassing nearly 900,000 participants with TBI and more than 98,000 fatal events, mortality was lower among females than males overall, with the association most evident for in-hospital and overall or unspecified mortality. However, substantial variation in the magnitude and direction of associations was observed across studies, mortality ascertainment periods, and clinical settings, indicating that sex should not be interpreted as a uniform predictor of mortality across TBI populations. Explainable ML analyses identified cohort age, sex composition, and study quality as important characteristics associated with variation in study-level effect estimates. These findings highlight the importance of population and study context when interpreting sex-related prognostic associations and developing data-driven models for TBI. From a digital health perspective, the observed heterogeneity has implications for the development and validation of prognostic models and clinical decision-support tools. Models that incorporate sex-related associations without accounting for differences in population composition, age distribution, and clinical context may have variable performance when applied across settings [20, 43, 44]. The identification of study-level characteristics through explainable ML provides a complementary, hypothesis-generating approach to conventional meta-analysis by highlighting characteristics that may contribute to variation in reported associations [45]. However, these findings should not be interpreted as evidence that the identified characteristics causally modify mortality risk.

Rather, they underscore the need for transparent reporting, external validation, and evaluation of model performance across diverse TBI populations before deployment in clinical digital health settings [46, 47]. Previous biomedical, clinical, and population-based studies reporting lower mortality in females after TBI [16, 48] have often been attributed to the neuroprotective effects of oestrogen and progesterone [14]. Experimental evidence supports biological plausibility for this hypothesis [49, 50, 51, 52]; however, if circulating sex hormones are major determinants of survival after TBI, one might expect the magnitude of sex differences to vary across age groups corresponding to major hormonal transitions, including puberty and menopause. Instead, sex specific mortality estimates remained heterogeneous even among studies with broadly comparable age and injury severity profiles (Table 1).

Importantly, mean age also emerged as one of the most important study-level characteristics in our explanatory ML analyses, indicating that the age composition of cohorts contributed meaningfully to variation in reported sex-specific mortality effects. This finding should be interpreted as evidence that suggests that age may capture a constellation of biological, clinical, and social factors, including hormonal profile, comorbidity, circumstances of injury, and quality care, that differ across samples included in this review [14, 16, 48], and may shape reported differences in mortality after TBI among males and females. This result is clinically important because age may modify sex differences in mortality through several pathways, including age-related frailty and comorbidity, different mechanisms of injury, differences in care timeliness and intensity, and changes in endogenous sex-hormone exposure across the lifespan [14, 16, 48]. Our ML findings should therefore be interpreted as hypothesis-generating: they identify characteristics that distinguish between-study differences in sex-related mortality effects. However, an important consideration for future research is that studies included in this review used binary terms labelled as sex (e.g., male/female) and gender (e.g., men/women) interchangeably, without distinguishing between their biological and social dimensions.

Contemporary research, including Sex and Gender Equity in Research (SAGER) guidelines [53], conceptualizes sex and gender as distinct but interconnected constructs: sex refers to biological attributes of people, and gender encompasses socially constructed roles, behaviours, relationships, and community context, that may influence injury exposure, care seeking, treatment, and survival. Because the included studies operationalized these constructs as binary variables, their analyses could not disentangle biological from social influences.

Accordingly, observed mortality differences between males and females should not be interpreted as biological sex differences alone [5, 54]. Rather, these estimates likely reflect sex- and gender-related biological, social, behavioural, and structural factors, which may also influence research participation, study sample composition, and outcome trajectories among individuals with TBI. This distinction is particularly important because sex and gender may influence outcomes through different mechanisms and may also shape the probability of research participation among people with TBI, thereby influencing the composition of study samples and the outcome trajectories observed [55, 56].

Evidence also suggests that females with TBI may be less likely than males to access specialized trauma care, and sex- and gender-driven differences have also been observed in post-acute discharge and rehabilitation pathways [16, 57]. Most studies included in this review were conducted in healthcare-linked research settings, and as such, the observed female-male mortality differences may also be influenced by who reaches, accesses, and is represented within these settings [16, 57]. Differential care pathways, including referral to specialized trauma services, and inclusion in healthcare-based research settings, could alter the composition of study samples, and potentially bias in mortality comparisons. Future research should therefore extend beyond healthcare-linked cohorts, to better determine whether observed differences in mortality reflect biological vulnerability, differential healthcare exposure, bias in research selection processes, or their interaction.

Place of residence, race and ethnicity, occupation, socioeconomic position, and other PROGRESS-Plus may shape both the likelihood and circumstances of sustaining a TBI and the probability of survival thereafter, yet these factors were rarely reported and integrated into the analyses of included studies (Table 1, S5

Appendix). Their omission is consequential because it limits assessment of whether observed sex differences in mortality are consistent across social groups or instead vary according to intersecting social and structural conditions. Daugherty et al. reported that TBI-related mortality rates among White and Black individuals in the United States were similar between 2001 and 2006 but in the following years became significantly higher among White individuals. This temporal divergence was attributed in part to a 32% increase in TBI-related suicide mortality among non-Hispanic White people between 2006-2008 and 2015-2017, with firearms accounting for approximately 97% of TBI-related suicides across racial and ethnic groups [58]. These findings suggest that differences associated with race or ethnicity cannot be understood independently of injury mechanism, social context, and other determinants of exposure to TBI and survival [58]. Accordingly, future TBI research should move beyond treating sex/gender, race, ethnicity, and other PROGRESS-Plus characteristics as isolated covariates, and instead adopt an intersectional framework that examines how these dimensions jointly structure TBI risk, access to care, and survivorship. In particular, understanding whether sex-related differences in mortality are amplified, attenuated, or reversed across racial and ethnic groups, occupations, socioeconomic positions, ages, and places of residence will be essential for distinguishing biological vulnerability from socially patterned exposure and structural disadvantage [59]. Intersectional analyses should therefore be a central priority in future TBI research aimed at explaining heterogeneity in mortality and targeting processes that generate unfair disparities in outcomes [60].

A key strength of our research is its equity-centered integration of systematic review, meta-analysis, and explanatory ML modelling. We restricted inclusion to studies that accounted for two major confounders of mortality - age and injury severity, examined differences in mortality ascertainment periods. We also systematically characterized study populations using PROGRESS-Plus factors and applied explainable ML to identify study-level characteristics associated with heterogeneity in sex-specific mortality estimates. Together, these approaches extend conventional evidence synthesis beyond estimating the overall association between sex and mortality by examining how population characteristics and study context may contribute to variation in reported effects. This integrated framework may also inform the development and evaluation of more transparent and generalizable data-driven prognostic approaches in TBI.

Several limitations should be considered. First, the explainable ML analyses were exploratory and based on study-level characteristics; therefore, the identified modifiers should be interpreted as hypothesis-generating rather than individual-level predictors or causal determinants of mortality. Second, substantial residual heterogeneity remained despite adjustment for age and injury severity, indicating that important clinical and health-care factors have rarely been reported across studies. If treatment timing and intensity and, or physiological response to treatment differs by sex, some of the observed variation in mortality may reflect differences arising during the care pathway rather than biological susceptibility alone [14].

Future studies of mortality and survivorship after TBI should therefore characterize and report key aspects of clinical management and examine whether treatment exposures and responses modify sex-associated differences in outcomes. This is important not only for improving causal interpretation of mortality differences, but also for identifying potentially modifiable aspects of care that could inform prevention of secondary injury, optimize care pathways, and reduce avoidable disparities in survival.

Third, we were unable to examine whether lifetime history of TBI contributed to sex differences in injury occurrence, injury severity, and reported mortality rates. Recurrent or cumulative TBI exposure may differ between males and females and could influence the severity and outcomes of the index injury [61, 62]. However, prior TBI history was inconsistently reported across studies, precluding assessment of its potential contribution to the observed sex-specific mortality associations. Incorporating longitudinal injury histories into future prognostic datasets may help clarify these relationships and improve the generalizability of digital prognostic models for TBI. In addition, studies did not apply a uniform definition of TBI [63, 64]. Although severity ascertainment may therefore have varied between studies, it was generally applied consistently within studies included in this review, supporting the validity of within-study comparisons of mortality risk between males and females across severity strata.

Fourth, the quality appraisal also identified several potential sources of bias, including limitations related to sample size, statistical power, and reporting quality, while the explanatory machine-learning analyses identified study quality as an important contributor to between-study variability. However, because these analyses were based on a relatively small number of studies and cohorts, the ML findings should be interpreted as exploratory and hypothesis-generating rather than confirmatory. Finally, the review was restricted to English-language publications, potentially excluding relevant evidence from non-English-speaking settings and limiting the global generalizability of the findings [65]. Future studies can address many of these limitations through standardized definitions of TBI severity, more complete reporting of lifetime injury exposure and treatment history, larger and more diverse study populations, and more consistent methodological and reporting practices.

## Conclusions

In conclusion, mortality following TBI differed by sex, with females showing lower mortality overall. Substantial between-study heterogeneity suggests that this association may vary according to population and methodological characteristics. Explainable ML identified age, sex composition, and study quality as potential contributors to this variation. These findings highlight the importance of representative populations, transparent reporting, and external validation when developing and evaluating digital prognostic models for TBI. Future digital health and prognostic research should prioritize standardized, comprehensive reporting and collection of demographic, clinical, and social characteristics in TBI datasets. Explainable ML can help identify patterns and sources of heterogeneity when relevant data are available but cannot recover information that was not collected. Strengthening the completeness, consistency, and interoperability of underlying research data is therefore essential for developing interpretable, transportable, and equitable data-driven approaches to TBI prognostication.

## Materials and Methods

### Protocol and registration

We registered the protocol in the prospective register of systematic reviews (PROSPERO 2025 CRD420251252136) [66, 67]. We followed the reporting guidelines of the Preferred Reporting Items for Systematic Reviews and Meta-Analyses (PRISMA) to conduct the systematic review [68].

### Data sources and search strategy

Systematic searches for included studies of all English peer-reviewed studies were conducted across eight databases, including PubMed, Embase, Web of Science, CENTRAL, MEDLINE, CINAHL, PsycINFO, and Scopus from inception (1996, 1947, 1971, 1961, 1971, 1984, 1967, and 2004, respectively) to August 18th, 2025. Reference lists of included studies have been reviewed to identify any additional relevant studies that may have been missed from searches. Three independent researchers (TT, DZ, and YY) screened titles, abstracts, and full-text reviews using Covidence software [69].

### PICO statement

A PICO framework was used to define the review question, inform the search strategy, and establish study eligibility criteria.

P (Population): Adults (≥18 years) with TBI. Studies without explicit age-based inclusion criteria were considered eligible if the reported mean or median age of the study sample was ≥18 years.

I (Intervention): Not applicable

C (Comparator): Female versus male participants O (Outcome): Mortality

Research question: Among adults with TBI, is sex associated with mortality, and which study-level characteristics modify the magnitude of this association across studies?

### Inclusion and exclusion criteria

We included published English-language observational studies (i.e. case-control, case-series, cross-sectional, and cohort studies) that included adults (≥18 years old) diagnosed with TBI from all severity levels (i.e., mild, moderate, and severe). Only studies that reported sex-specific mortality estimates and accounted for at least age and injury severity in adjusted analyses, or reported sex-specific subgroup analyses stratified by these factors were included. Studies that did not meet these criteria were excluded, with reasons for exclusion documented.

### Study selection

Two independent reviewers (DP and YY) screened titles, abstracts, and full text of original studies against the predefined PICO and eligibility criteria; the percent agreement reached 99%. Disagreements were resolved through discussion and consensus with a senior researcher (TM).

### Data extraction

Two researchers (DP and YY) independently extracted the following data from each included study: (1) study information (i.e., authors’ name, year of publication, country of origin, sample size, study objectives, design, and outcomes assessed and measured); (2) participant TBI-related variables (i.e., injury severity, external cause of injury, mechanisms of injury, mortality, comorbidities); (3) PROGRESS-Plus factors, including place of residence, race/ethnicity, occupation, gender/sex, religion, education, socioeconomic status, social capital, and relevant contextual factor [70]; statistical methods and covariates included in adjusted analyses; and (5) sex-specific findings.

For quantitative syntheses, we extracted reported sex-specific effect estimates, including odds ratios (ORs), relative risks (RRs), hazard ratios (HRs), and standardized mortality ratios (SMRs), and corresponding measures of uncertainty. We also extracted all variables identified and included by study authors as potential confounders of the association between sex and mortality.

### Risk of bias (quality) assessment

Study quality was assessed independently by two researchers (DP and YY) using the Quality In Prognosis Studies (QUIPS) tool developed by Hayden et al. [71] for evaluating risk of bias in prognostic studies. The appraisal followed two steps. In the first step, two researchers (DP and YY) independently assessed the items related to six potential sources of bias (study participation and attrition; prognostic factor and outcome measurements; confounding measurement and account; and analyses). At the second step, the same two reviewers judged the presence of potential biases as “Yes”, “Partly”, “No”, or “Unsure”. Following these steps, we utilized the Scottish Intercollegiate Guidelines Network (SIGN) methodology, where we assigned: “++” when all or most of the quality criteria are fulfilled (allowing one “Partly” while appraising all potential sources of bias); “+” when some of the criteria were fulfilled; and “-” when few or no criteria fulfilled (at least one “Yes”). In our review, we referred to group “++” as “high-quality studies” and group “+” as ‘moderate quality studies’. We abstracted data on the relationships between our outcomes of interest and primary (i.e., comorbidity) and secondary (i.e., socio-demographic, TBI-related) predictors only from studies with sufficient quality (i.e., high’ and ‘moderate’ quality studies).

### Data synthesis

#### Narrative synthesis

We extracted study-level data from all included studies, including sex-specific mortality outcomes, demographic characteristics, clinical variables, injury-related factors, and available social, contextual parameters (see 2.5), and adjusted outcome effect estimates (i.e., ORs, RRs, HRs, and SMRs). To ensure consistent interpretation, all effect estimates were harmonized to represent mortality in females relative to males, with males as the reference group. Estimates reported in the opposite direction were inverted by taking the reciprocal of the effect estimate and corresponding confidence interval (CI) limits, consistent with established meta-analytic methods [72]. We categorized mortality outcomes by reported time point or setting: in-hospital, specified time points, overall, and unspecified mortality.

We were guided by the PROGRESS-Plus framework when extracting potential moderators of the association between sex and mortality, including sample age, sex/gender composition, injury-related factors, social context (e.g., education and race/ethnicity), and country of study as a proxy for healthcare context [70]. We summarized distribution and completeness of demographic, clinical, and social parameters across included studies using descriptive statistics. For any study, if there was more than one study cohort or if participants of the study samples were divided into cohorts, we recorded the data for each cohort separately.

#### Meta-analysis

The primary meta-analytic endpoint was the pooled estimate of sex differences in mortality. Study-specific effect estimates were combined using a random-effects generic inverse-variance meta-analysis, with between-study variance estimated using restricted maximum likelihood (REML) [73]. Pooled ORs and HRs were reported with 95% CIs. Where sufficient data were available, separate meta-analyses were conducted by mortality type to assess whether the association between sex and mortality differed across outcome definitions. We assessed heterogeneity among study estimates using Cochran’s Q test, with a p value < 0.10 considered to represent evidence of heterogeneity [72]. To assess the magnitude of between study heterogeneity, we used an I² statistic estimate [74]; we considered I² value >50% to indicate substantial heterogeneity. Between-study variance was additionally estimated using τ², which represents the absolute variance of the true effect sizes across studies [75].

#### Publication bias

We assessed publication bias and small-study effects using funnel plots and Egger’s regression test [76, 77]. Effect estimates were transformed to the logarithmic scale and plotted against their standard errors, allowing different ratio measures (e.g., ORs, RRs, and SMRs) to be evaluated on a common scale. Funnel plot and Egger’s test analyses were not performed for HR meta-analyses because too few studies were available for reliable assessment [78].

#### Subgroup and sensitivity analyses

We explored potential sources of heterogeneity and assessed the robustness of pooled estimates using pre-specified subgroup and sensitivity analyses. Analyses were stratified by mortality ascertainment period (in-hospital, 1-, 3-, 6-, and 12-month mortality); overall and unspecified mortality were combined into a single category. Additional analyses were conducted by injury severity (i.e., severe TBI, moderate-to-severe TBI, and mixed-severity cohorts), study quality (i.e., moderate versus high), and study sample size (above versus below the median sample size).

All meta-analyses were conducted using R version 4.6.0 (R Foundation for Statistical Computing, Vienna, Austria) and RStudio version 2026.05.

#### Machine learning modelling

We anticipated that subgroup and sensitivity meta-analyses would provide limited insight into complex, nonlinear interactions among multiple study-level characteristics. We therefore conducted exploratory ML analyses to identify study-level features associated with variation in sex-specific mortality effect estimates and to characterize potential sources of between-study heterogeneity. The ML pipelines and analytic code have been described in detail in our previous work [79, 80].

#### Moderator coding and injury severity harmonization

For each study, we extracted social, demographic, and clinical characteristics as potential study-level moderators, guided by the PROGRESS-Plus framework [70]. PROGRESS factors included place of residence, race/ethnicity/culture/language, occupation, gender/sex, religion, education, socioeconomic status, and social capital. Plus factors included age, disability, and other context-specific characteristics relevant to TBI samples, including injury severity and mechanism of injury. Gender/sex composition of each study sample was coded as a proportion of male participants. When studies reported median age with interquartile range (IQR), we converted these values to estimated mean age and standard deviation (SD) using methods we applied and reported in our prior research [45, 81].

To harmonize injury severity across studies, cohorts were grouped into clinically relevant severity categories. Mixed-severity cohorts were classified as moderate-to-severe or severe TBI when more than 50% of participants fell within the respective severity category. Studies restricted to mild TBI were excluded from the ML analyses because only two eligible cohorts were available [23, 36].

#### Exploratory explainable ML analysis

After harmonization, candidate moderators were assessed for data completeness, and only variables reported in ≥80% of studies were retained to minimize missingness and reduce model instability. The retained moderators were entered into random forest (RF) [82, 83], gradient boosting (GB) [84, 85], and XGBoost [86] models to explore nonlinear associations and interactions contributing to between-study heterogeneity; we assessed consistency across models and interpreted results as exploratory.

#### Model performance

To evaluate each model performance, we used mean absolute error (MAE) and root mean square error (RMSE) [87].

#### Feature importance

Feature importance was assessed using feature importance heatmaps and the Shapley Additive Explanations (SHAP) values, which provided the contributions of each feature in terms of its predictive performance and effect on the difference between an actual prediction and a mean prediction [88].

#### Sensitivity analyses

We conducted three sensitivity analyses to evaluate the robustness of the ML findings.

First, the SD was added as an additional moderator to determine whether between-study variability in participant age influenced the importance of mean age or other moderators. Second, geographic region was added to the primary models to assess whether broader social, economic, cultural, and healthcare-system context influenced the feature importance of the Gender Inequality Index (GII) [89]. Third, study sample size was added as a moderator and dichotomized as above or below the median to assess whether between-study differences in sample size influenced feature-importance rankings.

#### Software and reporting

All ML analyses were conducted using were created using Python libraries (matplotlib (v3.x), seaborn (v0.x), and wordcloud (v1.x)) [18, 19]. We followed the Transparent Reporting of a multivariable prediction model for Individual Prognosis or Diagnosis - Artificial Intelligence (TRIPOD-AI) guidelines [20] to support transparent and reproducible reporting of the ML analyses.

#### Data and code availability

The full reproducible analysis code is available through the Open Science Framework (OSF), in accordance with the FAIR Guiding Principles [21].

## Data Availability

The minimal data set and accompanying code are available in the manuscript, and the submitted appendices.

## Acknowledgements

The authors thank Thaisa Tylinski Sant’Ana and Dian Zhang (KITE Research Institute – University Health Network and University of Toronto) for assistance with searches, abstract and full-text screening; trainee of the BRIDGE Lab (bridgelab.ca) Darren Kin-Tze Tang for data curation and contributions to the dataset for machine learning modelling and analyses. The authors also acknowledge Information Specialists from Library Services, Jessica Babineau and Cynthia Chui at the Toronto Rehabilitation Institute - University Health Network, for help with the comprehensive literature search. Resources and facilities were provided by the BRIDGE Lab, KITE Research Institute - University Health Network.

## Authorship Contributions

DP: Investigation; critical appraisal; data extraction; formal analysis; data visualization; scientific messages; equity, diversity, inclusion considerations; writing - original draft, review & editing.

YY: Investigation; critical appraisal; data extraction; scientific messages; equity, diversity, inclusion considerations; writing – review & editing.

US: Investigation; formal analysis; scientific messages; equity, diversity, inclusion considerations; equity, writing – review & editing.

NA: Investigation; equity, diversity, inclusion considerations; data visualization; writing – review & editing

TM: Conceptualization; methodology; searches; investigation; scientific messages; equity, diversity, inclusion considerations; validation; writing - review & editing; resources; funding acquisition; project supervision.

All authors contributed to the article, as well as read and approved the final manuscript.

## Declaration of Generative AI and AI-assisted Technologies in the Manuscript Preparation Process

During the preparation of this manuscript, the authors used ChatGPT (OpenAI) to assist with the development and refinement of R code used to generate heatmaps and meta-analysis figures, and Google Gemini to assist with the development and refinement of Python code used for the explainable machine-learning analyses. All AI-assisted code, analyses, and outputs were reviewed, tested, and revised by the authors before inclusion in the manuscript. The authors take full responsibility for the accuracy, integrity, and content of the published work. The final analysis code is attached as a supplement in S6 and S7.

## Supporting information

**S1 Appendix. Tables for search strategy and supporting documentation, exclusion reasons for studies assessed at full-text review, and quality assessment of observational studies using the NIH tool.**

**S2 Appendix. Funnel plot and Egger’s test and sensitivity analyses for meta-analyses**

**S3 Table. Performance of ML models for predicting modifiers of mortality in moderate-severe and severe TBI.**

**S4 Appendix. ML sensitivity analyses.**

**S5 Appendix. Mortality by PROGRESS-Plus parameters.**

**S6 Appendix. R codes used in analysis. Includes heatmap, forest plots, sensitivity analyses, funnel plots, and Egger’s tests.**

**S7 Appendix. Python code machine learning models**

