## Supplement material for "Evidence synthesis on sex differences in mortality after traumatic brain injury: Systematic review integrating meta-analysis and explainable machine learning"

*a KITE Toronto Rehabilitation Institute, University Health Network, Toronto, ON, Canada; b Faculty of Arts and Science, University of Toronto, Toronto, ON, Canada; c Dalla Lana School of Public Health, University of Toronto, Toronto, ON, Canada; d Temerty Faculty of Medicine, Rehabilitation Sciences Institute, University of Toronto, Toronto, ON, Canada; e Global Brain Health Institute, Trinity College Dublin, University of Dublin, Dublin, Ireland*

#### Supplementary aaterials

**S1 Table 1.** Search strategy and supporting documentation

SEARCH DETAILS

Searches were conducted in Cochrane Central Register of Controlled Trials (Ovid), CINAHL Ultimate (EBSCOhost), Embase (Ovid), Ovid MEDLINE (Epub Ahead of Print, In-Process & Other Non-Indexed Citations, Ovid MEDLINE(R) Daily and Ovid MEDLINE(R)), PsycINFO (Ovid), and Web of Science for an update of a systematic review (doi: 10.3389/fneur.2021.678971). In the initial review, searches covered from each database included to February 2018, with a repeat search of all databases in September 2019.

Search strategies were created by Jessica Babineau for the previous systematic review, which included the use of text words and subject headings (e.g. MeSH, Emtree) related to (1) traumatic brain injury and (2) sex or (3) gender. Searches were limited to human studies, and English language papers when possible. Case reports were excluded when possible. All searches were updated to include articles added to the database from September 2019 to March 26, 2024.

The updated search strategies included minor revisions (e.g. due to updates to databases):

- The search field for "kw" was changed to "kf" in Embase
- The search field "id" was added for PsycInfo, and "AK" for Web of Science to search for author keywords, where appropriate and consistent with the original Medline strategy from which it was translated from
- CINAHL changed to CINAHL Ultimate (broader coverage); specifying "TI" or "AB" in the search strategy, where appropriate and consistent with the original Medline strategy
- Truncation added for concuss* and postconcuss* in the Web of Science strategy

UPDATED SEARCH – NEW RESULTS PRE-DUPLICATE REMOVAL

TOTAL Results: 6839 citations

- Cochrane Central Register of Controlled Trials: 1292 citations
- CINAHL Ultimate: 573 citations
- Embase: 1754 citations
- MEDLINE: 1316 citations
- PsycINFO: 351 citations
- Web of Science: 1553 citations

SEARCH STRATEGIES

**Database:** Ovid MEDLINE: Epub Ahead of Print, In-Process & Other Non-Indexed Citations, Ovid MEDLINE(R) Daily and Ovid MEDLINE(R) <1946-Present>

| # | Query | Results from 25 Mar 2024 |
|---|---|---|
| 1 | Brain Injuries/ | 57,218 |
| 2 | exp Brain Injuries, Traumatic/ | 25,933 |
| 3 | exp Brain Concussion/ | 12,824 |
| 4 | Craniocerebral Trauma/ | 23,892 |
| 5 | tbi*2.tw,kf. | 36,995 |
| 6 | mtbi*2.tw,kf. | 4,338 |
| 7 | wrTBI*2.tw,kf. | 17 |
| 8 | concuss*.tw,kf. | 13,642 |
| 9 | postconcuss*.tw,kf. | 1,754 |
| 10 | ((head* or cerebr* or crani* or skull* or intracran*) adj2 (injur* or trauma* or damag* or wound* or swell* or oedema* or edema* or fracture* or contusion* or pressur*)).tw,kf. | 102,578 |
| 11 | ((brain* or cerebr* or intracerebr* or crani* or intracran* or head* or subdural* or epidural* or extradural*) adj (haematoma* or hematoma* or hemorrhag* or haemorrhag* or bleed*)).tw,kf. | 66,824 |
| 12 | or/1-11 | 241,567 |
| 13 | Sex Characteristics/ | 62,244 |
| 14 | exp sex distribution/ | 66,601 |
| 15 | exp Gender Identity/ | 27,958 |
| 16 | (sex adj2 (differenc* or charact*)).tw,kf. | 67,766 |
| 17 | sex-related.tw,kf. | 8,899 |
| 18 | sex-based.tw,kf. | 3,211 |
| 19 | (sexes adj2 (differenc* or charact*)).tw,kf. | 2,759 |
| 20 | (gender adj2 (differenc* or charact*)).tw,kf. | 53,107 |
| 21 | gender-related.tw,kf. | 6,249 |
| 22 | gender-based.tw,kf. | 5,861 |
| 23 | (male? or men or man or masculin*).ti,kf. | 343,391 |
| 24 | (female? or women or woman or feminin*).ti,kf. | 489,153 |

| | | |
|---|---|---|
| 25 | sex.ti,kf. | 148,257 |
| 26 | sexes.ti,kf. | 1,862 |
| 27 | gender*.ti,kf. | 81,068 |
| 28 | or/13-27 | 1,104,451 |
| 29 | 12 and 28 | 4,961 |
| 30 | 29 not (exp animals/ not exp humans/) | 4,548 |
| 31 | case reports/ | 2,391,969 |
| 32 | 30 not 31 | 3,968 |
| 33 | limit 32 to english language | 3,706 |
| 34 | (201909* or 20191* or 202*).dt,ez,da. | 7,705,588 |
| 35 | 33 and 34 | 1,316 |

**Database:** Cochrane Central Register of Controlled Trials <2014 to Present>

| # | Query | Results from 25 Mar 2024 |
|---|---|---|
| 1 | Brain Injuries/ | 2,501 |
| 2 | exp Brain Injuries, Traumatic/ | 1,638 |
| 3 | exp Brain Concussion/ | 660 |
| 4 | Craniocerebral Trauma/ | 446 |
| 5 | tbi*2.tw,kw. | 3,891 |
| 6 | mtbi*2.tw,kw. | 470 |
| 7 | wrTBI*2.tw,kw. | 0 |
| 8 | concuss*.tw,kw. | 1,068 |
| 9 | postconcuss*.tw,kw. | 319 |
| 10 | ((head* or cerebr* or crani* or skull* or intracran*) adj2 (injur* or trauma* or damag* or wound* or swell* or oedema* or edema* or fracture* or contusion* or pressur*)).tw,kw. | 6,969 |
| 11 | ((brain* or cerebr* or intracerebr* or crani* or intracran* or head* or subdural* or epidural* or extradural*) adj (haematoma* or hematoma* or hemorrhag* or haemorrhag* or bleed*)).tw,kw. | 9,532 |
| 12 | or/1-11 | 21,212 |

| | | |
|---|---|---|
| 13 | Sex Characteristics/ | 1,650 |
| 14 | exp sex distribution/ | 982 |
| 15 | exp Gender Identity/ | 380 |
| 16 | (sex adj2 (differenc* or charact*)).tw,kw. | 6,515 |
| 17 | sex-related.tw,kw. | 359 |
| 18 | sex-based.tw,kw. | 160 |
| 19 | (sexes adj2 (differenc* or charact*)).tw,kw. | 89 |
| 20 | (gender adj2 (differenc* or charact*)).tw,kw. | 3,028 |
| 21 | gender-related.tw,kw. | 242 |
| 22 | gender-based.tw,kw. | 313 |
| 23 | (male? or men or man or masculin*).ti,kw. | 426,415 |
| 24 | (female? or women or woman or feminin*).ti,kw. | 496,189 |
| 25 | sex.ti,kw. | 11,056 |
| 26 | sexes.ti,kw. | 33 |
| 27 | gender*.ti,kw. | 15,281 |
| 28 | or/13-27 | 569,609 |
| 29 | 12 and 28 | 7,146 |
| 30 | 29 not (exp animals/ not exp humans/) | 7,132 |
| 31 | case reports/ | 0 |
| 32 | 30 not 31 | 7,132 |
| 33 | limit 32 to english language | 6,982 |
| 34 | limit 33 to conference proceeding | 2,585 |
| 35 | 33 not 34 | 4,397 |
| 36 | (2019-09* or 2019-1* or 2022* or 2023* or 2024*).dl. | 341,603 |
| 37 | 35 and 36 | 1,292 |

**Database:** Embase <1974 to 2024 March 22>

| # | Query | Results from 25 Mar 2024 |
|---|---|---|
| 1 | exp traumatic brain injury/ | 70,114 |
| 2 | brain injury/ | 98,960 |
| 3 | exp concussion/ | 16,737 |
| 4 | head injury/ | 53,815 |
| 5 | tbi*2.tw,kf. | 60,091 |
| 6 | mtbi*2.tw,kf. | 6,955 |
| 7 | wrTBI*2.tw,kf. | 24 |
| 8 | concuss*.tw,kf. | 18,566 |
| 9 | postconcuss*.tw,kf. | 2,335 |
| 10 | ((head* or cerebr* or crani* or skull* or intracran*) adj2 (injur* or trauma* or damag* or wound* or swell* or oedema* or edema* or fracture* or contusion* or pressur*)).tw,kf. | 132,341 |
| 11 | ((brain* or cerebr* or intracerebr* or crani* or intracran* or head* or subdural* or epidural* or extradural*) adj (haematoma* or hematoma* or hemorrhag* or haemorrhag* or bleed*)).tw,kf. | 96,812 |
| 12 | or/1-11 | 390,358 |
| 13 | "gender and sex"/ | 1,262 |
| 14 | gender identity/ | 23,065 |
| 15 | (sex adj2 (differenc* or charact*)).tw,kf. | 87,082 |
| 16 | sex-related.tw,kf. | 10,935 |
| 17 | sex-based.tw,kf. | 4,251 |
| 18 | (sexes adj2 (differenc* or charact*)).tw,kf. | 3,619 |
| 19 | (gender adj2 (differenc* or charact*)).tw,kf. | 76,946 |
| 20 | gender-related.tw,kf. | 8,328 |
| 21 | gender-based.tw,kf. | 7,167 |
| 22 | (male? or men or man or masculin*).ti,kf. | 377,045 |
| 23 | (female? or women or woman or feminin*).ti,kf. | 592,436 |
| 24 | sex.ti,kf. | 165,307 |
| 25 | sexes.ti,kf. | 1,838 |
| 26 | gender*.ti,kf. | 108,989 |

| | | |
|---|---|---|
| 27 | or/13-26 | 1,209,704 |
| 28 | 12 and 27 | 7,074 |
| 29 | 28 not ((exp animals/ or exp animal experimentation/ or nonhuman/) not exp human/) | 5,859 |
| 30 | limit 29 to english language | 5,506 |
| 31 | case study/ | 99,911 |
| 32 | 30 not 31 | 5,476 |
| 33 | limit 32 to medline | 684 |
| 34 | 32 not 33 | 4,792 |
| 35 | limit 34 to dc=20190901-20240325 | 1,754 |

**Database:** APA PsycInfo <1806 to March Week 4 2024>

| # | Query | Results from 25 Mar 2024 |
|---|---|---|
| 1 | exp traumatic brain injury/ | 23,895 |
| 2 | head injuries/ | 4,842 |
| 3 | tbi*2.tw | 13,660 |
| 4 | mtbi*2.tw | 2,626 |
| 5 | wrTBI*2.tw | 9 |
| 6 | concuss*.tw | 4,942 |
| 7 | postconcuss*.tw | 1,027 |
| 8 | ((head* or cerebr* or crani* or skull* or intracran*) adj2 (injur* or trauma* or damag* or wound* or swell* or oedema* or edema* or fracture* or contusion* or pressur*)).tw | 13,850 |
| 9 | ((brain* or cerebr* or intracerebr* or crani* or intracran* or head* or subdural* or epidural* or extradural*) adj (haematoma* or hematoma* or hemorrhag* or haemorrhag* or bleed*)).tw | 4,050 |
| 10 | or/1-9 | 39,416 |
| 11 | human sex differences/ | 127,571 |
| 12 | exp gender identity/ | 49,814 |
| 13 | (sex adj2 (differenc* or charact*)).tw | 42,965 |
| 14 | sex-related.tw | 2,553 |

| | | |
|---|---|---|
| 15 | sex-based.tw | 779 |
| 16 | (sexes adj2 (differenc* or charact*)).tw | 561 |
| 17 | (gender adj2 (differenc* or charact*)).tw | 56,977 |
| 18 | gender-related.tw | 3,676 |
| 19 | gender-based.tw | 4,464 |
| 20 | (male? or men or man or masculin*).ti,id. | 218,106 |
| 21 | (female? or women or woman or feminin*).ti,id. | 239,935 |
| 22 | sex.ti,id. | 103,652 |
| 23 | sexes.ti,id. | 705 |
| 24 | gender*.ti,id. | 107,232 |
| 25 | or/11-24 | 600,099 |
| 26 | 10 and 25 | 2,071 |
| 27 | limit 26 to ("0200 book" or "0240 authored book" or "0280 edited book" or "0300 encyclopedia") | 69 |
| 28 | exp case report/ | 23,405 |
| 29 | 26 not (27 or 28) | 1,673 |
| 30 | limit 29 to english language | 1,600 |
| 31 | limit 30 to up=20190901-20240325 | 351 |

**Database:** CINAHL Ultimate

| # | Query | Limiters/Expanders | Last Run Via | Results from 25 March 2024 |
|---|---|---|---|---|
| S1 | (MH "Brain Injuries+") | Expanders - Apply equivalent subjects<br>Search modes - Boolean/Phrase | Interface - EBSCOhost Research Databases<br>Search Screen - Advanced Search<br>Database - CINAHL Ultimate | 34,219 |
| S2 | (MH "Brain Concussion+") | Expanders - Apply equivalent subjects<br>Search modes - Boolean/Phrase | Interface - EBSCOhost Research Databases<br>Search Screen - Advanced Search<br>Database - CINAHL Ultimate | 6,824 |

| | | | | |
|---|---|---|---|---|
| S3 | (MH "Head Injuries") | Expanders - Apply equivalent subjects<br>Search modes - Boolean/Phrase | Interface - EBSCOhost Research Databases<br>Search Screen - Advanced Search<br>Database - CINAHL Ultimate | 8,329 |
| S4 | TI (TBI##) OR AB (TBI##) | Expanders - Apply equivalent subjects<br>Search modes - Boolean/Phrase | Interface - EBSCOhost Research Databases<br>Search Screen - Advanced Search<br>Database - CINAHL Ultimate | 10,692 |
| S5 | TI (mTBI##) OR AB (mTBI##) | Expanders - Apply equivalent subjects<br>Search modes - Boolean/Phrase | Interface - EBSCOhost Research Databases<br>Search Screen - Advanced Search<br>Database - CINAHL Ultimate | 1,445 |
| S6 | TI (wrTBI##) OR AB (wrTBI##) | Expanders - Apply equivalent subjects<br>Search modes - Boolean/Phrase | Interface - EBSCOhost Research Databases<br>Search Screen - Advanced Search<br>Database - CINAHL Ultimate | 10 |
| S7 | TI (concuss*) OR AB (concuss*) | Expanders - Apply equivalent subjects<br>Search modes - Boolean/Phrase | Interface - EBSCOhost Research Databases<br>Search Screen - Advanced Search<br>Database - CINAHL Ultimate | 6,803 |
| S8 | TI (postconcuss*) OR AB (postconcuss*) | Expanders - Apply equivalent subjects<br>Search modes - Boolean/Phrase | Interface - EBSCOhost Research Databases<br>Search Screen - Advanced Search<br>Database - CINAHL Ultimate | 925 |
| S9 | TI ((head* or cerebr* or crani* or skull* or intracran*) n2 (injur* or trauma* or damag* or wound* or swell* or oedema* or edema* or fracture* or contusion* or pressur*)) OR AB ((head* or cerebr* or crani* or skull* or intracran*) n2 (injur* or | Expanders - Apply equivalent subjects<br>Search modes - Boolean/Phrase | Interface - EBSCOhost Research Databases<br>Search Screen - Advanced Search<br>Database - CINAHL Ultimate | 24,667 |

| | | | | |
|---|---|---|---|---|
| | trauma* or damag* or wound* or swell* or oedema* or edema* or fracture* or contusion* or pressur*)) | | | |
| S10 | TI ((brain* or cerebr* or intracerebr* or crani* or intracran* or head* or subdural* or epidural* or extradural*) n1 (haematoma* or hematoma* or hemorrhag* or haemorrhag* or bleed*)) OR AB ((brain* or cerebr* or intracerebr* or crani* or intracran* or head* or subdural* or epidural* or extradural*) n1 (haematoma* or hematoma* or hemorrhag* or haemorrhag* or bleed*)) | Expanders - Apply equivalent subjects<br>Search modes - Boolean/Phrase | Interface - EBSCOhost Research Databases<br>Search Screen - Advanced Search<br>Database - CINAHL Ultimate | 15,523 |
| S11 | S1 OR S2 OR S3 OR S4 OR S5 OR S6 OR S7 OR S8 OR S9 OR S10 | Expanders - Apply equivalent subjects<br>Search modes - Boolean/Phrase | Interface - EBSCOhost Research Databases<br>Search Screen - Advanced Search<br>Database - CINAHL Ultimate | 73,362 |
| S12 | (MH "Gender Identity+") | Expanders - Apply equivalent subjects<br>Search modes - Boolean/Phrase | Interface - EBSCOhost Research Databases<br>Search Screen - Advanced Search<br>Database - CINAHL Ultimate | 10,157 |
| S13 | TI (sex n2 (differenc* or charact*)) OR AB (sex n2 (differenc* or charact*)) | Expanders - Apply equivalent subjects<br>Search modes - Boolean/Phrase | Interface - EBSCOhost Research Databases<br>Search Screen - Advanced Search<br>Database - CINAHL Ultimate | 17,294 |
| S14 | TI (sex-related) OR AB (sex-related) | Expanders - Apply equivalent subjects<br>Search modes - Boolean/Phrase | Interface - EBSCOhost Research Databases<br>Search Screen - Advanced Search<br>Database - CINAHL Ultimate | 1,825 |

| | | | | |
|---|---|---|---|---|
| S15 | TI (sex-based) OR AB (sex-based) | Expanders - Apply equivalent subjects<br>Search modes - Boolean/Phrase | Interface - EBSCOhost Research Databases<br>Search Screen - Advanced Search<br>Database - CINAHL Ultimate | 993 |
| S16 | TI (sexes n2 (differenc* or charact*)) OR AB (sexes n2 (differenc* or charact*)) | Expanders - Apply equivalent subjects<br>Search modes - Boolean/Phrase | Interface - EBSCOhost Research Databases<br>Search Screen - Advanced Search<br>Database - CINAHL Ultimate | 17,294 |
| S17 | TI (gender n2 (differenc* or charact*)) OR AB (gender n2 (differenc* or charact*)) | Expanders - Apply equivalent subjects<br>Search modes - Boolean/Phrase | Interface - EBSCOhost Research Databases<br>Search Screen - Advanced Search<br>Database - CINAHL Ultimate | 23,176 |
| S18 | TI (gender-related) OR AB (gender-related) | Expanders - Apply equivalent subjects<br>Search modes - Boolean/Phrase | Interface - EBSCOhost Research Databases<br>Search Screen - Advanced Search<br>Database - CINAHL Ultimate | 1,803 |
| S19 | TI (male# or men or man or masculin*) | Expanders - Apply equivalent subjects<br>Search modes - Boolean/Phrase | Interface - EBSCOhost Research Databases<br>Search Screen - Advanced Search<br>Database - CINAHL Ultimate | 88,918 |
| S20 | TI (female# or women or woman or feminin*) | Expanders - Apply equivalent subjects<br>Search modes - Boolean/Phrase | Interface - EBSCOhost Research Databases<br>Search Screen - Advanced Search<br>Database - CINAHL Ultimate | 222,940 |
| S21 | TI (sex or sexes or gender*) | Expanders - Apply equivalent subjects<br>Search modes - Boolean/Phrase | Interface - EBSCOhost Research Databases<br>Search Screen - Advanced Search<br>Database - CINAHL Ultimate | 69,117 |
| S22 | S12 OR S13 OR S14 OR S15 OR S16 OR S17 OR S18 OR S19 OR S20 OR S21 | Expanders - Apply equivalent subjects | Interface - EBSCOhost Research Databases | 374,138 |

| | | | | |
|---|---|---|---|---|
| | | Search modes - Boolean/Phrase | Search Screen - Advanced Search<br>Database - CINAHL Ultimate | |
| S23 | S11 AND S22 | Expanders - Apply equivalent subjects<br>Search modes - Boolean/Phrase | Interface - EBSCOhost Research Databases<br>Search Screen - Advanced Search<br>Database - CINAHL Ultimate | 1,626 |
| S24 | PT (case study) | Expanders - Apply equivalent subjects<br>Search modes - Boolean/Phrase | Interface - EBSCOhost Research Databases<br>Search Screen - Advanced Search<br>Database - CINAHL Ultimate | 449,333 |
| S25 | TI (case n1 (study or report)) | Expanders - Apply equivalent subjects<br>Search modes - Boolean/Phrase | Interface - EBSCOhost Research Databases<br>Search Screen - Advanced Search<br>Database - CINAHL Ultimate | 118,802 |
| S26 | (MH "Case Studies") | Expanders - Apply equivalent subjects<br>Search modes - Boolean/Phrase | Interface - EBSCOhost Research Databases<br>Search Screen - Advanced Search<br>Database - CINAHL Ultimate | 27,900 |
| S27 | S24 OR S25 OR S26 | Expanders - Apply equivalent subjects<br>Search modes - Boolean/Phrase | Interface - EBSCOhost Research Databases<br>Search Screen - Advanced Search<br>Database - CINAHL Ultimate | 517,068 |
| S28 | S23 NOT S27 | Expanders - Apply equivalent subjects<br>Search modes - Boolean/Phrase | Interface - EBSCOhost Research Databases<br>Search Screen - Advanced Search<br>Database - CINAHL Ultimate | 1,432 |
| S29 | S28 | Limiters - English Language<br>Expanders - Apply equivalent subjects<br>Search modes - Boolean/Phrase | Interface - EBSCOhost Research Databases<br>Search Screen - Advanced Search<br>Database - CINAHL Ultimate | 1,427 |

| | | | | |
|---|---|---|---|---|
| S30 | (EM 20190901- OR (ZD "in process" AND RD 20190901-)) | Limiters - English Language<br>Expanders - Apply equivalent subjects<br>Search modes - Boolean/Phrase | Interface - EBSCOhost Research Databases<br>Search Screen - Advanced Search<br>Database - CINAHL Ultimate | 2,306,501 |
| S31 | S29 AND S30 | Expanders - Apply equivalent subjects<br>Search modes - Boolean/Phrase | Interface - EBSCOhost Research Databases<br>Search Screen - Advanced Search<br>Database - CINAHL Ultimate | 573 |

**Database:** Web of Science Core Collection
Date Searched: March 25, 2024
(Indexes: WOS.SCI: 1900 to 2024; WOS.AHCI: 1975 to 2024; WOS.BHCI: 2005 to 2024; WOS.BSCI: 2005 to 2024; WOS.ESCI: 2005 to 2024; WOS.ISTP: 1990 to 2024; WOS.SSCI: 1900 to 2024; WOS.ISSHP: 1990 to 2024)

| # | Search Query | Results from 25 March 2024 |
|---|---|---|
| 1 | TS=(tbi* OR mTBI* OR wrTBI* OR concuss OR postconcuss) | 48329 |
| 2 | TS=((head* or cerebr* or crani* or skull* or intracran*) NEAR/2 (injur* or trauma* or damag* or wound* or swell* or oedema* or edema* or fracture* or contusion* or pressur*)) | 132810 |
| 3 | TS=((brain* or cerebr* or intracerebr* or crani* or intracran* or head* or subdural* or epidural* or extradural*) NEAR/1 (haematoma* or hematoma* or hemorrhag* or haemorrhag* or bleed*)) | 75346 |
| 4 | #1 OR #2 OR #3 | 231009 |
| 5 | TS=(sex NEAR/2 (differenc* OR charact*)) | 134669 |
| 6 | TS=(sex-related) | 10803 |
| 7 | TS=(sex-based) | 4421 |
| 8 | TS=(sexes NEAR/2 (differenc* OR charact*)) | 134669 |
| 9 | TS=(gender NEAR/2 (differenc* OR charact*)) | 149115 |
| 10 | TS=(gender-related) | 9796 |

| | | |
|---|---|---|
| 11 | TS=(gender-based) | 12696 |
| 12 | (TI=(male* OR men OR man OR masculin*)) OR AK=(male* OR men OR man OR masculin*) | 638957 |
| 13 | (TI=(female* OR women OR woman OR feminin*)) OR AK=(female* OR women OR woman OR feminin*) | 898347 |
| 14 | (TI=(sex or sexes)) OR AK=(sex or sexes) | 254247 |
| 15 | #5 OR #6 OR #7 OR #8 OR #9 OR #10 OR #11 OR #12 OR #13 OR #14 | 1830682 |
| 16 | #4 AND #15 | 4222 |
| 17 | (#16) AND LA=(English) | 4114 |
| 18 | #17 Timespan: 2019-09-01 to 2024-03-25 | 1553 |

**S1 Table 2.** Exclusion reasons for studies assessed at full-text review (n=141)

| Author (Year) | Title | Exclusion Reason |
|---|---|---|
| 1. Adams 2023 | Pregnancy, fetal, and neonatal outcomes among women with traumatic brain injury | No sex comparison |
| 2. Adams 2025 | PACAP a mediator of inflammation following trauma exposure and mild traumatic brain injury: Differential effects in males and females | Lack of relevant data: No mortality-specific data presented |
| 3. Adhikari 2024 | A Four-Country Study of Strangulation-Related Alterations in Consciousness in Women Who Have Experienced Intimate Partner Violence: Co-Occurrence with Traumatic Brain Injuries and Measures of Psychological Distress | Female only; Based more on S-AICs |
| 4. Ahmadi 2015 | Traumatic brain injury, coronary atherosclerosis and cardiovascular mortality | No comparison of mortality and sex |
| 5. Ahmed 2020 | Early risk stratification of in hospital mortality following a ground level fall in geriatric patients with normal physiological parameters | Lack of relevant data: No specification of TBIs, only falls |
| 6. Aiolfi 2017 | Brain trauma foundation guidelines for intracranial pressure monitoring: compliance and effect on outcome | Cannot access |
| 7. Amara 2018 | Predictors of Employment Status in Male and Female Post-9/11 Veterans Evaluated for Traumatic Brain Injury | Lack of relevant data: No mortality-specific data presented |
| 8. Andelic 2021 | Frequency of fatigue and its changes in the first 6 months after traumatic brain injury: results from the CENTER-TBI study | Lack of relevant data: No mortality-specific data presented |
| 9. Anderson 2023 | COMT Val158Met and BDNF Val66Met Single-Nucleotide Polymorphisms Are Not Associated With Emotional Distress One Year After Moderate-Severe Traumatic Brain Injury. | Lack of relevant data: No mortality-specific data presented |
| 10. Anderson 2025 | The Relationship Between Sex, Fatigue, and Post-Concussion Symptoms Differs Between Those With Mild Traumatic Brain Injury Versus Trauma Control and Healthy Control Participants | Lack of relevant data: No mortality-specific data presented |
| 11. Arellano-Orden 2011 | Gender Influences Cerebral Oxygenation After Red Blood Cell Transfusion in Patients with Severe Traumatic Brain Injury | Lack of relevant data: No mortality-specific data presented |
| 12. Aude 2025 | Machine learning reveals demographic disparities in palliative care timing among patients with traumatic brain injury receiving neurosurgical consultation | No, no mortality comparisons |
| 13. Babakhanyan 2024 | Gender disparities in neurobehavioral symptoms and the role of post-traumatic symptoms in US service members following mild traumatic brain injury | Lack of relevant data: No mortality-specific data presented |
| 14. Bernanke 2025 | Traumatic Brain Injury and Suicidal Thoughts and Behaviors among Post-9/11 Veterans: Investigating Longitudinal Change and Interactions with Mental Health | Lack of relevant data: No mortality-specific data presented |
| 15. Bosarge 2015 | Stress-induced hyperglycemia is associated with higher mortality in severe traumatic brain injury | No comparison of mortality and sex |
| 16. Boutin 2016 | Transfusion of red blood cells in patients with traumatic brain injuries admitted to Canadian trauma health centres: a multicentre cohort study | No comparison of mortality and sex |
| 17. Brandel 2017 | The Association between Psychiatric Comorbidities and Outcomes for Inpatients with Traumatic Brain Injury | Injury severity not specified and/or considered |

| | | |
|---|---|---|
| 18. Brown 2012 | Mental Illness in the 2 Years Prior to Pregnancy in a Population With Traumatic Brain Injury: A Cross-Sectional Study: La maladie mentale dans les deux ans précédant une grossesse dans une population souffrant de lésion cérébrale traumatique : une étude transversale | Lack of relevant data: No mortality-specific data presented |
| 19. Brown 2024 | Mental Illness in the 2 Years Prior to Pregnancy in a Population With Traumatic Brain Injury: A Cross-Sectional Study: La maladie mentale dans les deux ans précédant une grossesse dans une population souffrant de lésion cérébrale traumatique : une étude transversale | No sex comparison |
| 20. Chan 2020 | Data Mining to Understand How Health Status Preceding Traumatic Brain Injury Affects Functional Outcome: A Population-Based Sex-Stratified Study | Lack of relevant data: No mortality-specific data presented |
| 21. Chen 2023 | Effects of Age, Sex, and Postconcussive Symptoms on Domain-Specific Quality of Life a Year After Mild Traumatic Brain Injury | Lack of relevant data: No mortality-specific data presented |
| 22. Chen 2024 | Effects of age, sex, and postconcussive symptoms on domain-specific quality of life a year after mild traumatic brain injury | Lack of relevant data: No mortality-specific data presented |
| 23. Cheng 2015 | One-Year Mortality after Traumatic Brain Injury in Liver Cirrhosis Patients—A Ten-Year Population-Based Study | Injury severity not specified and/or considered |
| 24. Chinese Head Trauma Study Collaborators 2021 | Chinese Head Trauma Data Bank: Effect of Gender on the Outcome of Patients With Acute Traumatic Brain Injury | No comparison of mortality and sex with adjustments |
| 25. Coimbra 2003 | Does Sexual Dimorphism Influence Outcome of Traumatic Brain Injury Patients? The Answer Is No! | No comparison of mortality and sex with adjustments |
| 26. Colantonio 2008 | Predictors of Postacute Mortality Following Traumatic Brain Injury in a Seriously Injured Population | No comparison of mortality and sex |
| 27. Corrigan 2007 | Employment After Traumatic Brain Injury: Differences Between Men and Women | Lack of relevant data: No mortality-specific data presented |
| 28. Cote 2012 | Factors Associated with the Withdrawal of Life-Sustaining Therapies in Patients with Severe Traumatic Brain Injury: A Multicenter Cohort Study | Lack of relevant data: No mortality-specific objective or hypothesis |
| 29. Dahdah 2016 | Variations in Inpatient Rehabilitation Functional Outcomes Across Centers in the Traumatic Brain Injury Model Systems Study and the Influence of Demographics and Injury Severity on Patient Outcomes | Lack of relevant data: No mortality-specific data presented |
| 30. Dams O Connor 2016 | Health Problems Precede Traumatic Brain Injury in Older Adults | Lack of relevant data: No mortality-specific data presented |
| 31. de Koning 2017 | Prediction of work resumption and sustainability up to 1 year after mild traumatic brain injury | Lack of relevant data: No mortality-specific data presented |
| 32. DeStefano 2025 | Objectively Measured Daytime Napping Patterns and All-Cause Mortality in Older Adults | No sex comparison |
| 33. Eggertsdottir Claesssen 2025 | Pituitary Dysfunction Following Mild Traumatic Brain Injury in Female Athletes: Neuropsychological and Psychological Findings | Self-reported concussion |

| | | |
|---|---|---|
| 34. Elkbuli 2020 | Mild and Moderate Traumatic Brain Injury and Gender-Based Critical Care Outcomes | No comparison of mortality and sex with adjustments |
| 35. El-Menyar 2019 | Females fall more from heights but males survive less among a geriatric population: insights from an American level 1 trauma center | Only falls; No comparison of mortality and sex with adjustments |
| 36. Eramudugolla 2014 | Long-Term Cognitive Correlates of Traumatic Brain Injury across Adulthood and Interactions with APOE Genotype, Sex, and Age Cohorts | Lack of relevant data: No mortality-specific data presented |
| 37. Falk 2015 | Prehospital management of traumatic brain injury patients – a gender perspective | No comparison of mortality and sex with adjustments |
| 38. Fleischer 2025 | Sex and age-related implications for preventive measures of intensive care admitted traumatic brain injury patients in Switzerland: an observational study | Lack of relevant data: No mortality-specific data presented |
| 39. Forslund 2017 | The trajectories of overall disability in the first 5 years after moderate and severe traumatic brain injury | Lack of relevant data: No mortality-specific data presented |
| 40. Gan 2004 | Outcome of Moderate and Severe Traumatic Brain Injury Amongst the Elderly in Singapore | No comparison of mortality and sex with adjustments |
| 41. Gao 2017 | Early Asymmetric Cardio-Cerebral Causality and Outcome after Severe Traumatic Brain Injury | No comparison of mortality and sex with adjustments |
| 42. Gerhart 2003 | Violence-Related Traumatic Brain Injury: A Population-Based Study | Lack of relevant data: No mortality-specific data presented |
| 43. Glenn 2001 | Depression amongst outpatients with traumatic brain injury | Lack of relevant data: No mortality-specific data presented |
| 44. Gosling 1999 | Rearranged marriages: marital relationships after head injury | Lack of relevant data: No mortality-specific data presented |
| 45. Griesdale 2009 | Glucose Control and Mortality in Patients with Severe Traumatic Brain Injury | No comparison of mortality and sex |
| 46. Gujral 2006 | Sex differences in mortality after traumatic brain injury, Colorado 1994–1998 | No comparison of mortality and sex with adjustments |
| 47. Gutman 1996 | The Experience of Head Injury on the Impairment of Gender Identity and Gender Role | Lack of relevant data: No mortality-specific data presented |
| 48. Harrison 2006 | Risk of schizophrenia and other non-affective psychosis among individuals exposed to head injury: Case control study | Lack of relevant data: No mortality-specific data presented |
| 49. Herrera-Melero 2015 | Acute predictors for mortality after severe TBI in Spain: Gender differences and clinical data | No comparison of mortality and sex with adjustments |
| 50. Hoffman 2012 | Change in Inpatient Rehabilitation Admissions for Individuals With Traumatic Brain Injury After Implementation of the Medicare Inpatient Rehabilitation Facility Prospective Payment System | Lack of relevant data: No mortality-specific data presented |
| 51. Iderdar 2025 | Cross-cultural adaptation of the Quality of Life after Brain Injury (QOLIBRI) questionnaire in patients after traumatic brain injury in Morocco | Lack of relevant data: No mortality-specific data presented |
| 52. Jain 2025 | Factors Associated With Return to Activity After Concussion Among Female Service Academy Members: Findings From the NCAA–DoD CARE Consortium | No sex comparison |

| | | |
|---|---|---|
| 53. Jak 2024 | Sex differences in postconcussive symptom reporting in those with history of concussion: Findings from the federal interagency traumatic brain injury research (FITBIR) database | Review Study |
| 54. Jovanovic 2016 | Twenty-Eight-Day Mortality of Blunt Traumatic Brain Injury and Co-Injuries Requiring Mechanical Ventilation | No comparison of mortality and sex |
| 55. Jung 2014 | Prognostic Factors of Neurocognitive and Functional Outcomes in Junior and Senior Elderly Patients with Traumatic Brain Injury Undergoing Disability Evaluation or Appointed Disability Evaluation | Lack of relevant data: No mortality-specific data presented |
| 56. Jung 2024 | Sex-specific impacts of obesity on long-term prognosis of traumatic brain injury: a multicenter prospective study | No comparison of mortality and sex with adjustments |
| 57. Kadar 2019 | Trends in demographics and outcome of patients presenting with traumatic brain injury | No comparison of mortality and sex with adjustments |
| 58. Kashyap 2025 | Gender Differences in Seeking and Receiving Healthcare After Mild Traumatic Brain Injury | Lack of relevant data: No mortality-specific data presented |
| 59. Kennedy 2024 | Phenotyping Depression After Mild Traumatic Brain Injury: Evaluating the Impact of Multiple Injury, Gender, and Injury Context | Lack of relevant data: No mortality-specific data presented |
| 60. Khan 2019 | Predictors of Outcome After Traumatic Brain Injuries: Experience of a Tertiary Health Care Institution in Northwest India | No comparison of mortality and sex with adjustments |
| 61. Kinney 2023 | Identifying and Predicting Subgroups of Veterans With Mild Traumatic Brain Injury Based on Distinct Configurations of Postconcussive Symptom Endorsement: A Latent Class Analysis | Lack of relevant data: No mortality-specific data presented |
| 62. Kirkness 2004 | Is There a Sex Difference in the Course Following Traumatic Brain Injury? | Lack of relevant data: No mortality-specific data presented |
| 63. Kisat 2012 | Predictors of Positive Head CT Scan and Neurosurgical Procedures After Minor Head Trauma | Lack of relevant data: No mortality-specific objective or hypothesis |
| 64. Klauber 1981 | Prospective Study of Patients Hospitalized with Head Injury in San Diego County, 1978 | No comparison of mortality and sex with adjustments |
| 65. Kokkinou 2020 | A population study on the epidemiology and outcome of brain injuries in intensive care | No comparison of mortality and sex with adjustments |
| 66. Kraus 2000 | The independent effect of gender on outcomes following traumatic brain injury: a preliminary investigation | No comparison of mortality and sex with adjustments |
| 67. LeBlanc 2021 | Cognitive, communication performance following mild traumatic brain injury: Influence of sex, age, education, site of lesion and bilingualism. | Lack of relevant data: No mortality-specific data presented |
| 68. Lee 2014 | Increased risk of ischemic stroke in patients with mild traumatic brain injury: a nationwide cohort study | Lack of relevant data: No mortality-specific data presented |
| 69. Leitbeg 2011 | Effects of Gender on Outcomes After Traumatic Brain Injury | No comparison of mortality and sex with adjustments |

| | | |
|---|---|---|
| 70. Lequerica 2025 | The Interaction Between Race/Ethnicity and Sex on Societal Participation Among Individuals With Traumatic Brain Injury: A NIDILRR Model Systems Study | Lack of relevant data: No mortality-specific data presented |
| 71. Ley 2013 | Gender impacts mortality after traumatic brain injury in teenagers | Age inclusion criteria not met |
| 72. Li 2018 | Effect of Age and Sex on Hospital Readmission in Traumatic Brain Injury | Lack of relevant data: No mortality-specific data presented |
| 73. Lian 2025 | Community Integration Challenges of Men and Women After Traumatic Brain Injury: A Reflexive Thematic Analysis of Lived Experiences Through a Gender Lens | Lack of relevant data: No mortality-specific data presented |
| 74. Liao 2012 | Risk and outcomes for traumatic brain injury in patients with mental disorders | Injury severity not specified and/or considered |
| 75. Lolk 2021 | Repeated traumatic brain injury and risk of epilepsy: A Danish nationwide cohort study. | Lack of relevant data: No mortality-specific data presented |
| 76. Ma 2021 | Screening for Poor Self-Reported Sleep Quality at 12 Weeks in Post-Mild Traumatic Brain Injury Patients Using the HF–Age–Gender (HAG) Index | Lack of relevant data: No mortality-specific data presented |
| 77. MacGregor 2024 | Sex Differences in Postinjury Health Profiles Among U.S. Military Personnel Following Deployment-Related Concussion. | Lack of relevant data: No mortality-specific data presented |
| 78. Mair 2022 | The outcome of severely injured patients following traumatic brain injury is affected by gender—A retrospective, multicenter, matched-pair analysis utilizing data of the TraumaRegister DGU | No comparison of mortality and sex with adjustments |
| 79. Marino 2006 | Posttraumatic cerebral infarction in patients with moderate or severe head trauma | Cannot access |
| 80. Martins 2009 | Mortality in Severe Traumatic Brain Injury: A Multivariated Analysis of 748 Brazilian Patients From Floriano´polis City | No comparison of mortality and sex with adjustments |
| 81. Meares 2007 | Mild traumatic brain injury does not predict acute postconcussion syndrome | Lack of relevant data: No mortality-specific data presented |
| 82. Mellick 2003 | Understanding outcomes based on the post-acute hospitalization pathways followed by persons with traumatic brain injury | Lack of relevant data: No mortality-specific data presented |
| 83. Mikolic 2021 | Explaining Outcome Differences between Men and Women following Mild Traumatic Brain Injury | Lack of relevant data: No mortality-specific data presented |
| 84. Miles 2015 | Examining the Influence of Mild Traumatic Brain Injury and Posttraumatic Stress Disorder on Alcohol Use Disorder in OEF/OIF Veterans | Lack of relevant data: No mortality-specific data presented |
| 85. Minglis 2025 | Sex Differences in Effects of Concussion History on Select Subscales From the Automated Neuropsychological Assessment Metrics (ANAM): Preliminary Findings of Performance in High School and Collegiate Athletes | Sport-related concussion |
| 86. Mollayeva 2015 | Modeling community integration in workers with delayed recovery from mild traumatic brain injury | Lack of relevant data: No mortality-specific data presented |
| 87. Mollayeva 2016 | Insomnia in workers with delayed recovery from mild traumatic brain injury | Lack of relevant data: No mortality-specific data presented |
| 88. Mollayeva 2016 | The relationship between insomnia and disability in workers with mild traumatic brain injury/concussion: Insomnia and disability in chronic mild traumatic brain injury | Lack of relevant data: No mortality-specific data presented |

| | | |
|---|---|---|
| 89. Mollayeva 2017 | Concussion/mild traumatic brain injury-related chronic pain in males and females: A diagnostic modelling study | Lack of relevant data: No mortality-specific data presented |
| 90. Mollayeva 2017 | Sleep stage distribution in persons with mild traumatic brain injury: a polysomnographic study according to American Academy of Sleep Medicine standards | Lack of relevant data: No mortality-specific data presented |
| 91. Mollayeva 2019 | Sex-specific incident dementia in patients with central nervous system trauma | Lack of relevant data: No mortality-specific data presented |
| 92. Mollayeva 2019 | Knowledge of sex and gender and related information needs in patients with traumatic brain injury: in-depth interview study | Lack of relevant data: No mortality-specific data presented |
| 93. Mollayeva 2023 | The effect of sleep disorders on dementia risk in patients with traumatic brain injury: A large-scale cohort study | Lack of relevant data: No mortality-specific data presented |
| 94. Morrisson 2004 | Gender and age effects on outcome after pediatric traumatic brain injury | Age inclusion criteria not met |
| 95. Munivenkatappa 2016 | Traumatic brain injury: Does gender influence outcomes? | No comparison of mortality and sex with adjustments |
| 96. Myrga 2016 | A Dopamine Pathway Gene Risk Score for Cognitive Recovery Following Traumatic Brain Injury: Methodological Considerations, Preliminary Findings, and Interactions With Sex | Lack of relevant data: No mortality-specific data presented |
| 97. Nguyen 2014 | Effect of marijuana use on outcomes in traumatic brain injury | Cannot access |
| 98. O'Reilly 2023 | Hospitalisations and in-hospital deaths following moderate to severe traumatic brain injury in Australia, 2015–20: a registry data analysis for the Australian Traumatic Brain Injury National Data (ATBIND) project | No comparison of mortality and sex with adjustments |
| 99. Ostberg 2014 | Smoking and outcome of traumatic brain injury | Lack of relevant data: No mortality-specific data presented |
| 100. Payen 2025 | Sex differences in neurological outcome at 6 and 12 months following severe traumatic brain injury. An observational analysis of the OXY-TC trial | No comparison of mortality and sex |
| 101. Peck 2014 | Death after discharge: Predictors of mortality in older brain-injured patients | No comparison of mortality and sex with adjustments |
| 102. Pogoda 2012 | Multisensory impairment reported by veterans with and without mild traumatic brain injury history | Lack of relevant data: No mortality-specific data presented |
| 103. Portiz 2019 | Gender Differences in Employment and Economic Quality of Life Following Traumatic Brain Injury | Lack of relevant data: No mortality-specific data presented |
| 104. Rappold 2002 | Female Gender Does Not Protect Blunt Trauma Patients from Complications and Mortality | No comparison of mortality and sex with adjustments |
| 105. Rao 2008 | Prevalence and types of sleep disturbances acutely after traumatic brain injury | Lack of relevant data: No mortality-specific data presented |
| 106. Renner 2012 | The influence of gender on the injury severity, course and outcome of traumatic brain injury | Lack of relevant data: No mortality-specific data presented |

| | | |
|---|---|---|
| 107.Ringey 2025 | Long-Term Brain Health Outcomes in Females With a History of Contact Sports: A Cross-Sectional Survey Analysis | Age, Sport-related concussion, Survey |
| 108.Robles 2021 | Older age, male sex, and cerebral microbleeds predict white matter loss after traumatic brain injury | Lack of relevant data: No mortality-specific data presented |
| 109.Rowe 2022 | Sex-based differences in outcomes for adult patients presenting to the emergency department with a concussion | Lack of relevant data: No mortality-specific data presented |
| 110.Ruiter-Lopez 2024 | Resilience and Concussion Recovery in Minority Women: Promoting Health Equity | Female only |
| 111.Rush 2016 | Craniotomy vs craniectomy for acute traumatic subdural hematoma in the United States: a national retrospective cohort analysis | No comparison of mortality and sex |
| 112.Saatian 2018 | Epidemiology and Pattern of Traumatic Brain Injury in a Developing Country; Regional Trauma Center | No comparison of mortality and sex with adjustments |
| 113.Saban 2011 | Sex Differences in Perceived Life Satisfaction and Functional Status One Year After Severe Traumatic Brain Injury | Lack of relevant data: No mortality-specific data presented |
| 114.Scheetz 2015 | Injury patterns, severity and outcomes among older adults who sustained brain injury following a same level fall: A retrospective analysis | No comparison of mortality and sex |
| 115.Schiraldi 2015 | Effect of insurance and racial disparities on outcomes in traumatic brain injury | Cannot access |
| 116.Schneider 2023 | Traumatic Brain Injury and Long-Term Risk of Stroke Among US Military Veterans | Lack of relevant data: No mortality-specific data presented |
| 117.Schonberger 2009 | The Relationship between Age, Injury Severity, and MRI Findings after Traumatic Brain Injury | Lack of relevant data: No mortality-specific data presented |
| 118.Sedlak 2025 | Sex-stratified patterns in geriatric patients with mild traumatic brain injury and intracranial bleeding: a retrospective cohort study | No comparison of mortality and sex |
| 119.Shafi 2005 | Hypotension does not increase mortality in brain- injured patients more than it does in Non- Brain- Injured patients | No comparison of mortality and sex |
| 120.Shirvani 2020 | Predicting the quality of life of patients with mild traumatic brain injury: a study based on psychological variables | Lack of relevant data: No mortality-specific data presented |
| 121.Skandsen 2021 | Personal Factors Associated With Postconcussion Symptoms 3 Months After Mild Traumatic Brain Injury | Lack of relevant data: No mortality-specific data presented |
| 122.Spitz 2015 | Mortality following Traumatic Brain Injury Inpatient Rehabilitation | Injury severity not specified and/or considered |
| 123.Sutton 2019 | Neck Injury Comorbidity in Concussion-Related Emergency Department Visits: A Population-Based Study of Sex Differences Across the Life Span | Lack of relevant data: No mortality-specific data presented |
| 124.Svedung Wettervik 2022 | Females Exhibit Better Cerebral Pressure Autoregulation, Less Mitochondrial Dysfunction, and Reduced Excitotoxicity after Severe Traumatic Brain Injury | No comparison of mortality and sex with adjustments |
| 125.Takada 2016 | Social participation and quality-of-life of patients with traumatic brain injury living in the community: A mixed methods study | Lack of relevant data: No mortality-specific data presented |
| 126.Tawil 2007 | Posttraumatic Cerebral Infarction: Incidence, Outcome, and Risk Factors | No comparison of mortality and sex |

| | | |
|---|---|---|
| 127. Thompson 2012 | Prevalence of Comorbidity and its Association with Traumatic Brain Injury and Outcomes in Older Adults | No comparison of mortality and sex |
| 128. Thuss 2024 | Mild traumatic brain injury in older adults: recovery course and insights on early predictors of outcome | Lack of relevant data: No mortality-specific data presented |
| 129. Tsushima 2009 | Sex differences in the long-term neuropsychological outcome of mild traumatic brain injury | Lack of relevant data: No mortality-specific data presented |
| 130. Utomo 2009 | Predictors of in-hospital mortality and 6-month functional outcomes in older adults after moderate to severe traumatic brain injury | No comparison of mortality and sex |
| 131. van der Horn 2013 | Postconcussive Complaints, Anxiety, and Depression Related to Vocational Outcome in Minor to Severe Traumatic Brain Injury | Lack of relevant data: No mortality-specific data presented |
| 132. van Walsem 2020 | Trajectories of self-reported competency up to 10 years following moderate-to-severe traumatic brain injury | Lack of relevant data: No mortality-specific data presented |
| 133. Vikane 2016 | Predictors for Return to Work in Subjects with Mild Traumatic Brain Injury | Lack of relevant data: No mortality-specific data presented |
| 134. Wagner 2004 | Relationships between Cerebrospinal Fluid Markers of Excitotoxicity, Ischemia, and Oxidative Damage after Severe TBI: The Impact of Gender, Age, and Hypothermia | Lack of relevant data: No mortality-specific data presented |
| 135. Wagner 2007 | Sex and genetic associations with cerebrospinal fluid dopamine and metabolite production after severe traumatic brain injury | Lack of relevant data: No mortality-specific data presented |
| 136. Wang 2018 | Sex Differences in Abnormal Intrinsic Functional Connectivity After Acute Mild Traumatic Brain Injury | Lack of relevant data: No mortality-specific data presented |
| 137. Williamson 2016 | Trajectories of Life Satisfaction Over the First 10 Years After Traumatic Brain Injury: Race, Gender, and Functional Ability | Lack of relevant data: No mortality-specific data presented |
| 138. Yeung 2011 | Protection From Traumatic Brain Injury in Hormonally Active Women vs Men of a Similar Age | No comparison of mortality and sex with adjustments |
| 139. Yong 2025 | Sex differences in work-related traumatic brain injury: a concurrent mixed methods study employing the person-environment-occupation model | Lack of relevant data: No mortality-specific data presented |
| 140. Yu 2022 | Predicting Global Functional Outcomes Among Post-traumatic Epilepsy Patients After Moderate-to-Severe Traumatic Brain Injury: Development of a Prognostic Model | Lack of relevant data: No mortality-specific data presented |
| 141. Yue 2019 | Age and sex-mediated differences in six-month outcomes after mild traumatic brain injury in young adults: a TRACK-TBI study | Lack of relevant data: No mortality-specific data presented |

**S1 Table 3.** Quality assessment of observational studies using the NIH tool

| **Study** | **1** | **2** | **3** | **4** | **5** | **6** | **7** | **8** | **9** | **10** | **11** | **12** | **13** | **14** | **Score** | **Summary Score** |
|---|---|---|---|---|---|---|---|---|---|---|---|---|---|---|---|---|
| 1. Albrecht (2016) | Yes | Yes | No | Yes/Yes | No | Yes | NA | No | Yes | NA | Yes | NA | NA | Yes | 7/11 | + Good |
| 2. Baguley (2012) | Yes | Yes | NR | Yes/Yes | No | Yes | CD | Yes | Yes | No | Yes | NA | NA | Yes | 8/14 | + |

| | | | | | | | | | | | | | | | | |
|---|---|---|---|---|---|---|---|---|---|---|---|---|---|---|---|---|
| | | | | | | | | | | | | | | | | Good |
| 3. Beijer (2023) | Yes | Yes | NR | Yes/Yes | Yes | Yes | NA | Yes | Yes | NA | Yes | NA | NA | Yes | 9/11 | ++<br>Excellent |
| 4. Berry (2009) | Yes | Yes | NR | Yes/Yes | NR | Yes | NA | Yes | Yes | NA | Yes | NA | NA | Yes | 8/11 | +<br>Good |
| 5. Clond (2011) | Yes | Yes | NR | Yes/Yes | Yes | Yes | NA | No | Yes | NA | Yes | NA | NA | Yes | 8/11 | +<br>Good |
| 6. Davis (2006) | Yes | Yes | NR | Yes/Yes | No | Yes | NA | Yes | Yes | NA | Yes | NA | NA | Yes | 8/11 | +<br>Good |
| 7. De Guise (2014) | Yes | Yes | NR | Yes/Yes | Yes | Yes | NA | No | Yes | NA | Yes | NA | NA | Yes | 8/11 | +<br>Good |
| 8. Donohue (2007) | Yes | Yes | NR | Yes/Yes | Yes | Yes | CD | Yes | Yes | No | Yes | NA | NA | Yes | 9/14 | +<br>Good |
| 9. Fortuna (2008) | Yes | Yes | Yes | Yes/Yes | Yes | Yes | NA | Yes | Yes | NA | Yes | NA | NA | Yes | 10/11 | ++<br>Excellent |
| 10. Han (2017) | Yes | Yes | NR | Yes/Yes | No | Yes | CD | No | Yes | No | Yes | NA | NA | Yes | 7/14 | +<br>Good |
| 11. Harrison-Felix (2012) | Yes | Yes | NR | Yes/Yes | No | Yes | CD | Yes | Yes | Yes | Yes | NA | Yes | Yes | 10/14 | +<br>Good |
| 12. Hong (2020) | Yes | Yes | NR | Yes/Yes | Yes | Yes | NA | Yes | Yes | NA | Yes | NA | NA | Yes | 9/11 | ++<br>Excellent |
| 15. Hosomi (2021) | Yes | Yes | NR | Yes/Yes | Yes | Yes | NA | No | Yes | NA | Yes | NA | NA | Yes | 8/11 | +<br>Good |
| 16. Mikolic (2020) | Yes | Yes | NR | Yes/Yes | Yes | Yes | CD | Yes | Yes | NA | Yes | NA | Yes | Yes | 10/14 | +<br>Good |
| 17. Ng (2006) | Yes | Yes | NR | Yes/Yes | Yes | Yes | CD | Yes | Yes | No | Yes | NA | NA | Yes | 9/14 | +<br>Good |
| 18. Ottochian (2008) | Yes | Yes | NR | Yes/Yes | Yes | Yes | NA | Yes | Yes | NA | Yes | NA | NA | Yes | 9/11 | ++<br>Excellent |
| 19. Ponsford (2008) | Yes | Yes | No | Yes/Yes | Yes | Yes | CD | Yes | Yes | No | Yes | NA | Yes | Yes | 10/14 | +<br>Good |
| 20. Selassie (2011) | Yes | Yes | NR | Yes/Yes | No | Yes | NA | Yes | Yes | NA | Yes | NA | NA | Yes | 8/11 | +<br>Good |
| 21. Selassie (2014) | Yes | Yes | NR | Yes/Yes | No | Yes | CD | Yes | Yes | NA | Yes | NA | NA | Yes | 8/14 | +<br>Good |

| | | | | | | | | | | | | | | | |
|---|---|---|---|---|---|---|---|---|---|---|---|---|---|---|---|
| 22. Shandro (2008) | Yes | Yes | NR | Yes/Yes | No | Yes | CD | No | Yes | No | Yes | NA | NA | Yes | 7/14 | + Good |
| 23. Shibahashi (2017) | Yes | Yes | Yes | Yes/Yes | Yes | Yes | CD | No | Yes | No | Yes | NA | NA | Yes | 9/14 | + Good |

† CD: Cannot determine; NA: Not applicable; NR: Not reported

**Criteria:**

1. **1.** Was the research question or objective in this paper clearly stated?
2. **2.** Was the study population clearly specified and defined?
3. **3.** Was the participation rate of eligible persons at least 50%?
    - If a study has both cross-sectional analysis and longitudinal analysis, separate the participants into different cohorts (sub-cohorts): cross-sectional cohort and longitudinal cohorts
    - Then report the participation rate separately for each sub-cohort
4. Were all the subjects selected or recruited from the same or similar populations (including the same time period)? Were inclusion and exclusion criteria for being in the study prespecified and applied uniformly to all participants? \
5. Was a sample size justification, power description, or variance and effect estimates provided?
    - If one of them is mentioned, the answer will be Yes.
6. For the analyses in this paper, were the exposure(s) of interest measured prior to the outcome(s) being measured?
7. Was the timeframe sufficient so that one could reasonably expect to see an association between exposure and outcome if it existed?
8. For exposures that can vary in amount or level, did the study examine different levels of the exposure as related to the outcome (e.g., categories of exposure, or exposure measured as continuous variable)?
9. Were the exposure measures (independent variables) clearly defined, valid, reliable, and implemented consistently across all study participants?
10. Was the exposure(s) assessed more than once over time?
    - Discussed; TBI screening does not count for measurement as it is binary (positive vs negative), and the outcome of interest is TBI severity which is assessed differently.
    - The example of the exposure assessed more than once: multiple TBIs and each is assessed separately
11. Were the outcome measures (dependent variables) clearly defined, valid, reliable, and implemented consistently across all study participants?
12. Were the outcome assessors blinded to the exposure status of participants?
13. Was loss to follow-up after baseline 20% or less?
14. Were key potential confounding variables measured and adjusted statistically for their impact on the relationship between exposure(s) and outcome(s)?

**QUIPS Six Potential Sources of Bias:**

1. Study Design à 1,10, 12
2. Study Participation à 2,3,4
3. Study Attrition à 13
4. Associated Factors à 9
5. Outcome Measures/ Confounding Account à11,14
6. Analysis à 5,6,7,8

**Summary of Scoring:**

In addition to crude calculation of total number of biases present out of total number of possible, each study quality was summarized qualitatively into three groups:

(1) Excellent (++) when all or most of the quality were fulfilled (i.e. allowing one "CD" or "NR" based six potential sources of bias (i.e. study participation, design and attrition, associated factors, outcome measurements, confounding variables, and analyses).);
(2) Good (+) when half items of criteria were fulfilled;
(3) Fair (-) when less than half of items of criteria were fulfilled.

**S2 Figure 1.** Funnel plot and Egger's test of included studies of adjusted odds ratios for mortality in females compared with males following traumatic brain injury.

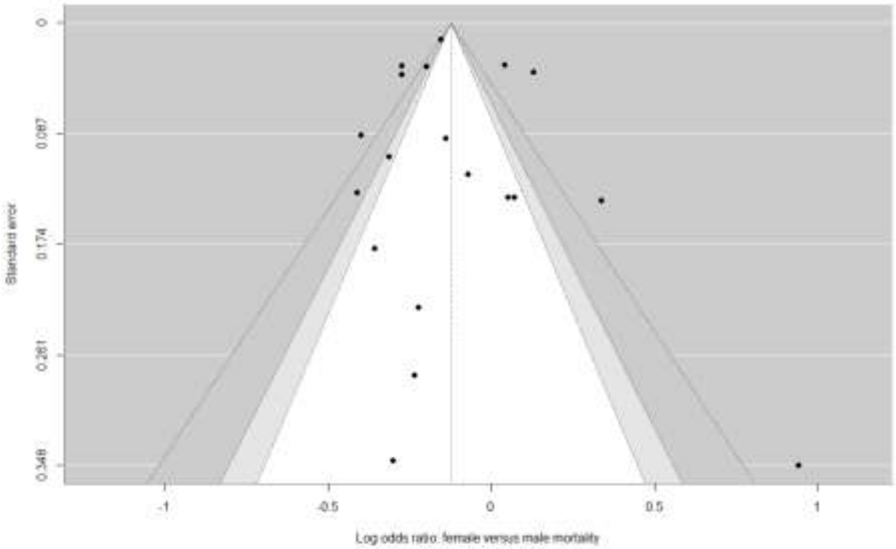

**S2 Figure 2.** Funnel plot and Egger's test of included studies of adjusted hazard ratios for mortality in females compared with males following traumatic brain injury.

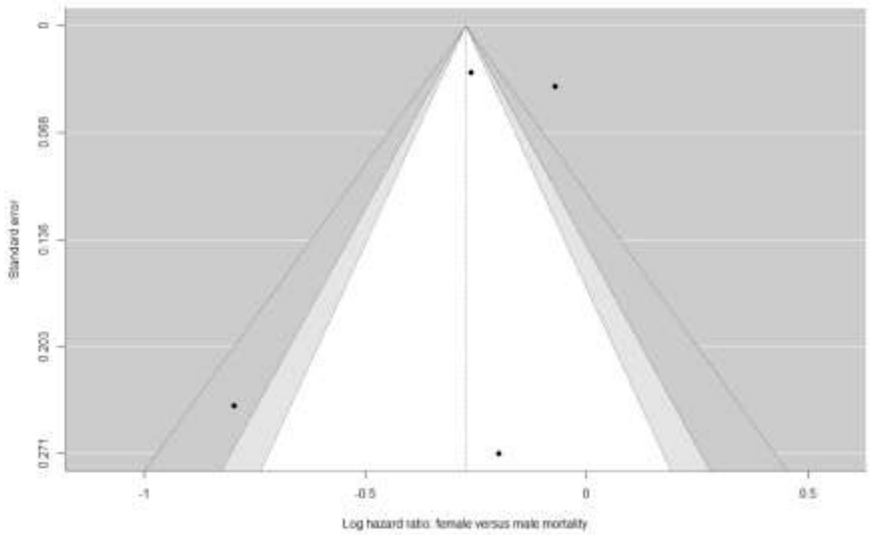

**S2 Figure 3.** Forest plots of pooled effects estimate of mortality after TBI by study quality, moderate and excellent quality groups, comparing females versus males.

Results are stratified by effect estimate: (A) adjusted odds ratios (B) and hazard ratios of mortality after TBI.

(A)

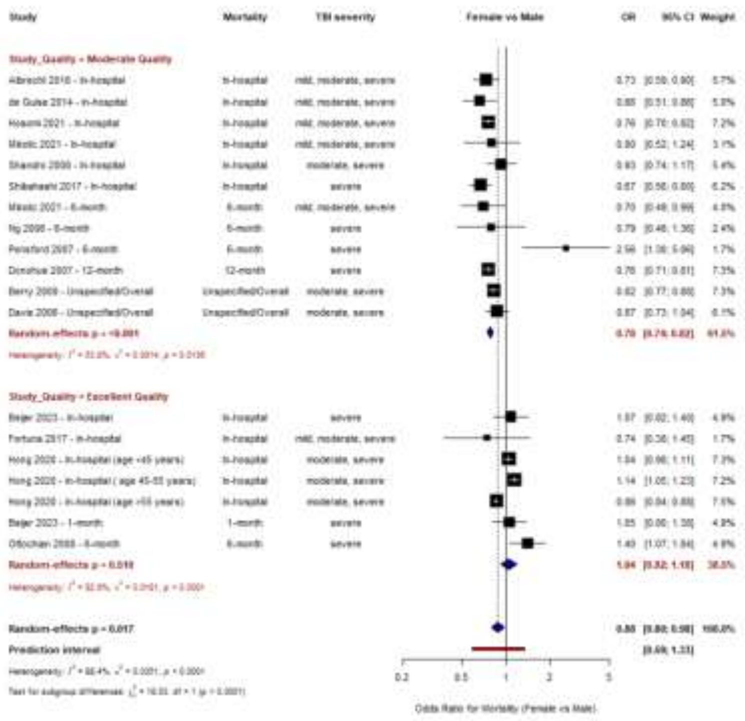

(B)

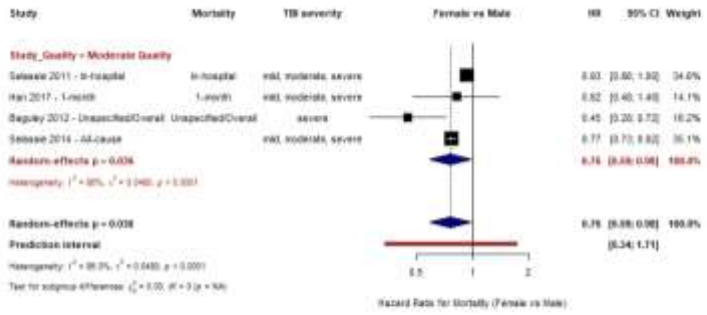
Study
Mortality
TBI severity
Female vs Male
HR
95% CI
Weight
Han 2017 - 1-month
1-month
mild, moderate, severe
0.82 [0.48; 1.40] 14.1%
Baguley 2012 - Unspecified/Overall
Unspecified/Overall
severe
0.45 [0.28; 0.72] 18.2%
Random-effects p = 0.038
Prediction interval
[0.34; 1.71]
0.5
1
2
Hazard Ratio for Mortality (Female vs Male)

**S2 Figure 4.** Forest plots of pooled effects estimate of mortality after TBI by sample size below and above the median sample size groups, comparing females versus males.

Results are stratified by effect estimate: (A) adjusted odds ratios (B) and hazard ratios of mortality after TBI.

(A)

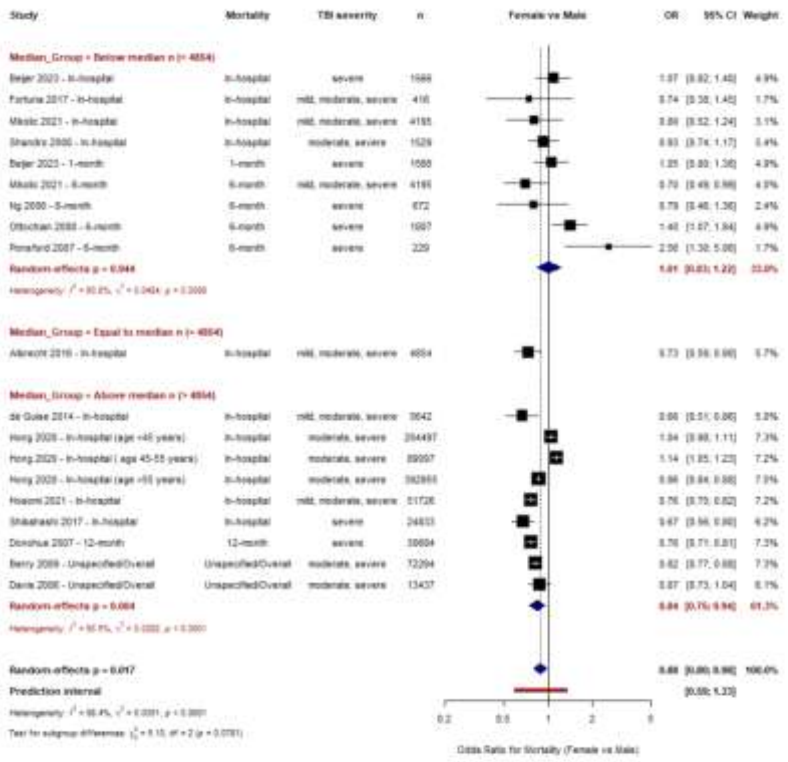

(B)

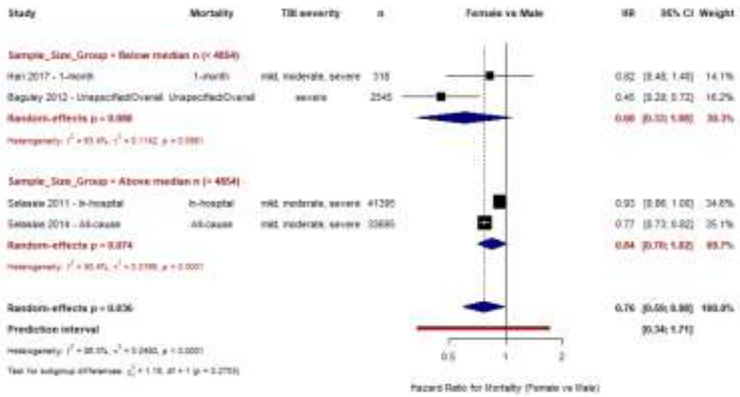
Study
Mortality
TBI severity
n
Female vs Male
HR
95% CI
Weight
Han 2017 - 1-month
1-month
mild, moderate, severe
318
0.82
[0.48; 1.40]
14.1%
Baguley 2012 - UnspecifiedOverall
UnspecifiedOverall
severe
2545
0.45
[0.28; 0.72]
16.2%
Selassie 2011 - In-hospital
In-hospital
mild, moderate, severe
41295
0.93
[0.86; 1.00]
34.6%
Selassie 2014 - All-cause
All-cause
mild, moderate, severe
33695
0.77
[0.73; 0.82]
35.1%
Prediction interval
[0.34; 1.71]
0.5
1
2
Hazard Ratio for Mortality (Female vs Male)

**S3 Table.** Performance of ML models for predicting modifiers of mortality in moderate-severe and severe TBI.

| Main Model | | | | |
|---|---|---|---|---|
| | MAE | | RMSE | |
| ML Model | Mod-sev | Severe | Mod-sev | Severe |
| Random Forest | 0.1655 | 0.0676 | 0.20627 | 0.07732 |
| Gradient Boosting | 0.11048 | 0.10329 | 0.1453 | 0.13794 |
| XGBoost | 0.17656 | 0.04185 | 0.21871 | 0.05113 |
| Main Model + Mean Sample Age and SD | | | | |
| | MAE | | RMSE | |
| ML Model | Mod-sev | Severe | Mod-sev | Severe |
| Random Forest | 0.1694 | 0.04733 | 0.21364 | 0.06642 |
| Gradient Boosting | 0.15823 | 0.11593 | 0.24956 | 0.19258 |
| XGBoost | 0.22827 | 0.04185 | 0.29374 | 0.05113 |
| Main Model + Region of Study Origin | | | | |
| | MAE | | RMSE | |
| ML Model | Mod-sev | Severe | Mod-sev | Severe |
| Random Forest | 0.17543 | 0.06721 | 0.2245 | 0.07645 |
| Gradient Boosting | 0.12379 | 0.10699 | 0.14476 | 0.17492 |
| XGBoost | 0.20237 | 0.04185 | 0.24881 | 0.05113 |
| Main Model + Median Sample Size | | | | |
| | MAE | | RMSE | |
| ML Model | Mod-sev | Severe | Mod-sev | Severe |
| Random Forest | 0.19092 | 0.03674 | 0.19092 | 0.04247 |
| Gradient Boosting | 0.15473 | 0.06166 | 0.15473 | 0.08263 |
| XGBoost | 0.206 | 0.04185 | 0.24893 | 0.05113 |

Abbreviations: MAE = mean absolute error; ML= machine learning; Mod-sev = moderate–severe; RMSE = root mean squared error; XGBoost = extreme gradient boosting

**S4 Figure 1.** Logarithmic effect estimates of mortality by injury severity for moderate-severe and severe TBI.

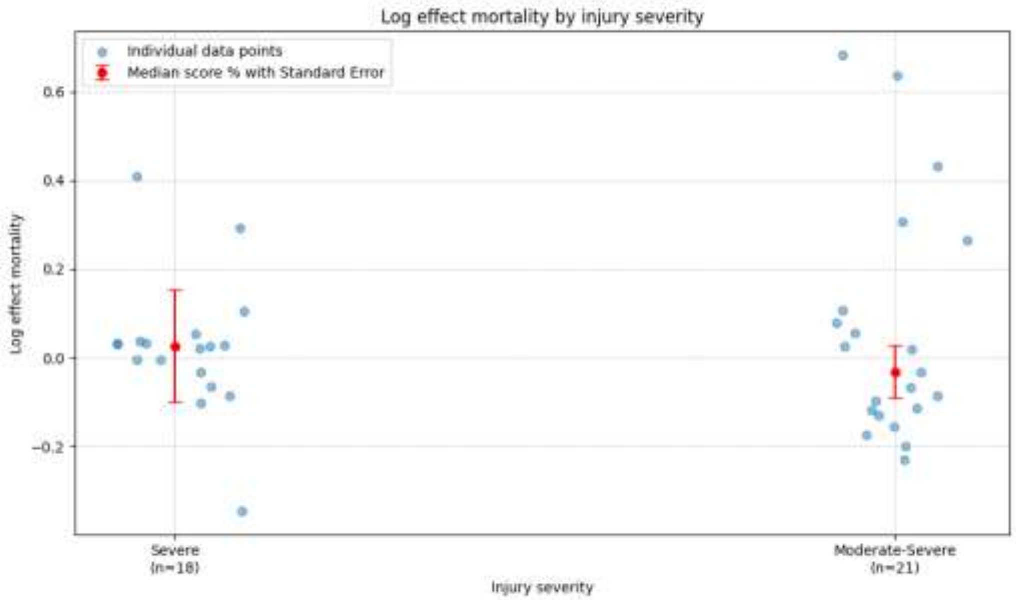

**S4 Figure 2.** Logarithmic effect estimates of mortality for moderate-severe and severe TBI for important predictors with available data: by men/males proportion by injury severity, mean age, and GII (a, b, c, respectively) and by mortality ascertainment period with sufficient data: in-hospital, 1-month, and unspecified/overall mortality (d, e, f, respectively).

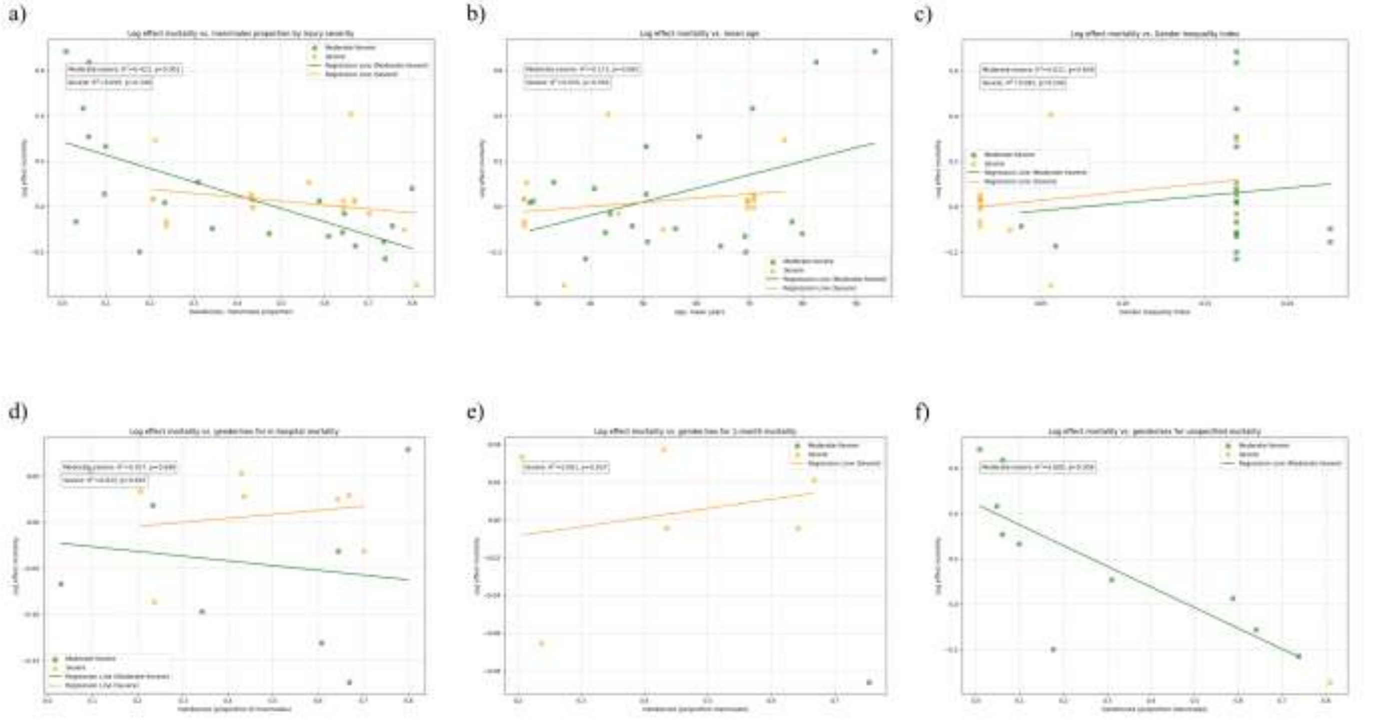

**S4 Figure 3.** Predictor correlation heatmaps for (a) moderate severe and (b) severe TBI cohorts showing correlation coefficients for associations between variables. Red signifies a positive correlation, while blue represents a negative correlation. The intensity of the color reflects the magnitude of the correlation coefficient, with more vibrant shades indicating stronger correlations. Specifically, shades tending towards red represent coefficients approaching 1, while those leaning towards blue represent coefficients approaching -1.

a)

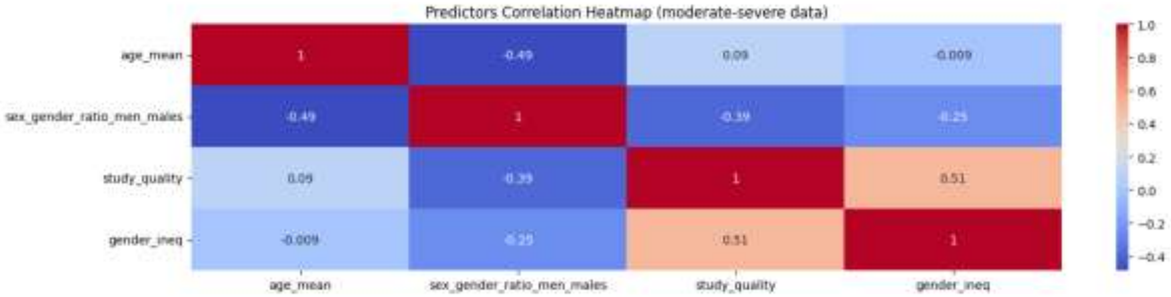

b)

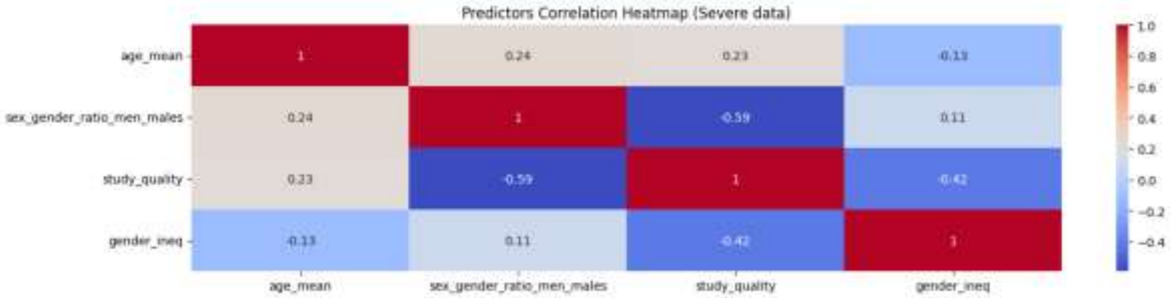

**S4 Figure 4.** Ranking of feature contributions for prediction of effect modifiers in mortality in (a) moderate-severe and (b) severe TBI cohorts, with age standard deviation (sd) added as a predictor. The colour scale reflects the strength of positive associations, with darker red indicating the strongest positive correlations and darker blue indicating the weakest positive correlations.

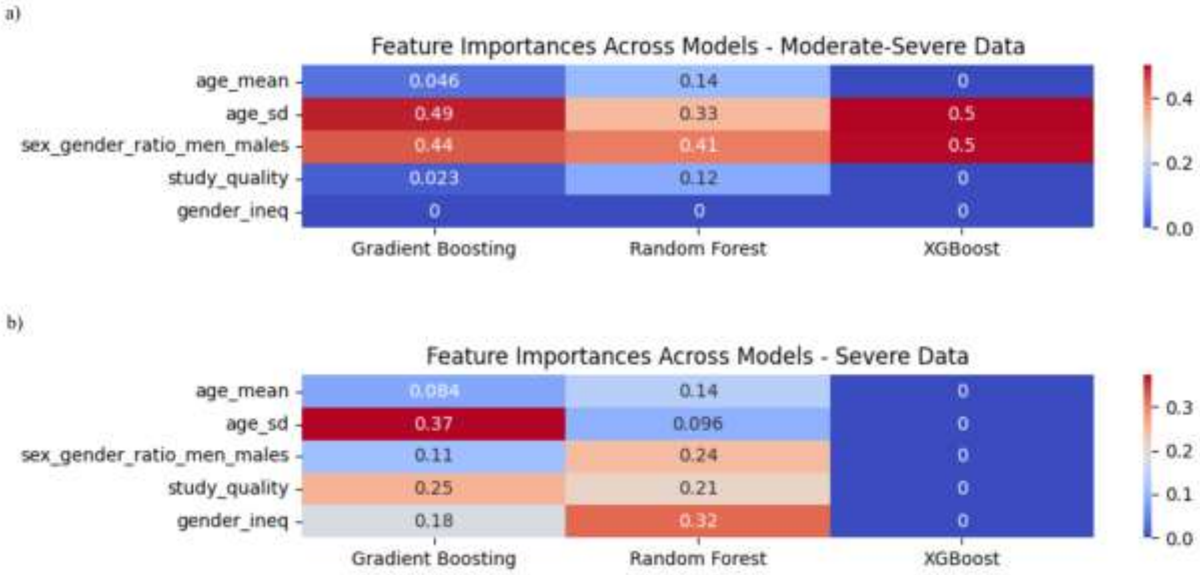

**Commented [PD1]:** age + sd

**S4 Figure 5.** SHAP summary plots illustrating the influence of input variables on mortality model predictions across models for (a) moderate-severe and (b) severe TBI cohorts, with age standard deviation (sd) added as a predictor. The colour of each point indicates the value of the feature, with red corresponding to higher values and blue to lower values. The position along the x-axis reflects whether the feature increases or decreases the prediction in the model output. Features are ranked on the y-axis based on their overall importance, with those contributing most to the model's predictions appearing at the top.

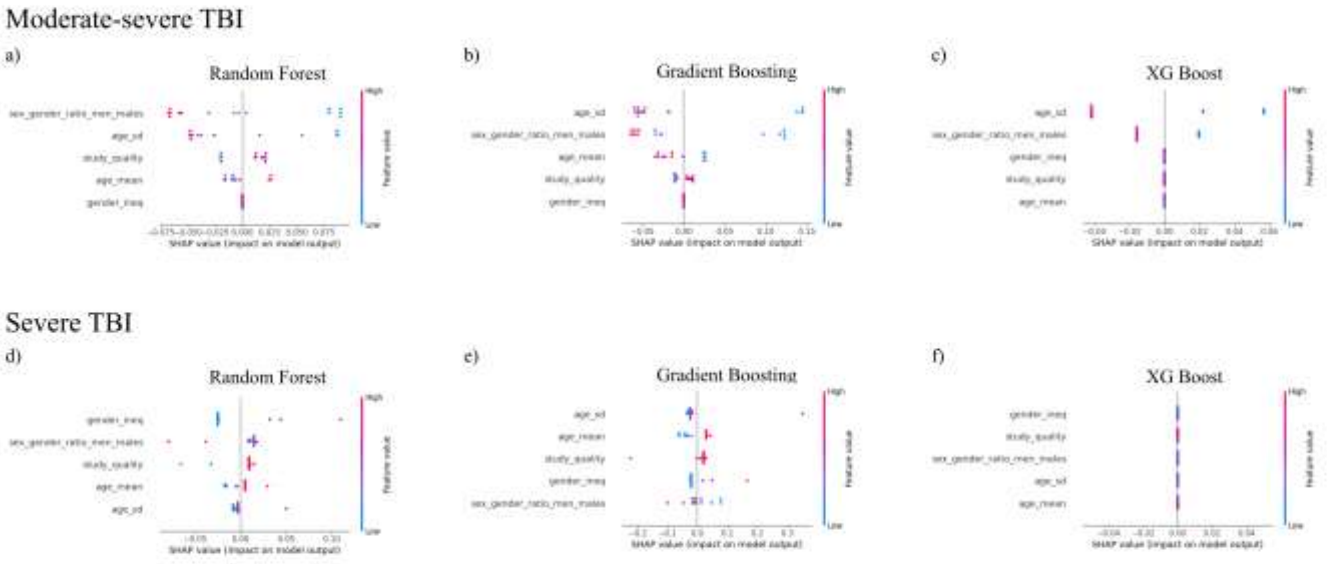

**S4 Figure 6.** Ranking of feature contributions for prediction of effect modifiers in mortality in (a) moderate-severe and (b) severe TBI cohorts, with region/country of study origin added as a predictor. The colour scale reflects the strength of positive associations, with darker red indicating the strongest positive correlations and darker blue indicating the weakest positive correlations.

Commented [PD2]: region

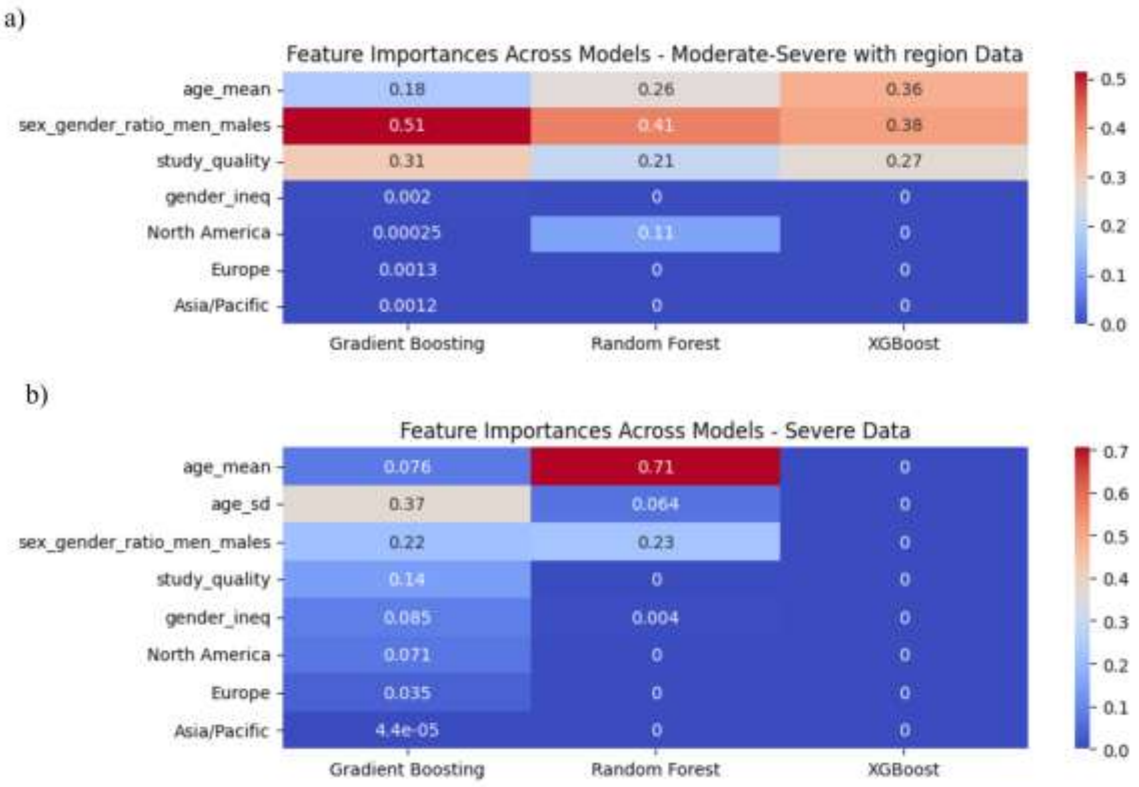

**S4 Figure 7.** SHAP summary plots illustrating the influence of input variables on mortality model predictions across models for (a) moderate-severe and (b) severe TBI cohorts, with region/country of study origin added as a predictor. The colour of each point indicates the value of the feature, with red corresponding to higher values and blue to lower values. The position along the x-axis reflects whether the feature increases or decreases the prediction in the model output. Features are ranked on the y-axis based on their overall importance, with those contributing most to the model's predictions appearing at the top.

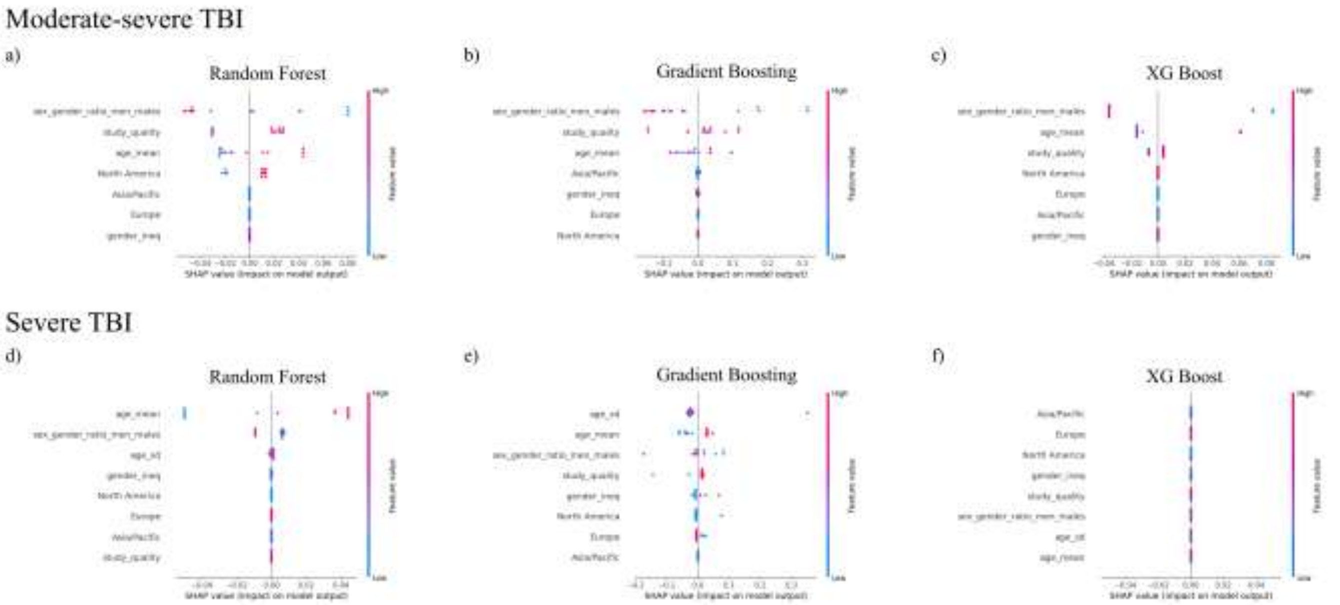

**S4 Figure 8.** Ranking of feature contributions for prediction of effect modifiers in mortality in (a) moderate-severe and (b) severe TBI cohorts, with sample size included in models. The colour scale reflects the strength of positive associations, with darker red indicating the strongest positive correlations and darker blue indicating the weakest positive correlations.

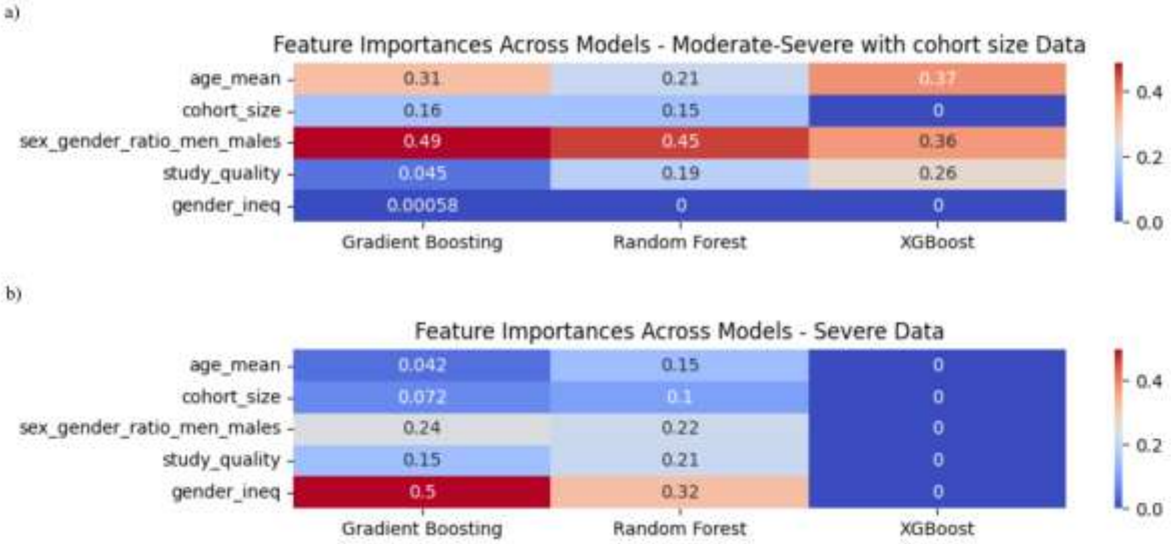

Commented [PD3]: sample size

**S4 Figure 9.** SHAP summary plots illustrating the influence of input variables on model predictions across models for (a) moderate-severe and (b) severe TBI cohorts, with sample size included in models. The colour of each point indicates the value of the feature, with red corresponding to higher values and blue to lower values. The position along the x-axis reflects whether the feature increases or decreases the prediction in the model output. Features are ranked on the y-axis based on their overall importance, with those contributing most to the model's predictions appearing at the top.

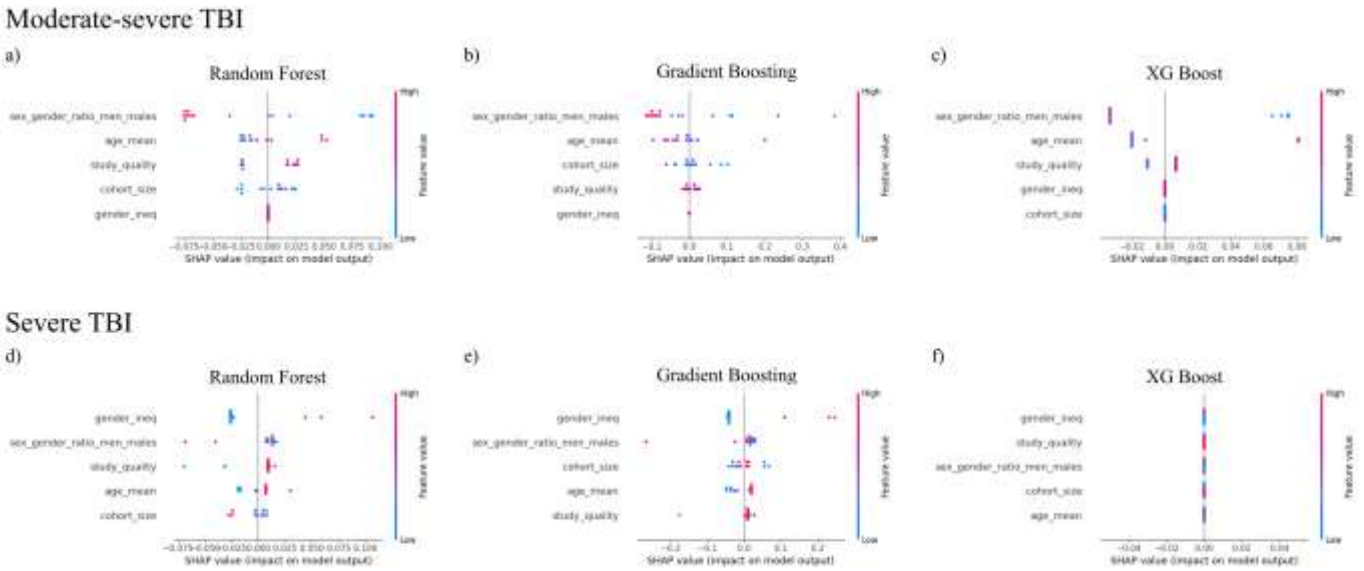

**S5 Results. Appendix**

PROGRESS-Plus parameters, including place of residence, race, occupation, socioeconomic status, social capital, and insurance status, were examined across seven studies (Figure 2) [49, 50, 51, 52, 53, 54, 55].

Place of residence

Place of residence was examined in one study [53]. Selassie et al. [53] reported that rural residence was associated with a greater mortality compared to urban residence (reference group); however, this association was not statistically significant (HR = 1.04, 95% CI: 0.97-1.11, $p = 0.321$).

Race, ethnicity, culture, and language

Race/ethnicity/culture/language were reported in seven studies [49, 50, 51, 52, 53, 54, 55]. The study by Albrecht et al. [49] found that Black individuals were associated with non-significant increased mortality following TBI (OR = 1.31, 95% CI: 0.76-2.24), while "other" races were associated with significant decreases in mortality (OR = 0.27, 95% CI: 0.10-0.77) compared to White individuals (reference group). De Guise et al. [50] reported that French speaking individuals were at lower odds of mortality compared to non-French speakers (OR = 0.87, 95% CI: 0.69–1.1, $p = 0.232$). Harrison-Felix et al. [51] reported Black (RR = 0.86, 95% CI: 0.69-1.08), Hispanic (RR = 0.61, 95% CI: 0.33-1.11), and Asian/Pacific Islander individuals (RR = 0.51, 95% CI: 0.33-0.81) were associated with non-statistically significant lower mortality compared to White individuals (reference group), but Indigenous individuals (RR = 0.87, 95% CI: 0.36-2.13) were associated with a statistically significant lower mortality compared to White individuals. Hong et al. [52] reported that Black individuals were significantly associated with higher mortality odds in the under 45-year-old age group (OR =

1.111, 95 % CI: 1.023–1.207, $p = 0.012$). The direction of association was supported in the 45–55-year-old age group, but was not significant (OR = 1.038, 95% CI: 0.937–1.15, $p = 0.477$). In the above 55-year-old age group, Black individuals were significantly associated with lower odds of mortality compared to White individuals (OR = 0.682, 95% CI: 0.645–0.721, $p < 0.001$). Asian individuals were found to be associated with greater mortality odds compared to White individuals in all age groups, but results were not significant (under 45 years: OR =1.166, 95% CI: 0.953–1.427, $p = 0.135$; 45-55 years: OR = 1.367, 95% CI: 1.068–1.751, $p = 0.013$; above 55 years: OR = 1.013, 95% CI: 0.936–1.096, $p = 0.744$). American Indian individuals were reported to be associated with lower mortality odds compared to White individuals, but results were only significant for the age group above 45 years old (under 45 years: OR = 0.901 95% CI: 0.729–1.115, $p = 0.339$; 45-55 years: OR = 0.788, 95% CI: 0.585–1.062, $p = 0.117$; 55 years: OR = 0.588, 95% CI: 0.479–0.721, $p < 0.001$). For individuals in the "other/unknown race" category, mortality odds were greater in the under 45-year-old and 45–55-year-old age groups but the above 55 year age group had significantly lower odds of mortality (under 45 years: OR = 1.056, 95% CI: 0.985–1.133, $p = 0.124$; 45-55 years: OR = 1.109, 95% CI: 1.009–1.219, $p = 0.032$; above 55 years: OR = 0.814, 95% CI: 0.78–0.85, $p < 0.001$). Selassie et al. [53] and Selassie et al. [54] reported that Black individuals ((HR = 0.98, 95% CI: 0.91-1.07, $p = 0.694$; HR = 0.99, 95% CI: 0.92-1.06, $p = 0.763$) were associated with lower mortality compared to White (reference group), but Sellasie et al. [54] reported a significant lower association of mortality in "other" (HR = 0.68, 95% CI: 0.55-0.83, $p < 0.001$) compared to White individuals. Shandro et al. [55] found that White (OR = 0.99, 95% CI: 0.77-1.29) and "other" races (OR = 0.84, 95% CI: 0.61-1.14) were associated with non-significantly lower mortality compared to Hispanic individuals (reference group).

Occupation status

Occupation status was examined in one study, Harrison-Felix et al. [51], which compared to competitively employed individuals (reference group), unemployed (RR = 1.52, 95% CI: 1.15-2.01) and retired individuals (RR = 1.72, 95% CI: 1.34-2.21) had a statistically significant increased risk of mortality. Students demonstrated a lower mortality risk (RR = 0.94, 95% CI: 0.48-1.86) compared to the reference group, which did not reach the level of statistical significance. Individuals classified as "other" (homemaker, volunteer, or other occupations) showed a statistically significant higher mortality risk (RR = 2.14, 95% CI: 15-3.07).

Social capital

Social capital was assessed in one study Harrison-Felix et al. [51] which compared to married individuals (reference group), divorced or widowed individuals demonstrated an increased mortality risk (RR = 1.35, 95% CI: 1.11-1.64), while individuals who were never married showed a decreased but non-significant mortality risk (RR = 0.91, 95% CI: 0.71-1.17).

Insurance status

Insurance status (Plus parameter) was examined across three studies and showed mixed associations with mortality risk [53, 54, 55]. Selassie et al. [53] and Selassie et al. [54] found that compared to individuals with commercial insurance (reference group), individuals who were uninsured/no had statistically significant higher mortality (HR = 1.29, 95% CI: 1.15-1.45, $p$ <0.001; HR = 1.27, 95% CI: 1.11-1.45, $p$ <0.001). Individuals who had Medicare had greater mortality in Selassie et al. [53] (HR 1.02, 95% CI 0.9-1.16, $p$ = 0.102), but was not statistically significant, and Selassie et al. [54] reported significantly greater mortality in Medicare groups (HR = 1.54, 95% CI: 1.4-1.69, $p$ <0.001). Individuals with indigent care/Medicaid were reported to have significantly lower mortality in

Selassie et al. [53] (HR = 0.89, 95% CI: 0.8-1.00, $p$ <0.041) and Selassie et al. [54] reported significantly greater mortality in Medicare groups (HR = 1.67, 95% CI: 1.48-1.87, $p$ <0.001). Shandro et al. [55] reported that Medicaid (OR = 3.48, 95% CI: 1.94-6.23), private (OR = 3.11, 95% CI: 2.06-4.7), and private + Medicare (OR = 1.92, 95% CI: 1.32-2.8) were associated with higher mortality compared to the reference group (uninsured/no insurance), Medicare only (OR = 1.27, 95% CI: 0.83-1.94) had a non-statistically significant higher mortality, while the “other” insurance category demonstrated non-statistically significant lower mortality (OR = 0.27, 95% CI: 0.1-0.73), compared to reference group.

**S6 Appendix. R codes used in analyses**

**Heatmap**

```
# ========================
# Install/load packages
# ========================
packages <- c("readxl", "dplyr", "tidyr", "ggplot2", "janitor", "stringr")
for (pkg in packages) {
if (!require(pkg, character.only = TRUE)) {
install.packages(pkg)
library(pkg, character.only = TRUE)
}
}
# ========================
# Load data
# ========================
```

```
df <- readxl::read_excel(file.choose())
df <- janitor::clean_names(df)
print(colnames(df))
# ========================
# Clean key text columns
# ========================
df <- df %>%
mutate(
mortality_outcome = str_trim(as.character(mortality_outcome)),
author = str_trim(as.character(author))
)
# ========================
# Recode mortality outcome labels
# This fixes NA rows by converting them to Unspecified/mixed
# ========================
df <- df %>%
mutate(
mortality_outcome = case_when(
is.na(mortality_outcome) ~ "Unspecified/mixed",
mortality_outcome == "" ~ "Unspecified/mixed",
mortality_outcome %in% c(
"NA",
"N/A",
"Overall",
"Overall/Unspecified",
"Unspecified",
"Mixed",
"Unspecified/Mixed",
"unspecified/mixed"
```

```
) ~ "Unspecified/mixed",
mortality_outcome == "1 year" ~ "1-year",
mortality_outcome == "2 year" ~ "2-year",
mortality_outcome == "1-year" ~ "1-year",
mortality_outcome == "2-year" ~ "2-year",
TRUE ~ mortality_outcome
)
)
# Check mortality outcome labels after cleaning
print(unique(df$mortality_outcome))
# ========================
# Optional: fix author labels
# This changes "Albrecht 2016" to "Albrecht et al. (2016)"
# ========================
df <- df %>%
mutate(
author_label = case_when(
str_detect(author, "\\(") ~ author,
TRUE ~ str_replace(author, "^(.*)\\s(\\d{4})$", "\\1 et al. (\\2)")
)
)
# ========================
# Mortality outcome order
# ========================
mortality_order <- c(
"In-hospital",
"1-month",
"3-month",
"6-month",
```

```
"12-month",
"24-month",
"1-year",
"2-year",
"Unspecified/Overall"
)
df <- df %>%
mutate(
mortality_outcome = factor(
mortality_outcome,
levels = mortality_order
)
)
# ========================
# Safety check for unmatched mortality outcomes
# ========================
if (any(is.na(df$mortality_outcome))) {
warning("Some mortality outcomes still did not match the mortality_order levels.")
print(df %>% filter(is.na(mortality_outcome)) %>% select(author, mortality_outcome))
}
# ========================
# Order rows
# ========================
df <- df %>%
arrange(mortality_outcome, author_label) %>%
mutate(
study = paste(author_label, mortality_outcome, sep = " - ")
)
df$study <- factor(df$study, levels = rev(unique(df$study)))
```

```
# ========================
# Heatmap variables
# ========================
heatmap_vars <- c(
"place_of_residence",
"race_ethnicity_language",
"occupation",
"gender_sex",
"religion",
"education",
"socioeconomic_status",
"social_capital",
"plus_age",
"plus_other",
"severity_mod_sev",
"severity_sev",
"severity_mixed",
"quality_moderate",
"quality_excellent",
"below_median_n",
"above_median_n"
)
# Check that all expected columns exist
missing_vars <- setdiff(heatmap_vars, colnames(df))
if (length(missing_vars) > 0) {
stop(
paste(
"These expected columns are missing from your dataset:",
paste(missing_vars, collapse = ", ")
```

```
)
)
}
# =========================
# Force heatmap columns to numeric
# This fixes pivot_longer type errors
# =========================
df <- df %>%
mutate(
across(
all_of(heatmap_vars),
~ as.numeric(as.character(.))
)
)
# =========================
# Pivot heatmap variables
# =========================
df_long <- df %>%
pivot_longer(
cols = all_of(heatmap_vars),
names_to = "variable",
values_to = "included"
)
# =========================
# Variable labels and order
# =========================
df_long <- df_long %>%
mutate(
variable = factor(
```

```
variable,
levels = heatmap_vars,
labels = c(
"Place of Residence",
"Race/Ethnicity/Language",
"Occupation",
"Gender/Sex",
"Religion",
"Education",
"Socioeconomic Status",
"Social Capital",
"Age",
"Other",
"Moderate-Severe TBI",
"Severe TBI",
"Mixed Severity",
"Moderate Quality",
"Excellent Quality",
"Below Median N",
"Above Median N"
)
)
)
# ========================
# Included labels
# ========================
df_long <- df_long %>%
mutate(
included_label = case_when(
```

```
included == 1 ~ "Yes",
included == 0 ~ "No",
TRUE ~ "No"
),
included_label = factor(included_label, levels = c("No", "Yes"))
)
# ========================
# Heatmap
# ========================
heatmap_plot <- ggplot(
df_long,
aes(x = variable, y = study, fill = included_label)
) +
geom_tile(color = "white", linewidth = 0.3) +
scale_fill_manual(
values = c(
"No" = "grey90",
"Yes" = "steelblue"
),
name = "Included"
) +
theme_minimal(base_size = 11) +
theme(
axis.text.x = element_text(
angle = 45,
hjust = 1,
vjust = 1,
size = 10
),
```

```
axis.text.y = element_text(size = 8),
axis.title.x = element_text(size = 12),
axis.title.y = element_text(size = 12),
plot.title = element_text(face = "bold", size = 16),
plot.subtitle = element_text(size = 12),
panel.grid = element_blank(),
legend.position = "right",
legend.title = element_text(size = 12),
legend.text = element_text(size = 11)
) +
labs(
title = "Study Characteristics and PROGRESS-Plus Variables",
subtitle = "Median sample size = 4854 (29118)",
x = "Variable",
y = "Study and Mortality Outcome"
)
print(heatmap_plot)
# ========================
# Save heatmap
# ========================
ggsave(
filename = "heatmap_study_characteristics_progress_plus.png",
plot = heatmap_plot,
width = 16,
height = 10,
dpi = 300
)
```

**Meta-analysis**
**Forest Plot by Mortality**

install.packages(c("readxl", "meta", "dplyr", "stringr"))
library(readxl) library(meta) library(dplyr) library(stringr)

-------------------------

Load data

-------------------------

df <- read_excel( file.choose(), sheet = "OR MA (sorted+adj) (3)" )

-------------------------

Clean data

-------------------------

ma_data <- df %>% mutate( Author = trimws(as.character(Author)), Author_Sort = tolower(Author), Study_Label = trimws(as.character(Study Label)), Mortality_Group = trimws(as.character(Mortality Group)), Severity = trimws(as.character(TBI Severity)),
OR = as.numeric(`Adjusted OR`),
lower = as.numeric(`CI Lower`),
upper = as.numeric(`CI Upper`)

) %>% filter( !is.na(OR), !is.na(lower), !is.na(upper), OR > 0, lower > 0, upper > 0 ) %>% mutate( logOR = log(OR), SE = (log(upper) - log(lower)) / (2 * 1.96) )

-------------------------

Standardize mortality labels

-------------------------

ma_data <- ma_data %>% mutate( Mortality_Group = case_when( Mortality_Group %in% c("In hospital", "In-hospital mortality") ~ "In-hospital", Mortality_Group %in% c("30-day", "1-month mortality") ~ "1-month", Mortality_Group %in% c("6 month", "6-month

```
mortality") ~ "6-month", Mortality_Group %in% c("12 month", "12-month mortality") ~ "12-month", Mortality_Group %in% c("Unspecified", "Overall", "Unspecified/Overall") ~ "Unspecified/Overall", TRUE ~ Mortality_Group ) )
-------------------------
Order mortality groups
-------------------------
ma_data$Mortality_Group <- factor( ma_data$Mortality_Group, levels = c( "In-hospital", "1-month", "3-month", "6-month", "12-month", "Unspecified/Overall", "Unspecified/Mixed" ) )
-------------------------
Sort:
1. Mortality type
2. Author alphabetically, ignoring case
-------------------------
ma_data <- ma_data %>% arrange( Mortality_Group, Author_Sort )
-------------------------
Random-effects meta-analysis grouped by mortality type
-------------------------
meta_mortality <- metagen( TE = logOR, seTE = SE, studlab = Study_Label, data = ma_data, sm = "OR", common = FALSE, random = TRUE, method.tau = "REML", subgroup = Mortality_Group )
summary(meta_mortality)
-------------------------
Random-effects p-value labels
-------------------------
subgroup_p_text <- paste0( "Random-effects p = ", ifelse( meta_mortality$pval.random.w < 0.001, "<0.001", sprintf("%.3f", meta_mortality$pval.random.w) ) )
overall_p_text <- paste0( "Random-effects p = ", ifelse( meta_mortality$pval.random < 0.001, "<0.001", sprintf("%.3f", meta_mortality$pval.random) ) )
```

```
-------------------------
Save forest plot as PDF
-------------------------
pdf( "forest_plot_TBI_by_mortality_type_alphabetical_fixed.pdf", width = 16, height = 22 )
forest( meta_mortality,
prediction = TRUE, print.subgroup.labels = TRUE,
print.I2 = TRUE, print.tau2 = TRUE, print.pval.Q = TRUE, test.subgroup = TRUE,
leftcols = c("studlab", "Severity"), leftlabs = c("Study", "TBI severity"),
rightcols = c("effect", "ci", "w.random"), rightlabs = c("OR", "95% CI", "Weight"),
xlab = "Odds Ratio for Mortality (Female vs Male)", smlab = "Female vs Male",
text.random.w = subgroup_p_text, text.random = overall_p_text,
backtransf = TRUE,
col.square = "black", col.study = "black", col.diamond = "darkblue", col.diamond.lines = "black", col.subgroup = "darkred",
fontsize = 8, spacing = 1.2, digits = 2 )
dev.off()
-------------------------
Display forest plot in R
-------------------------
forest( meta_mortality,
prediction = TRUE, print.subgroup.labels = TRUE,
print.I2 = TRUE, print.tau2 = TRUE, print.pval.Q = TRUE, test.subgroup = TRUE,
leftcols = c("studlab", "Severity"), leftlabs = c("Study", "TBI severity"),
rightcols = c("effect", "ci", "w.random"), rightlabs = c("OR", "95% CI", "Weight"),
xlab = "Odds Ratio for Mortality (Female vs Male)", smlab = "Female vs Male",
text.random.w = subgroup_p_text, text.random = overall_p_text,
backtransf = TRUE,
col.square = "black", col.study = "black", col.diamond = "darkblue", col.diamond.lines = "black", col.subgroup = "darkred",
fontsize = 8, spacing = 1.2, digits = 2 )
cat( "\nForest plot saved as:\n", "forest_plot_TBI_by_mortality_type_alphabetical_fixed.pdf\n" )
```

**Forest Plot by Quality**

```
install.packages(c("readxl", "meta", "dplyr"))

library(readxl)
library(meta)
library(dplyr)

df <- read_excel(
  file.choose(),
  sheet = "OR MA (sample & study quality)"
)

ma_data <- df %>%
  mutate(
    Author = trimws(as.character(Author)),
    Author_Sort = tolower(Author),
    Study_Label = trimws(as.character(`Study Label`)),
    Study_Quality = trimws(as.character(`Study Quality`)),
    Mortality_Group = trimws(as.character(`Mortality Group`)),
    Severity = trimws(as.character(`TBI Severity`)),

    OR = as.numeric(`Adjusted OR`),
    lower = as.numeric(`CI Lower`),
```

```
  upper = as.numeric(`CI Upper`)
 ) %>%
 mutate(
  Mortality_Group = case_when(
   Mortality_Group %in% c("In hospital", "In-hospital mortality") ~ "In-hospital",
   Mortality_Group %in% c("30-day", "1-month mortality") ~ "1-month",
   Mortality_Group %in% c("6 month", "6-month mortality") ~ "6-month",
   Mortality_Group %in% c("12 month", "12-month mortality") ~ "12-month",
   Mortality_Group %in% c("Unspecified", "Overall", "Unspecified/Overall") ~ "Unspecified/Overall",
   TRUE ~ Mortality_Group
  )
 ) %>%
 filter(
  !is.na(OR),
  !is.na(lower),
  !is.na(upper),
  OR > 0,
  lower > 0,
  upper > 0
 ) %>%
 mutate(
  logOR = log(OR),
  SE = (log(upper) - log(lower)) / (2 * 1.96)
 )

ma_data$Study_Quality <- factor(
 ma_data$Study_Quality,
 levels = c(
  "Moderate Quality",
```

```
    "Excellent Quality"
  )
)

ma_data$Mortality_Group <- factor(
  ma_data$Mortality_Group,
  levels = c(
    "In-hospital",
    "1-month",
    "3-month",
    "6-month",
    "12-month",
    "Unspecified/Overall",
    "Unspecified/Mixed"
  )
)

ma_data <- ma_data %>%
  arrange(
    Study_Quality,
    Mortality_Group,
    Author_Sort
  )

meta_or_quality <- metagen(
  TE = logOR,
  seTE = SE,
  studlab = Study_Label,
  data = ma_data,
```

```
  sm = "OR",
  common = FALSE,
  random = TRUE,
  method.tau = "REML",
  subgroup = Study_Quality
)

summary(meta_or_quality)

subgroup_p_text <- paste0(
  "Random-effects p = ",
  ifelse(
    meta_or_quality$pval.random.w < 0.001,
    "<0.001",
    sprintf("%.3f", meta_or_quality$pval.random.w)
  )
)

overall_p_text <- paste0(
  "Random-effects p = ",
  ifelse(
    meta_or_quality$pval.random < 0.001,
    "<0.001",
    sprintf("%.3f", meta_or_quality$pval.random)
  )
)

pdf(
  "forest_plot_OR_by_study_quality_fixed.pdf",
```

```
  width = 16,
  height = 22
)

forest(
  meta_or_quality,

  prediction = TRUE,
  print.subgroup.labels = TRUE,
  print.I2 = TRUE,
  print.tau2 = TRUE,
  print.pval.Q = TRUE,
  test.subgroup = TRUE,

  leftcols = c("studlab", "Mortality_Group", "Severity"),
  leftlabs = c("Study", "Mortality", "TBI severity"),

  rightcols = c("effect", "ci", "w.random"),
  rightlabs = c("OR", "95% CI", "Weight"),

  xlab = "Odds Ratio for Mortality (Female vs Male)",
  smlab = "Female vs Male",

  text.random.w = subgroup_p_text,
  text.random = overall_p_text,

  backtransf = TRUE,

  col.square = "black",
```

```
  col.study = "black",
  col.diamond = "darkblue",
  col.diamond.lines = "black",
  col.subgroup = "darkred",

  fontsize = 8,
  spacing = 1.2,
  digits = 2
)

dev.off()

forest(
  meta_or_quality,

  prediction = TRUE,
  print.subgroup.labels = TRUE,
  print.I2 = TRUE,
  print.tau2 = TRUE,
  print.pval.Q = TRUE,
  test.subgroup = TRUE,

  leftcols = c("studlab", "Mortality_Group", "Severity"),
  leftlabs = c("Study", "Mortality", "TBI severity"),

  rightcols = c("effect", "ci", "w.random"),
  rightlabs = c("OR", "95% CI", "Weight"),

  xlab = "Odds Ratio for Mortality (Female vs Male)",
```

```
  smlab = "Female vs Male",

  text.random.w = subgroup_p_text,
  text.random = overall_p_text,

  backtransf = TRUE,

  col.square = "black",
  col.study = "black",
  col.diamond = "darkblue",
  col.diamond.lines = "black",
  col.subgroup = "darkred",

  fontsize = 8,
  spacing = 1.2,
  digits = 2
)
```

**Forest Plot by Sample**

```
install.packages(c("readxl", "meta", "dplyr"))

library(readxl)
library(meta)
library(dplyr)

df <- read_excel(
  file.choose(),
  sheet = "OR MA (sample & study quality)"
)

ma_data <- df %>%
  mutate(
    Author = trimws(as.character(Author)),
    Author_Sort = tolower(Author),
    Study_Label = trimws(as.character(`Study Label`)),
    Mortality_Group = trimws(as.character(`Mortality Group`)),
    Severity = trimws(as.character(`TBI Severity`)),

    n_total = as.numeric(`n total`),

    OR = as.numeric(`Adjusted OR`),
    lower = as.numeric(`CI Lower`),
    upper = as.numeric(`CI Upper`)
  ) %>%
  mutate(
```

```
    Mortality_Group = case_when(
      Mortality_Group %in% c("In hospital", "In-hospital mortality") ~ "In-hospital",
      Mortality_Group %in% c("30-day", "1-month mortality") ~ "1-month",
      Mortality_Group %in% c("6 month", "6-month mortality") ~ "6-month",
      Mortality_Group %in% c("12 month", "12-month mortality") ~ "12-month",
      Mortality_Group %in% c("Unspecified", "Overall", "Unspecified/Overall") ~ "Unspecified/Overall",
      TRUE ~ Mortality_Group
    )
  ) %>%
  filter(
    !is.na(OR),
    !is.na(lower),
    !is.na(upper),
    !is.na(n_total),
    OR > 0,
    lower > 0,
    upper > 0,
    n_total > 0
  ) %>%
  mutate(
    logOR = log(OR),
    SE = (log(upper) - log(lower)) / (2 * 1.96)
  )

# Calculate median n from included studies
median_n <- median(ma_data$n_total, na.rm = TRUE)

# Create median subgroup directly from n
ma_data <- ma_data %>%
```

```
  mutate(
    Median_Group = case_when(
      n_total < median_n ~ paste0("Below median n (< ", median_n, ")"),
      n_total > median_n ~ paste0("Above median n (> ", median_n, ")"),
      n_total == median_n ~ paste0("Equal to median n (= ", median_n, ")"),
      TRUE ~ "Missing n"
    )
  ) %>%
  filter(Median_Group != "Missing n")

ma_data$Median_Group <- factor(
  ma_data$Median_Group,
  levels = c(
    paste0("Below median n (< ", median_n, ")"),
    paste0("Equal to median n (= ", median_n, ")"),
    paste0("Above median n (> ", median_n, ")")
  )
)

ma_data$Mortality_Group <- factor(
  ma_data$Mortality_Group,
  levels = c(
    "In-hospital",
    "1-month",
    "3-month",
    "6-month",
    "12-month",
    "Unspecified/Overall",
    "Unspecified/Mixed"
```

```
  )
)

ma_data <- ma_data %>%
  arrange(
    Median_Group,
    Mortality_Group,
    Author_Sort
  )

print(table(ma_data$Median_Group, useNA = "ifany"))

meta_or_sample <- metagen(
  TE = logOR,
  seTE = SE,
  studlab = Study_Label,
  data = ma_data,
  sm = "OR",
  common = FALSE,
  random = TRUE,
  method.tau = "REML",
  subgroup = Median_Group
)

summary(meta_or_sample)

subgroup_p_text <- paste0(
  "Random-effects p = ",
  ifelse(
```

```
    meta_or_sample$pval.random.w < 0.001,
    "<0.001",
    sprintf("%.3f", meta_or_sample$pval.random.w)
  )
)

overall_p_text <- paste0(
  "Random-effects p = ",
  ifelse(
    meta_or_sample$pval.random < 0.001,
    "<0.001",
    sprintf("%.3f", meta_or_sample$pval.random)
  )
)

pdf(
  "forest_plot_OR_by_median_sample_size_fixed.pdf",
  width = 16,
  height = 22
)

forest(
  meta_or_sample,

  prediction = TRUE,
  print.subgroup.labels = TRUE,
  print.I2 = TRUE,
  print.tau2 = TRUE,
  print.pval.Q = TRUE,
```

```
  test.subgroup = TRUE,

  leftcols = c("studlab", "Mortality_Group", "Severity", "n_total"),
  leftlabs = c("Study", "Mortality", "TBI severity", "n"),

  rightcols = c("effect", "ci", "w.random"),
  rightlabs = c("OR", "95% CI", "Weight"),

  xlab = "Odds Ratio for Mortality (Female vs Male)",
  smlab = "Female vs Male",

  text.random.w = subgroup_p_text,
  text.random = overall_p_text,

  backtransf = TRUE,

  col.square = "black",
  col.study = "black",
  col.diamond = "darkblue",
  col.diamond.lines = "black",
  col.subgroup = "darkred",

  fontsize = 8,
  spacing = 1.2,
  digits = 2
)

dev.off()
```

```
forest(
  meta_or_sample,

  prediction = TRUE,
  print.subgroup.labels = TRUE,
  print.I2 = TRUE,
  print.tau2 = TRUE,
  print.pval.Q = TRUE,
  test.subgroup = TRUE,

  leftcols = c("studlab", "Mortality_Group", "Severity", "n_total"),
  leftlabs = c("Study", "Mortality", "TBI severity", "n"),

  rightcols = c("effect", "ci", "w.random"),
  rightlabs = c("OR", "95% CI", "Weight"),

  xlab = "Odds Ratio for Mortality (Female vs Male)",
  smlab = "Female vs Male",

  text.random.w = subgroup_p_text,
  text.random = overall_p_text,

  backtransf = TRUE,

  col.square = "black",
  col.study = "black",
  col.diamond = "darkblue",
  col.diamond.lines = "black",
  col.subgroup = "darkred",
```

```
  fontsize = 8,
  spacing = 1.2,
  digits = 2
)

cat("\nMedian n =", median_n, "\n")
```

**Forest Plot by Severity**

```
install.packages(c("readxl", "meta", "dplyr", "stringr"))

library(readxl)
library(meta)
library(dplyr)
library(stringr)

df <- read_excel(
  file.choose(),
  sheet = "OR MA (sorted+adj) (3)"
)

ma_data <- df %>%
  mutate(
    Author = trimws(as.character(Author)),
    Author_Sort = tolower(Author),
    Study_Label = trimws(as.character(`Study Label`)),
    Mortality_Group = trimws(as.character(`Mortality Group`)),
    Severity = str_to_lower(trimws(as.character(`TBI Severity`))),
```

```
  OR = as.numeric(`Adjusted OR`),
  lower = as.numeric(`CI Lower`),
  upper = as.numeric(`CI Upper`)
) %>%
filter(
  !is.na(OR),
  !is.na(lower),
  !is.na(upper),
  OR > 0,
  lower > 0,
  upper > 0
) %>%
mutate(
  logOR = log(OR),
  SE = (log(upper) - log(lower)) / (2 * 1.96),
  Severity_Group = case_when(
    str_detect(Severity, "moderate") &
      str_detect(Severity, "severe") &
      !str_detect(Severity, "mild") ~ "Moderate + severe",

    str_detect(Severity, "mild") &
      str_detect(Severity, "moderate") &
      str_detect(Severity, "severe") ~ "Mild + moderate + severe",

    str_detect(Severity, "severe") &
      !str_detect(Severity, "mild|moderate") ~ "Severe only",

    TRUE ~ "Unspecified"
  )
```

```
  )

ma_data$Severity_Group <- factor(
  ma_data$Severity_Group,
  levels = c(
    "Severe only",
    "Moderate + severe",
    "Mild + moderate + severe",
    "Unspecified"
  )
)

ma_data$Mortality_Group <- factor(
  ma_data$Mortality_Group,
  levels = c(
    "In-hospital",
    "1-month",
    "3-month",
    "6-month",
    "12-month",
    "Unspecified/Overall",
    "Unspecified/Mixed"
  )
)

ma_data <- ma_data %>%
  arrange(
    Severity_Group,
    Mortality_Group,
```

```
    Author_Sort
  )

meta_severity <- metagen(
  TE = logOR,
  seTE = SE,
  studlab = Study_Label,
  data = ma_data,
  sm = "OR",
  common = FALSE,
  random = TRUE,
  method.tau = "REML",
  subgroup = Severity_Group
)

summary(meta_severity)

subgroup_p_text <- paste0(
  "Random-effects p = ",
  ifelse(
    meta_severity$pval.random.w < 0.001,
    "<0.001",
    sprintf("%.3f", meta_severity$pval.random.w)
  )
)

overall_p_text <- paste0(
  "Random-effects p = ",
  ifelse(
```

```
    meta_severity$pval.random < 0.001,
    "<0.001",
    sprintf("%.3f", meta_severity$pval.random)
  )
)

pdf(
  "forest_plot_TBI_by_severity_alphabetical_fixed.pdf",
  width = 16,
  height = 22
)

forest(
  meta_severity,
  prediction = TRUE,
  print.subgroup.labels = TRUE,
  print.I2 = TRUE,
  print.tau2 = TRUE,
  print.pval.Q = TRUE,
  test.subgroup = TRUE,

  leftcols = c("studlab", "Mortality_Group"),
  leftlabs = c("Study", "Mortality"),

  rightcols = c("effect", "ci", "w.random"),
  rightlabs = c("OR", "95% CI", "Weight"),

  xlab = "Odds Ratio for Mortality (Female vs Male)",
  smlab = "Female vs Male",
```

```
  text.random.w = subgroup_p_text,
  text.random = overall_p_text,

  backtransf = TRUE,

  col.square = "black",
  col.study = "black",
  col.diamond = "darkblue",
  col.diamond.lines = "black",
  col.subgroup = "darkred",

  fontsize = 8,
  spacing = 1.2,
  digits = 2
)

dev.off()

forest(
  meta_severity,
  prediction = TRUE,
  print.subgroup.labels = TRUE,
  print.I2 = TRUE,
  print.tau2 = TRUE,
  print.pval.Q = TRUE,
  test.subgroup = TRUE,

  leftcols = c("studlab", "Mortality_Group"),
```

```
  leftlabs = c("Study", "Mortality"),

  rightcols = c("effect", "ci", "w.random"),
  rightlabs = c("OR", "95% CI", "Weight"),

  xlab = "Odds Ratio for Mortality (Female vs Male)",
  smlab = "Female vs Male",

  text.random.w = subgroup_p_text,
  text.random = overall_p_text,

  backtransf = TRUE,

  col.square = "black",
  col.study = "black",
  col.diamond = "darkblue",
  col.diamond.lines = "black",
  col.subgroup = "darkred",

  fontsize = 8,
  spacing = 1.2,
  digits = 2
)
```

**Funnel Plots and Egger Test**

Install/load packages

packages <- c("readxl", "dplyr", "metafor")

for (pkg in packages) { if (!require(pkg, character.only = TRUE)) { install.packages(pkg) library(pkg, character.only = TRUE) } }

=========================

Load data

=========================

dat <- readxl::read_excel(file.choose())

Check column names

print(colnames(dat))

=========================

Prepare data

=========================

Assumes your columns are:

Author, Study Label, Adjusted OR, CI Lower, CI Upper, Ref Group

dat_or <- dat %>% mutate( or_original = as.numeric(Adjusted OR), ci_lower_original = as.numeric(CI Lower), ci_upper_original = as.numeric(CI Upper),

### If any studies still use female as the reference group,

### this converts them so the effect is female versus male, with male as reference.

OR = ifelse(`Ref Group` == "F", 1 / or_original, or_original),

CI_lower = ifelse(`Ref Group` == "F", 1 / ci_upper_original, ci_lower_original),

CI_upper = ifelse(`Ref Group` == "F", 1 / ci_lower_original, ci_upper_original),

```
yi = log(OR),
sei = (log(CI_upper) - log(CI_lower)) / (2 * 1.96)

) %>% filter( !is.na(yi), !is.na(sei), sei > 0 )
=========================
Random-effects meta-analysis
=========================
res_or <- rma( yi = yi, sei = sei, data = dat_or, method = "REML" )
summary(res_or)
=========================
Funnel plot on log OR scale
=========================
funnel( res_or, xlab = "Log odds ratio: female versus male mortality", ylab = "Standard error", refline = coef(res_or) )
=========================
Funnel plot back-transformed to OR scale
=========================
funnel( res_or, atransf = exp, xlab = "Odds ratio: female versus male mortality", ylab = "Standard error", refline = coef(res_or), at =
log(c(0.5, 0.75, 1, 1.25, 1.5, 2)) )
=========================
Funnel plot with study labels
=========================
funnel( res_or, xlab = "Log odds ratio: female versus male mortality", ylab = "Standard error", refline = coef(res_or), label = TRUE,
slab = dat_or$Study Label )
=========================
Contour-enhanced funnel plot
=========================
funnel( res_or, xlab = "Log odds ratio: female versus male mortality", ylab = "Standard error", refline = coef(res_or), level = c(90, 95,
99), shade = c("white", "gray90", "gray80") )
```

========================
Egger's test
========================

```
egger_or <- regtest( res_or, model = "rma", predictor = "sei" )
egger_or
```

To extract only the p-value:

```
egger_or$pval
```

**S7 Appendix. Python code machine learning models**

**1. Mounting dataset**

```
from google.colab import drive
drive.mount('/content/drive')
import pandas as pd
import re
import matplotlib.pyplot as plt
import seaborn as sns
import numpy as np
from mpl_toolkits.mplot3d import Axes3D

import statsmodels.stats.meta_analysis as anova
from sklearn.decomposition import PCA
from sklearn.preprocessing import StandardScaler,LabelEncoder,OneHotEncoder
from statsmodels.stats.outliers_influence import variance_inflation_factor
from statsmodels.tools.tools import add_constant
from scipy.stats.mstats import winsorize
from scipy.stats import randint, uniform
from sklearn.utils.class_weight import compute_class_weight
from sklearn.metrics import silhouette_score

from sklearn.model_selection import train_test_split, cross_val_score, learning_curve, RandomizedSearchCV,KFold
from sklearn.ensemble import RandomForestRegressor,GradientBoostingRegressor
# import optuna
import xgboost as xgb
from xgboost import XGBRegressor
from sklearn.metrics import mean_absolute_error, mean_squared_error, mean_absolute_percentage_error,r2_score, precision_score,
recall_score, f1_score,roc_auc_score, roc_curve

datafile = '/content/drive/MyDrive/ML Copy of Gender-Split TBI Dataset_08072026.xlsx'
data = pd.read_excel(datafile, sheet_name="tbi_datasheet_f_only")
```

```
# Display total number of rows
print(f"Total number of rows: {len(data)}")

# Display valid data count (non-null) for each column
print("\nValid data count per column:")
print(data.count())

# Display a complete list of column names
print("\nFull list of columns:")
print(data.columns.tolist())

print(data.info())
Standardize Severity into two categories: Mild and Moderate-Severe
Case-insensitive mapping for various labels
severity_mapping = { 'severe': 'Severe', 'moderate_severe': 'Moderate-severe' }
Convert to lowercase and strip whitespace for robust matching
data['injury_severity'] = data['injury_severity'].astype(str).str.lower().str.strip() data['injury_severity'] =
data['injury_severity'].replace(severity_mapping)
Verify the result
print("Standardized severity counts:") print(data['injury_severity'].value_counts())
Define the primary outcome based on the specified priority logic
data['outcome'] = data['log_effect']
Display summary of the new outcome column
print("Outcome column created. Non-null values:") print(data['outcome'].notna().sum())

Distribution by severity
feature_under_study = 'injury_severity'
levels = data[feature_under_study].unique()
xticklabels = [f"{sev}" for sev in levels]

# Group by severity and calculate median for the primary outcome (Baseline/FU %)
changes_total = data.groupby(feature_under_study)['outcome']
changes_total_mean = changes_total.median()
```

```
print("Median scores by severity:")
print(changes_total_mean)
Print out the standardized severity categories found in the dataset
print("Unique severity levels:", levels)
import numpy as np

# Re-evaluate levels to ensure it reflects the latest standardized severity categories
levels = data[feature_under_study].unique()

# Initialize dictionary to store mean standard error for each severity level
mean_std_err_severity_total = {}

for l in levels:
    severity_df = data[data[feature_under_study] == l].copy()

    # Ensure 'std_err' is numeric, coercing errors to NaN
    severity_df['std_err'] = pd.to_numeric(severity_df['std_err'], errors='coerce')

    # Calculate the mean of the existing 'std_err' column for the group
    if not severity_df['std_err'].dropna().empty:
        mean_std_err_severity_total[l] = severity_df['std_err'].mean()
    else:
        print(f"No valid data (std_err) for group: {l}. Setting mean_std_err to NaN.")
        mean_std_err_severity_total[l] = np.nan

# The ci_lower_severity_total and ci_upper_severity_total dictionaries are no longer needed
# as we are now using standard error for plotting instead of confidence intervals.

Calculate error bars using the mean standard error for each severity level
For standard error, yerr will be symmetric.
Ensure levels are consistent with mean_std_err_severity_total
error_values_total = pd.Series({sev: mean_std_err_severity_total.get(sev, np.nan) for sev in levels})
```

```
The errors for plotting will be the array of mean standard error values
errors_for_plot = error_values_total.values
print(error_values_total)

data_points_per_severity = data.groupby('injury_severity').size()
# Filter out NaN values from levels before using them as keys
valid_levels = [l for l in levels if pd.notna(l)]
custom_xticks = [f"{l}\n(n={int(data_points_per_severity[l])})" for l in valid_levels]

import pandas as pd
import numpy as np
import matplotlib.pyplot as plt
import seaborn as sns
import statsmodels.stats.meta_analysis as anova

# --- Refactored Plotting Code ---
# Using pre-calculated 'outcome' (Baseline/FU %) and 'SE outcome' from previous cells

plt.figure(figsize=(10, 6))

# Use valid levels identified earlier
valid_levels = [l for l in levels if pd.notna(l)]

# Create a numerical representation for plotting
severity_mapping_plot = {sev: i for i, sev in enumerate(valid_levels)}
data['Severity_numeric'] = data['injury_severity'].map(severity_mapping_plot)

# Add jitter to the x-axis for individual data points
jitter = 0.1
x_jittered = data['Severity_numeric'] + np.random.uniform(-jitter, jitter, size=len(data))

# Scatter plot of individual outcome points
plt.scatter(x_jittered, data['outcome'], alpha=0.5, label='Individual data points')
```

```
# Reindex medians to match valid_levels order
mean_changes_series = changes_total_mean.reindex(valid_levels)

# Use the pre-calculated errors_for_plot (mean standard error) from the previous cell
# Plot median outcome with error bars (Standard Error)
plt.errorbar(list(severity_mapping_plot.values()), mean_changes_series.values, yerr=errors_for_plot, fmt='o', color='red',
label='Median score % with Standard Error', capsize=5)

# Set x-axis ticks and labels
data_points_per_severity = data.groupby('injury_severity').size()
custom_xticks = [f"{l}\n(n={int(data_points_per_severity[l])})" for l in valid_levels]
plt.xticks(list(severity_mapping_plot.values()), custom_xticks)

# Add labels and title reflecting the primary outcome
plt.xlabel("Injury severity")
plt.ylabel("Log effect mortality")
plt.title("Log effect mortality by injury severity")
plt.legend()
plt.grid(True, linestyle='--', alpha=0.6)
plt.tight_layout()
plt.show()

1. Create a copy of the original data for modifications
modified_data = data.copy()
Ensure the 'outcome' column is present in modified_data
modified_data['outcome'] = modified_data['log_effect']
Standardize Severity into two categories: Moderate-Severe and Severe within modified_data
This ensures consistency for all subsequent analyses that rely on modified_data
severity_mapping = { 'severe': 'Severe', 'moderate_severe': 'Moderate-Severe' }
Convert to lowercase and strip whitespace for robust matching on the original data first
Then replace based on the mapping
```

modified_data['injury_severity'] = modified_data['injury_severity'].astype(str).str.lower().str.strip() modified_data['injury_severity'] = modified_data['injury_severity'].replace(severity_mapping)
print("Standardized severity counts in modified_data:") print(modified_data['injury_severity'].value_counts())

Check for outliers
Check for positive extreme outliers using the current outcome metric
Using a threshold relevant to test score % (e.g., studies with scores > 0.5)
positive_extreme_outliers = modified_data[modified_data['outcome'] > 0.5]
if positive_extreme_outliers.empty: print("No data points found with 'outcome' greater than 0.4 in the entire dataset.") else:
print("Data points with 'outcome' greater than 0.4:") display(positive_extreme_outliers[['outcome', 'Severity_numeric', 'author_year']])
Identify extreme outliers
outlier = modified_data[ (modified_data['Severity_numeric']) & (modified_data['outcome'] < 0.2) | (modified_data['outcome'] > 0.3) ]
print("Identified extreme outliers based on outcome:") display(outlier)
from matplotlib import pyplot as plt
### Use 'Test score %' as the column to align with the current outcome definition
outlier['outcome'].plot(kind='hist', bins=20, title='Distribution of log effect for outliers')
plt.xlabel('Log effect mortality')
plt.ylabel('Frequency')
plt.gca().spines[['top', 'right']].set_visible(False)
plt.show()

**2.Linear Regression**
Gender/sex
cleaned_gendersex_data = modified_data.dropna(subset=['sex_gender_ratio_men_males', 'outcome'])

mod_sev_gendersex_data = cleaned_gendersex_data[cleaned_gendersex_data['injury_severity'] == 'Moderate-Severe']
sev_gendersex_data = cleaned_gendersex_data[cleaned_gendersex_data['injury_severity'] == 'Severe']

print("Shape of Moderate-Severe data:", mod_sev_gendersex_data.shape)
print("Shape of Severe data:", sev_gendersex_data.shape)

```
import statsmodels.api as sm

# Linear Regression for Moderate-Severe Severity
X_mod_sev_gendersex = mod_sev_gendersex_data['sex_gender_ratio_men_males']
y_mod_sev_gendersex = mod_sev_gendersex_data['outcome']
X_mod_sev_gendersex = sm.add_constant(X_mod_sev_gendersex) # Add a constant for the intercept
model_mod_sev_gendersex = sm.OLS(y_mod_sev_gendersex, X_mod_sev_gendersex).fit()
print("\nLinear Regression Results for Moderate-Severe Severity (Gender/sex):")
print(model_mod_sev_gendersex.summary())

# Linear Regression for Severe Severity
X_sev_gendersex = sev_gendersex_data['sex_gender_ratio_men_males']
y_sev_gendersex = sev_gendersex_data['outcome']
X_sev_gendersex = sm.add_constant(X_sev_gendersex) # Add a constant for the intercept
model_sev_gendersex = sm.OLS(y_sev_gendersex, X_sev_gendersex).fit()
print("\nLinear Regression Results for Severe Severity (Gender/sex):")
print(model_sev_gendersex.summary())

plt.figure(figsize=(10, 7))

# Plot Moderate-Severe severity data points
sns.scatterplot(
    x=mod_sev_gendersex_data['sex_gender_ratio_men_males'],
    y=mod_sev_gendersex_data['outcome'],
    label='Moderate-Severe',
    color='green',
    alpha=0.6,
    marker='s', # square markers
    s=80 # size of points
)

# Plot Severe severity data points
sns.scatterplot(
```

```
    x=sev_gendersex_data['sex_gender_ratio_men_males'],
    y=sev_gendersex_data['outcome'],
    label='Severe',
    color='orange',
    alpha=0.6,
    marker='s', # square markers
    s=80 # size of points
)

# Overlay regression line for Moderate-Severe Severity
x_range_mod_sev = np.linspace(mod_sev_gendersex_data['sex_gender_ratio_men_males'].min(),
mod_sev_gendersex_data['sex_gender_ratio_men_males'].max(), 100)
y_pred_mod_sev = model_mod_sev_gendersex.predict(sm.add_constant(x_range_mod_sev))
plt.plot(x_range_mod_sev, y_pred_mod_sev, color='darkgreen', linestyle='-', label='Regression Line (Moderate-Severe)')

# Overlay regression line for Severe Severity
x_range_sev = np.linspace(sev_gendersex_data['sex_gender_ratio_men_males'].min(),
sev_gendersex_data['sex_gender_ratio_men_males'].max(), 100)
y_pred_sev = model_sev_gendersex.predict(sm.add_constant(x_range_sev))
plt.plot(x_range_sev, y_pred_sev, color='darkorange', linestyle='-', label='Regression Line (Severe)')

plt.xlabel('Gender/sex, men/males proportion')
plt.ylabel('Log effect mortality')
plt.title('Log effect mortality vs. men/males proportion by injury severity')

r_squared_mod_sev = model_mod_sev_gendersex.rsquared
p_value_mod_sev = model_mod_sev_gendersex.pvalues['sex_gender_ratio_men_males']
plt.text(0.05, 0.90, f'Moderate-severe: $R^2$={r_squared_mod_sev:.3f}, p={p_value_mod_sev:.3f}', transform=plt.gca().transAxes,
fontsize=10, verticalalignment='top', bbox=dict(boxstyle='round,pad=0.3', fc='white', alpha=0.5))

r_squared_sev = model_sev_gendersex.rsquared
p_value_sev = model_sev_gendersex.pvalues['sex_gender_ratio_men_males']
```

```
plt.text(0.05, 0.85, f'Severe: $R^2$={r_squared_sev:.3f}, p={p_value_sev:.3f}', transform=plt.gca().transAxes, fontsize=10, verticalalignment='top', bbox=dict(boxstyle='round,pad=0.3', fc='white', alpha=0.5))

plt.legend()
plt.grid(True, linestyle='--', alpha=0.7)
plt.tight_layout()
plt.show()
```

In-hospital - for sensitivity

```
in_hospital_data= modified_data[modified_data['mortality_type: in_hospital'] == 1]

in_hospital_outcome_data = in_hospital_data.dropna(subset=['sex_gender_ratio_men_males', 'outcome'])

print("Shape of the filtered DataFrame (in_hospital_outcome_data):")
print(in_hospital_outcome_data.shape)

print("First 5 rows of the filtered DataFrame:")
print(in_hospital_outcome_data.head())

mod_sev_ih_data = in_hospital_outcome_data[in_hospital_outcome_data['injury_severity'] == 'Moderate-Severe']
sev_ih_data = in_hospital_outcome_data[in_hospital_outcome_data['injury_severity'] == 'Severe']

print("Shape of Moderate-Severe data:", mod_sev_ih_data.shape)
print("Shape of Severe data:", sev_ih_data.shape)
```

Linear Regression for Moderate-Severe Severity

```
X_mod_sev_ih = mod_sev_ih_data['sex_gender_ratio_men_males'] y_mod_sev_ih = mod_sev_ih_data['outcome'] X_mod_sev_ih = sm.add_constant(X_mod_sev_ih) # Add a constant for the intercept model_mod_sev_ih = sm.OLS(y_mod_sev_ih, X_mod_sev_ih).fit() print("\nLinear Regression Results for Moderate-Severe Severity (Gender/sex):")
print(model_mod_sev_ih.summary())
```

Linear Regression for Severe Severity

```
X_sev_ih = sev_ih_data['sex_gender_ratio_men_males'] y_sev_ih = sev_ih_data['outcome'] X_sev_ih = sm.add_constant(X_sev_ih) # Add a constant for the intercept model_sev_ih = sm.OLS(y_sev_ih, X_sev_ih).fit() print("\nLinear Regression Results for Severe Severity (Gender/sex):") print(model_sev_ih.summary())
plt.figure(figsize=(10, 7))

# Plot Moderate-Severe severity data points
sns.scatterplot(
    x=mod_sev_ih_data['sex_gender_ratio_men_males'],
    y=mod_sev_ih_data['outcome'],
    label='Moderate-Severe',
    color='green',
    alpha=0.6,
    marker='s', # square markers
    s=80 # size of points
)

# Plot Severe severity data points
sns.scatterplot(
    x=sev_ih_data['sex_gender_ratio_men_males'],
    y=sev_ih_data['outcome'],
    label='Severe',
    color='orange',
    alpha=0.6,
    marker='s', # square markers
    s=80 # size of points
)

# Overlay regression line for Moderate-Severe Severity
x_range_mod_sev = np.linspace(mod_sev_ih_data['sex_gender_ratio_men_males'].min(),
mod_sev_ih_data['sex_gender_ratio_men_males'].max(), 100)
y_pred_mod_sev = model_mod_sev_ih.predict(sm.add_constant(x_range_mod_sev))
plt.plot(x_range_mod_sev, y_pred_mod_sev, color='darkgreen', linestyle='-', label='Regression Line (Moderate-Severe)')
```

```
# Overlay regression line for Severe Severity
x_range_sev = np.linspace(sev_ih_data['sex_gender_ratio_men_males'].min(), sev_ih_data['sex_gender_ratio_men_males'].max(), 100)
y_pred_sev = model_sev_ih.predict(sm.add_constant(x_range_sev))
plt.plot(x_range_sev, y_pred_sev, color='darkorange', linestyle='-', label='Regression Line (Severe)')

plt.xlabel('Gender/sex (proportion of men/males)')
plt.ylabel('Log effect mortality')
plt.title('Log effect mortality vs. gender/sex for in-hospital mortality')

r_squared_mod_sev = model_mod_sev_ih.rsquared
p_value_mod_sev = model_mod_sev_ih.pvalues['sex_gender_ratio_men_males']
plt.text(0.05, 0.90, f'Moderate-severe: $R^2$={r_squared_mod_sev:.3f}, p={p_value_mod_sev:.3f}', transform=plt.gca().transAxes, fontsize=10, verticalalignment='top', bbox=dict(boxstyle='round,pad=0.3', fc='white', alpha=0.5))

r_squared_sev = model_sev_ih.rsquared
p_value_sev = model_sev_ih.pvalues['sex_gender_ratio_men_males']
plt.text(0.05, 0.85, f'Severe: $R^2$={r_squared_sev:.3f}, p={p_value_sev:.3f}', transform=plt.gca().transAxes, fontsize=10, verticalalignment='top', bbox=dict(boxstyle='round,pad=0.3', fc='white', alpha=0.5))

plt.legend()
plt.grid(True, linestyle='--', alpha=0.7)
plt.tight_layout()
plt.show()
```

Unspecified mortality

```
unspecified_data= modified_data[modified_data['mortality_type: unspecified_overall'] == 1]
unspecified_outcome_data = unspecified_data.dropna(subset=['sex_gender_ratio_men_males', 'outcome'])

mod_sev_unspecified_data = unspecified_outcome_data[unspecified_outcome_data['injury_severity'] == 'Moderate-Severe']
sev_unspecified_data = unspecified_outcome_data[unspecified_outcome_data['injury_severity'] == 'Severe']

print("Shape of Moderate-Severe data:", mod_sev_unspecified_data.shape)
```

print("Shape of Severe data:", sev_unspecified_data.shape)

Linear Regression for Moderate-Severe Severity
X_mod_sev_unspecified = mod_sev_unspecified_data['sex_gender_ratio_men_males'] y_mod_sev_unspecified = mod_sev_unspecified_data['outcome'] X_mod_sev_unspecified = sm.add_constant(X_mod_sev_unspecified) # Add a constant for the intercept model_mod_sev_unspecified = sm.OLS(y_mod_sev_unspecified, X_mod_sev_unspecified).fit() print("\nLinear Regression Results for Moderate-Severe Severity (Gender/sex):") print(model_mod_sev_unspecified.summary())
Linear Regression for Severe Severity cannot be done b/c only 1 data point

```
plt.figure(figsize=(10, 7))

# Plot Moderate-Severe severity data points
sns.scatterplot(
    x=mod_sev_unspecified_data['sex_gender_ratio_men_males'],
    y=mod_sev_unspecified_data['outcome'],
    label='Moderate-Severe',
    color='green',
    alpha=0.6,
    marker='s', # square markers
    s=80 # size of points
)

# Plot Severe severity data points
sns.scatterplot(
    x=sev_unspecified_data['sex_gender_ratio_men_males'],
    y=sev_unspecified_data['outcome'],
    label='Severe',
    color='orange',
    alpha=0.6,
    marker='s', # square markers
    s=80 # size of points
)
```

```
# Overlay regression line for Moderate-Severe Severity
x_range_mod_sev = np.linspace(mod_sev_unspecified_data['sex_gender_ratio_men_males'].min(),
mod_sev_unspecified_data['sex_gender_ratio_men_males'].max(), 100)
y_pred_mod_sev = model_mod_sev_unspecified.predict(sm.add_constant(x_range_mod_sev))
plt.plot(x_range_mod_sev, y_pred_mod_sev, color='darkgreen', linestyle='-', label='Regression Line (Moderate-Severe)')

plt.xlabel('Gender/sex (proportion men/males)')
plt.ylabel('Log effect mortality')
plt.title('Log effect mortality vs. gender/sex for unspecified mortality')

r_squared_mod_sev = model_mod_sev_unspecified.rsquared
p_value_mod_sev = model_mod_sev_unspecified.pvalues['sex_gender_ratio_men_males']
plt.text(0.05, 0.90, f'Moderate-severe: $R^2$={r_squared_mod_sev:.3f}, p={p_value_mod_sev:.3f}', transform=plt.gca().transAxes,
fontsize=10, verticalalignment='top', bbox=dict(boxstyle='round,pad=0.3', fc='white', alpha=0.5))

plt.legend()
plt.grid(True, linestyle='--', alpha=0.7)
plt.tight_layout()
plt.show()
```

1-month mortality

```
onem_data= modified_data[modified_data['mortality_type: 1_month'] == 1]
onem_outcome_data = onem_data.dropna(subset=['sex_gender_ratio_men_males', 'outcome'])

mod_sev_onem_data = onem_outcome_data[onem_outcome_data['injury_severity'] == 'Moderate-severe']
sev_onem_data = onem_outcome_data[onem_outcome_data['injury_severity'] == 'Severe']

print("Shape of Moderate-Severe data:", mod_sev_onem_data.shape)
print("Shape of Severe data:", sev_onem_data.shape)
```

```
import statsmodels.api as sm

# Linear Regression for Moderate-Severe Severity
if len(mod_sev_onem_data) >= 2:
    X_mod_sev_onem = mod_sev_onem_data['sex_gender_ratio_men_males']
    y_mod_sev_onem = mod_sev_onem_data['outcome']
    X_mod_sev_onem = sm.add_constant(X_mod_sev_onem) # Add a constant for the intercept
    model_mod_sev_onem = sm.OLS(y_mod_sev_onem, X_mod_sev_onem).fit()
    print("\nLinear Regression Results for Moderate-Severe Severity (Gender/sex, 1-month):")
    print(model_mod_sev_onem.summary())
else:
    print("\nSkipping Linear Regression for Moderate-Severe Severity (Gender/sex, 1-month) due to insufficient data (less than 2 observations).")

# Linear Regression for Severe Severity
if len(sev_onem_data) >= 2:
    X_sev_onem = sev_onem_data['sex_gender_ratio_men_males']
    y_sev_onem = sev_onem_data['outcome']
    X_sev_onem = sm.add_constant(X_sev_onem) # Add a constant for the intercept
    model_sev_onem = sm.OLS(y_sev_onem, X_sev_onem).fit()
    print("\nLinear Regression Results for Severe Severity (Gender/sex, 1-month):")
    print(model_sev_onem.summary())
else:
    print("\nSkipping Linear Regression for Severe Severity (Gender/sex, 1-month) due to insufficient data (less than 2 observations).")

plt.figure(figsize=(10, 7))

# Plot Moderate-Severe severity data points if available
if len(mod_sev_onem_data) > 0:
    sns.scatterplot(
        x=mod_sev_onem_data['sex_gender_ratio_men_males'],
```

```
        y=mod_sev_onem_data['outcome'],
        label='Moderate-Severe',
        color='green',
        alpha=0.6,
        marker='s', # square markers
        s=80 # size of points
    )

# Plot Severe severity data points if available
if len(sev_onem_data) > 0:
    sns.scatterplot(
        x=sev_onem_data['sex_gender_ratio_men_males'],
        y=sev_onem_data['outcome'],
        label='Severe',
        color='orange',
        alpha=0.6,
        marker='s', # square markers
        s=80 # size of points
    )

# Overlay regression line for Moderate-Severe Severity if model exists
if 'model_mod_sev_onem' in locals() and len(mod_sev_onem_data) >= 2:
    x_range_mod_sev = np.linspace(mod_sev_onem_data['sex_gender_ratio_men_males'].min(),
mod_sev_onem_data['sex_gender_ratio_men_males'].max(), 100)
    y_pred_mod_sev = model_mod_sev_onem.predict(sm.add_constant(x_range_mod_sev))
    plt.plot(x_range_mod_sev, y_pred_mod_sev, color='darkgreen', linestyle='-', label='Regression Line (Moderate-Severe)')

# Overlay regression line for Severe Severity if model exists
if 'model_sev_onem' in locals() and len(sev_onem_data) >= 2:
    x_range_sev = np.linspace(sev_onem_data['sex_gender_ratio_men_males'].min(),
sev_onem_data['sex_gender_ratio_men_males'].max(), 100)
    y_pred_sev = model_sev_onem.predict(sm.add_constant(x_range_sev))
    plt.plot(x_range_sev, y_pred_sev, color='darkorange', linestyle='-', label='Regression Line (Severe)')
```

```
plt.xlabel('Gender/sex (proportion men/males)')
plt.ylabel('Log effect mortality')
plt.title('Log effect mortality vs. gender/sex for 1-month mortality')

# Add R-squared and p-value for Moderate-Severe if model exists
if 'model_mod_sev_onem' in locals() and len(mod_sev_onem_data) >= 2:
    r_squared_mod_sev = model_mod_sev_onem.rsquared
    p_value_mod_sev = model_mod_sev_onem.pvalues['sex_gender_ratio_men_males']
    plt.text(0.05, 0.90, f'Moderate-severe: $R^2$={r_squared_mod_sev:.3f}, p={p_value_mod_sev:.3f}',
transform=plt.gca().transAxes, fontsize=10, verticalalignment='top', bbox=dict(boxstyle='round,pad=0.3', fc='white', alpha=0.5))

# Add R-squared and p-value for Severe if model exists
if 'model_sev_onem' in locals() and len(sev_onem_data) >= 2:
    r_squared_sev = model_sev_onem.rsquared
    p_value_sev = model_sev_onem.pvalues['sex_gender_ratio_men_males']
    # Adjust position if Moderate-Severe data is also plotted
    if 'model_mod_sev_onem' in locals() and len(mod_sev_onem_data) >= 2:
        plt.text(0.05, 0.85, f'Severe: $R^2$={r_squared_sev:.3f}, p={p_value_sev:.3f}', transform=plt.gca().transAxes, fontsize=10,
verticalalignment='top', bbox=dict(boxstyle='round,pad=0.3', fc='white', alpha=0.5))
    else:
        plt.text(0.05, 0.90, f'Severe: $R^2$={r_squared_sev:.3f}, p={p_value_sev:.3f}', transform=plt.gca().transAxes, fontsize=10,
verticalalignment='top', bbox=dict(boxstyle='round,pad=0.3', fc='white', alpha=0.5))

plt.legend()
plt.grid(True, linestyle='--', alpha=0.7)
plt.tight_layout()
plt.show()
```

6-month mortality

```
sixm_data= modified_data[modified_data['mortality_type: 6_month'] == 1]
sixm_outcome_data = sixm_data.dropna(subset=['sex_gender_ratio_men_males', 'outcome'])
```

```
mod_sev_sixm_data = sixm_outcome_data[sixm_outcome_data['injury_severity'] == 'moderate-severe']
sev_sixm_data = sixm_outcome_data[sixm_outcome_data['injury_severity'] == 'Severe']

print("Shape of Moderate-Severe data:", mod_sev_sixm_data.shape)
print("Shape of Severe data:", sev_sixm_data.shape)
```

12-month mortality

```
twelvem_data= modified_data[modified_data['mortality_type: 12_month'] == 1]
twelvem_outcome_data = twelvem_data.dropna(subset=['sex_gender_ratio_men_males', 'outcome'])

mod_sev_twelvem_data = twelvem_outcome_data[twelvem_outcome_data['injury_severity'] == 'moderate-severe']
sev_twelvem_data = twelvem_outcome_data[twelvem_outcome_data['injury_severity'] == 'Severe']

print("Shape of Moderate-Severe data:", mod_sev_twelvem_data.shape)
print("Shape of Severe data:", sev_twelvem_data.shape)
```

Age

```
cleaned_age_data = modified_data.dropna(subset=['age_mean', 'outcome'])

mod_sev_age_data = cleaned_age_data[cleaned_age_data['injury_severity'] == 'moderate_severe']
sev_age_data = cleaned_age_data[cleaned_age_data['injury_severity'] == 'severe']

print("Shape of Moderate-Severe data:", mod_sev_age_data.shape)
print("Shape of Severe data:", sev_age_data.shape)

# Adding a debug print to inspect injury_severity values in cleaned_age_data
print("\nValue counts for 'injury_severity' in cleaned_age_data before filtering:")
print(cleaned_age_data['injury_severity'].value_counts())

import statsmodels.api as sm
```

```
# Linear Regression for Moderate-Severe Severity
X_mod_sev_age = mod_sev_age_data['age_mean']
y_mod_sev_age = mod_sev_age_data['outcome']
X_mod_sev_age = sm.add_constant(X_mod_sev_age) # Add a constant for the intercept
model_mod_sev_age = sm.OLS(y_mod_sev_age, X_mod_sev_age).fit()
print("\nLinear Regression Results for Moderate-Severe Severity (Age):")
print(model_mod_sev_age.summary())

# Linear Regression for Severe Severity
X_sev_age = sev_age_data['age_mean']
y_sev_age = sev_age_data['outcome']
X_sev_age = sm.add_constant(X_sev_age) # Add a constant for the intercept
model_sev_age = sm.OLS(y_sev_age, X_sev_age).fit()
print("\nLinear Regression Results for Severe Severity (Age):")
print(model_sev_age.summary())

plt.figure(figsize=(10, 7))

# Plot Moderate-Severe severity data points
sns.scatterplot(
    x=mod_sev_age_data['age_mean'],
    y=mod_sev_age_data['outcome'],
    label='Moderate-Severe',
    color='green',
    alpha=0.6,
    marker='s', # square markers
    s=80 # size of points
)

# Plot Severe severity data points
sns.scatterplot(
    x=sev_age_data['age_mean'],
```

```
    y=sev_age_data['outcome'],
    label='Severe',
    color='orange',
    alpha=0.6,
    marker='s', # square markers
    s=80 # size of points
)

# Overlay regression line for Moderate-Severe Severity
x_range_mod_sev = np.linspace(mod_sev_age_data['age_mean'].min(), mod_sev_age_data['age_mean'].max(), 100)
y_pred_mod_sev = model_mod_sev_age.predict(sm.add_constant(x_range_mod_sev))
plt.plot(x_range_mod_sev, y_pred_mod_sev, color='darkgreen', linestyle='-', label='Regression Line (Moderate-Severe)')

# Overlay regression line for Severe Severity
x_range_sev = np.linspace(sev_age_data['age_mean'].min(), sev_age_data['age_mean'].max(), 100)
y_pred_sev = model_sev_age.predict(sm.add_constant(x_range_sev))
plt.plot(x_range_sev, y_pred_sev, color='darkorange', linestyle='-', label='Regression Line (Severe)')

plt.xlabel('Age, mean years')
plt.ylabel('Log effect mortality')
plt.title('Log effect mortality vs. mean age')

r_squared_mod_sev = model_mod_sev_age.rsquared
p_value_mod_sev = model_mod_sev_age.pvalues['age_mean']
plt.text(0.05, 0.90, f'Moderate-severe: $R^2$={r_squared_mod_sev:.3f}, p={p_value_mod_sev:.3f}', transform=plt.gca().transAxes,
fontsize=10, verticalalignment='top', bbox=dict(boxstyle='round,pad=0.3', fc='white', alpha=0.5))

r_squared_sev = model_sev_age.rsquared
p_value_sev = model_sev_age.pvalues['age_mean']
plt.text(0.05, 0.85, f'Severe: $R^2$={r_squared_sev:.3f}, p={p_value_sev:.3f}', transform=plt.gca().transAxes, fontsize=10,
verticalalignment='top', bbox=dict(boxstyle='round,pad=0.3', fc='white', alpha=0.5))

plt.legend()
```

```
plt.grid(True, linestyle='--', alpha=0.7)
plt.tight_layout()
plt.show()
```

Age SD

```
cleaned_age_data = modified_data.dropna(subset=['age_mean', 'outcome'])
```

Study quality

```
cleaned_qual_data = modified_data.dropna(subset=['study_quality', 'outcome'])

mod_sev_qual_data = cleaned_qual_data[cleaned_qual_data['injury_severity'] == 'Moderate-Severe']
sev_qual_data = cleaned_qual_data[cleaned_qual_data['injury_severity'] == 'Severe']

print("Shape of Moderate-Severe data:", mod_sev_qual_data.shape)
print("Shape of Severe data:", sev_qual_data.shape)
```

Linear Regression for Moderate-Severe Severity

```
X_mod_sev_qual = mod_sev_qual_data['study_quality'] y_mod_sev_qual = mod_sev_qual_data['outcome'] X_mod_sev_qual =
sm.add_constant(X_mod_sev_qual) # Add a constant for the intercept model_mod_sev_qual = sm.OLS(y_mod_sev_qual,
X_mod_sev_qual).fit() print("\nLinear Regression Results for Moderate-Severe Severity (Study quality):")
print(model_mod_sev_qual.summary())
```

Linear Regression for Severe Severity

```
X_sev_qual = sev_qual_data['study_quality'] y_sev_qual = sev_qual_data['outcome'] X_sev_qual = sm.add_constant(X_sev_qual) #
Add a constant for the intercept model_sev_qual = sm.OLS(y_sev_qual, X_sev_qual).fit() print("\nLinear Regression Results for
Severe Severity (Study quality)):") print(model_sev_qual.summary())
plt.figure(figsize=(10, 7))

# Plot Moderate-Severe severity data points
sns.scatterplot(
    x=mod_sev_qual_data['study_quality'],
    y=mod_sev_qual_data['outcome'],
```

```
    label='Moderate-Severe',
    color='green',
    alpha=0.6,
    marker='s', # square markers
    s=80 # size of points
)

# Plot Severe severity data points
sns.scatterplot(
    x=sev_qual_data['study_quality'],
    y=sev_qual_data['outcome'],
    label='Severe',
    color='orange',
    alpha=0.6,
    marker='s', # square markers
    s=80 # size of points
)

# Overlay regression line for Moderate-Severe Severity
x_range_mod_sev = np.linspace(mod_sev_qual_data['study_quality'].min(), mod_sev_qual_data['study_quality'].max(), 100)
y_pred_mod_sev = model_mod_sev_qual.predict(sm.add_constant(x_range_mod_sev))
plt.plot(x_range_mod_sev, y_pred_mod_sev, color='darkgreen', linestyle='-', label='Regression Line (Moderate-Severe)')

# Overlay regression line for Severe Severity
x_range_sev = np.linspace(sev_qual_data['study_quality'].min(), sev_qual_data['study_quality'].max(), 100)
y_pred_sev = model_sev_qual.predict(sm.add_constant(x_range_sev))
plt.plot(x_range_sev, y_pred_sev, color='darkorange', linestyle='-', label='Regression Line (Severe)')

plt.xlabel('Study quality (proportion)')
plt.ylabel('Log effect mortality')
plt.title('Log effect mortality vs. study quality')

r_squared_mod_sev = model_mod_sev_qual.rsquared
```

```
p_value_mod_sev = model_mod_sev_qual.pvalues['study_quality']
plt.text(0.05, 0.90, f'Moderate-severe: $R^2$={r_squared_mod_sev:.3f}, p={p_value_mod_sev:.3f}', transform=plt.gca().transAxes,
fontsize=10, verticalalignment='top', bbox=dict(boxstyle='round,pad=0.3', fc='white', alpha=0.5))

r_squared_sev = model_sev_qual.rsquared
p_value_sev = model_sev_qual.pvalues['study_quality']
plt.text(0.05, 0.85, f'Severe: $R^2$={r_squared_sev:.3f}, p={p_value_sev:.3f}', transform=plt.gca().transAxes, fontsize=10,
verticalalignment='top', bbox=dict(boxstyle='round,pad=0.3', fc='white', alpha=0.5))

plt.legend()
plt.grid(True, linestyle='--', alpha=0.7)
plt.tight_layout()
plt.show()
```

GII

```
cleaned_gii_data = modified_data.dropna(subset=['gender_ineq', 'outcome'])

mod_sev_gii_data = cleaned_gii_data[cleaned_gii_data['injury_severity'] == 'Moderate-Severe']
sev_gii_data = cleaned_gii_data[cleaned_gii_data['injury_severity'] == 'Severe']

print("Shape of Moderate-Severe data:", mod_sev_gii_data.shape)
print("Shape of Severe data:", sev_gii_data.shape)
```

Linear Regression for Moderate-Severe Severity

```
X_mod_sev_gii = mod_sev_gii_data['gender_ineq'] y_mod_sev_gii = mod_sev_gii_data['outcome'] X_mod_sev_gii =
sm.add_constant(X_mod_sev_gii) # Add a constant for the intercept model_mod_sev_gii = sm.OLS(y_mod_sev_gii,
X_mod_sev_gii).fit() print("\nLinear Regression Results for Moderate-Severe Severity (Gender inequality index):")
print(model_mod_sev_gii.summary())
```

Linear Regression for Severe Severity

```
X_sev_gii = sev_gii_data['gender_ineq'] y_sev_gii = sev_gii_data['outcome'] X_sev_gii = sm.add_constant(X_sev_gii) # Add a
constant for the intercept model_sev_gii = sm.OLS(y_sev_gii, X_sev_gii).fit() print("\nLinear Regression Results for Severe Severity
(Gender inequality index):") print(model_sev_gii.summary())
```

```
plt.figure(figsize=(10, 7))

# Plot Moderate-Severe severity data points
sns.scatterplot(
    x=mod_sev_gii_data['gender_ineq'],
    y=mod_sev_gii_data['outcome'],
    label='Moderate-Severe',
    color='green',
    alpha=0.6,
    marker='s', # square markers
    s=80 # size of points
)

# Plot Severe severity data points
sns.scatterplot(
    x=sev_gii_data['gender_ineq'],
    y=sev_gii_data['outcome'],
    label='Severe',
    color='orange',
    alpha=0.6,
    marker='s', # square markers
    s=80 # size of points
)

# Overlay regression line for Moderate-Severe Severity
x_range_mod_sev = np.linspace(mod_sev_gii_data['gender_ineq'].min(), mod_sev_gii_data['gender_ineq'].max(), 100)
y_pred_mod_sev = model_mod_sev_gii.predict(sm.add_constant(x_range_mod_sev))
plt.plot(x_range_mod_sev, y_pred_mod_sev, color='darkgreen', linestyle='-', label='Regression Line (Moderate-Severe)')

# Overlay regression line for Severe Severity
x_range_sev = np.linspace(sev_gii_data['gender_ineq'].min(), sev_gii_data['gender_ineq'].max(), 100)
y_pred_sev = model_sev_gii.predict(sm.add_constant(x_range_sev))
plt.plot(x_range_sev, y_pred_sev, color='darkorange', linestyle='-', label='Regression Line (Severe)')
```

```
plt.xlabel('Gender Inequality Index')
plt.ylabel('Log effect mortality')
plt.title('Log effect mortality vs. Gender Inequality Index')

r_squared_mod_sev = model_mod_sev_gii.rsquared
p_value_mod_sev = model_mod_sev_gii.pvalues['gender_ineq']
plt.text(0.05, 0.90, f'Moderate-severe: $R^2$={r_squared_mod_sev:.3f}, p={p_value_mod_sev:.3f}', transform=plt.gca().transAxes,
fontsize=10, verticalalignment='top', bbox=dict(boxstyle='round,pad=0.3', fc='white', alpha=0.5))

r_squared_sev = model_sev_gii.rsquared
p_value_sev = model_sev_gii.pvalues['gender_ineq']
plt.text(0.05, 0.85, f'Severe: $R^2$={r_squared_sev:.3f}, p={p_value_sev:.3f}', transform=plt.gca().transAxes, fontsize=10,
verticalalignment='top', bbox=dict(boxstyle='round,pad=0.3', fc='white', alpha=0.5))

plt.legend()
plt.grid(True, linestyle='--', alpha=0.7)
plt.tight_layout()
plt.show()
```

Sample size (n) - sensitivity

```
cleaned_n_data= modified_data['cohort_size']

mod_sev_n_data = cleaned_n_data[cleaned_n_data['injury_severity'] == 'Moderate-Severe']
sev_n_data = cleaned_n_data[cleaned_n_data['injury_severity'] == 'Severe']

print("Shape of Moderate-Severe data:", mod_sev_n_data.shape)
print("Shape of Severe data:", sev_n_data.shape)

non_numeric_cohort_size_rows = modified_data[pd.to_numeric(modified_data['cohort_size'], errors='coerce').isna()]

if not non_numeric_cohort_size_rows.empty:
```

```
    print("Rows with non-numeric values in 'cohort_size' (e.g., '#REF!'):")
    display(non_numeric_cohort_size_rows[['author_year', 'injury_severity', 'cohort_size', 'outcome']])
else:
    print("No non-numeric values found in the 'cohort_size' column of the original data.")

import statsmodels.api as sm

# Linear Regression for Moderate-Severe Severity
if not mod_sev_n_data.empty and pd.to_numeric(mod_sev_n_data['cohort_size'], errors='coerce').notna().all():
    X_mod_sev_n = mod_sev_n_data['cohort_size']
    y_mod_sev_n = mod_sev_n_data['outcome']
    X_mod_sev_n = sm.add_constant(X_mod_sev_n, prepend=False) # Add a constant for the intercept
    model_mod_sev_n = sm.OLS(y_mod_sev_n, X_mod_sev_n).fit()
    print("\nLinear Regression Results for Moderate-Severe Severity (Cohort size):")
    print(model_mod_sev_n.summary())
else:
    print("\nSkipping Linear Regression for Moderate-Severe Severity (Cohort size) due to empty or non-numeric data.")

# Linear Regression for Severe Severity
if not sev_n_data.empty and pd.to_numeric(sev_n_data['cohort_size'], errors='coerce').notna().all():
    X_sev_n = sev_n_data['cohort_size']
    y_sev_n = sev_n_data['outcome']
    X_sev_n = sm.add_constant(X_sev_n, prepend=False) # Add a constant for the intercept
    model_sev_n = sm.OLS(y_sev_n, X_sev_n).fit()
    print("\nLinear Regression Results for Severe Severity (Cohort size):")
    print(model_sev_n.summary())
else:
    print("\nSkipping Linear Regression for Severe Severity (Cohort size) due to empty or non-numeric data.")

plt.figure(figsize=(10, 7))

# Plot Moderate-Severe severity data points
```

```
sns.scatterplot(
    x=mod_sev_n_data['cohort_size'],
    y=mod_sev_n_data['outcome'],
    label='Moderate-Severe',
    color='green',
    alpha=0.6,
    marker='s', # square markers
    s=80 # size of points
)

# Plot Severe severity data points
sns.scatterplot(
    x=sev_n_data['cohort_size'],
    y=sev_n_data['outcome'],
    label='Severe',
    color='orange',
    alpha=0.6,
    marker='s', # square markers
    s=80 # size of points
)

# Overlay regression line for Moderate-Severe Severity
x_range_mod_sev = np.linspace(mod_sev_n_data['cohort_size'].min(), mod_sev_n_data['cohort_size'].max(), 100)
y_pred_mod_sev = model_mod_sev_n.predict(sm.add_constant(x_range_mod_sev))
plt.plot(x_range_mod_sev, y_pred_mod_sev, color='darkgreen', linestyle='-', label='Regression Line (Moderate-Severe)')

# Overlay regression line for Severe Severity
x_range_sev = np.linspace(sev_n_data['cohort_size'].min(), sev_n_data['cohort_size'].max(), 100)
y_pred_sev = model_sev_n.predict(sm.add_constant(x_range_sev))
plt.plot(x_range_sev, y_pred_sev, color='darkorange', linestyle='-', label='Regression Line (Severe)')

plt.xlabel('Cohort size')
plt.ylabel('Log effect mortality')
```

```
plt.title('Log effect mortality vs. cohort size')

r_squared_mod_sev = model_mod_sev_n.rsquared
p_value_mod_sev = model_mod_sev_n.pvalues['cohort_size']
plt.text(0.05, 0.90, f'Moderate-severe: $R^2$={r_squared_mod_sev:.3f}, p={p_value_mod_sev:.3f}', transform=plt.gca().transAxes,
fontsize=10, verticalalignment='top', bbox=dict(boxstyle='round,pad=0.3', fc='white', alpha=0.5))

r_squared_sev = model_sev_n.rsquared
p_value_sev = model_sev_n.pvalues['cohort_size']
plt.text(0.05, 0.85, f'Severe: $R^2$={r_squared_sev:.3f}, p={p_value_sev:.3f}', transform=plt.gca().transAxes, fontsize=10,
verticalalignment='top', bbox=dict(boxstyle='round,pad=0.3', fc='white', alpha=0.5))

plt.legend()
plt.grid(True, linestyle='--', alpha=0.7)
plt.tight_layout()
plt.show()

plt.figure(figsize=(10, 6))
sns.countplot(data=modified_data, x='injury_severity', hue='above_median', palette='viridis')
plt.title('Distribution of above/below median cohort size by injury severity')
plt.xlabel('Injury Severity')
plt.ylabel('Count')
plt.legend(title='Above Median (1=True)')
plt.grid(axis='y', linestyle='--', alpha=0.7)
plt.tight_layout()
plt.show()
```

**3.Heatmaps by severity**

this is where you describe your predictors, you can add and remove as neede

```
predictors_overall = ['age_mean', 'sex_gender_ratio_men_males', 'study_quality', 'gender_ineq'] outcome = 'outcome'
df = [predictors_overall + [outcome]]
def compute_corr_subset(df_input, outcome_col):
```

```
    # Ensure the outcome_col is in the dataframe
    if outcome_col not in df_input.columns:
        return pd.Series(dtype='float64')

    # Calculate correlations of all other columns with the outcome column
    predictor_cols = [col for col in df_input.columns if col != outcome_col]
    if not predictor_cols:
        return pd.Series(dtype='float64') # Return empty series if no predictors

    # corrwith computes pairwise correlation between rows or columns of DataFrame with a Series or DataFrame.
    correlations = df_input[predictor_cols].corrwith(df_input[outcome_col])
    return correlations
def find_high_corr_features(corr_matrix, threshold=0.6, severity=None):
  # Extract upper triangle of the correlation matrix without the diagonal
  mask = np.triu(np.ones(corr_matrix.shape), k=1).astype(bool)
  upper = corr_matrix.where(mask)

  # Find pairs with correlation greater than the threshold
  high_corr_pairs = (
      upper.stack()
      .reset_index()
      .rename(columns={'level_0': 'Feature_1', 'level_1': 'Feature_2', 0: 'Correlation'})
  )
  # Filter by threshold
  high_corr_pairs = high_corr_pairs[(high_corr_pairs['Correlation'] > threshold) | (high_corr_pairs['Correlation'] < -threshold)]
  if severity:
      print(f"High Correlation Pairs {severity}(threshold={threshold}):")
  else:
      print(f"High Correlation Pairs (threshold={threshold}):")
  print(high_corr_pairs)
  return high_corr_pairs

def find_high_corr_features(corr_matrix, severity, threshold=0.6):
```

```
  # Extract upper triangle of the correlation matrix without the diagonal
  mask = np.triu(np.ones(corr_matrix.shape), k=1).astype(bool)
  upper = corr_matrix.where(mask)

  # Find pairs with correlation greater than the threshold
  high_corr_pairs = (
      upper.stack()
      .reset_index()
      .rename(columns={'level_0': 'Feature_1', 'level_1': 'Feature_2', 0: 'Correlation'})
  )
  # Filter by threshold
  high_corr_pairs = high_corr_pairs[(high_corr_pairs['Correlation'] > threshold) | (high_corr_pairs['Correlation'] < -threshold)]
  print(f"High Correlation Pairs {severity}(threshold={threshold}):")
  print(high_corr_pairs)
  return high_corr_pairs

#modsev/sev data split
df_modsev = modified_data[modified_data['injury_severity'] == 'moderate-severe']
df_sev = modified_data[modified_data['injury_severity'] == 'Severe']

for df_severity, name_severity in [(df_modsev, "moderate-severe"), (df_sev, "Severe")]:
  data = df_severity.copy()
  data = data.dropna(subset=predictors_overall + [outcome])

  if data.empty or len(data) < 2:
      print(f"Skipping analysis due to insufficient {name_severity} data after dropping NaNs.")
  else:
      X = data[predictors_overall + [outcome]].copy() # Include outcome for correlation calculation
      X = pd.get_dummies(X)
      X = X.dropna()  # Make sure there are no NaNs
      X = X.apply(pd.to_numeric)
      X = X.astype(float)
```

```
if X.empty or len(X) < 1:
    print(f"Skipping analysis after get_dummies and dropna due to insufficient data for {name_severity}.")
else:
    current_predictors = [col for col in X.columns if col != outcome] # These are the actual columns in X (including dummies)

    # Predictors Correlation Heatmap
    if len(current_predictors) > 0:
        try:
            corr = X[current_predictors].corr()
            print(f"Predictors Correlation Heatmap ({name_severity} data)")

            plt.figure(figsize=(15, len(corr) * 0.9))
            sns.heatmap(corr, annot=True, cmap='coolwarm')
            plt.title(f"Predictors Correlation Heatmap ({name_severity} data)")
            plt.tight_layout()
            plt.show()
        except Exception as e:
            print(f"Could not compute or plot correlation for {name_severity} data: {e}")
    else:
        print(f"No predictors left for correlation analysis for {name_severity} data.")

    high_corr_features = find_high_corr_features(corr, severity=name_severity, threshold=0.3)

    # Correlation against change heatmap
    if len(current_predictors) > 0: # Use current_predictors for the outcome correlation
        try:
            corr_outcome = compute_corr_subset(X, outcome)
            print(f"Correlation with log effect mortality ({name_severity} data):")
            print(corr_outcome)

            plt.figure(figsize=(8, len(corr_outcome) * 0.4))
            sns.heatmap(corr_outcome.to_frame(), annot=True, cmap='coolwarm')
            plt.title(f"Predictors-outcome Correlation Heatmap ({name_severity} data)")
```

```
            plt.tight_layout()
            plt.show()
        except Exception as e:
            print(f"Could not compute or plot predictor-outcome correlation for {name_severity} data: {e}")
    else:
        print(f"No predictors left for predictor-outcome correlation analysis for {name_severity} data.")
```

**4.Modelling (replace with your severity levels, predictors, outcome)**

```
def xgb_objective(trial):
  xgbr = XGBRegressor(random_state=42,early_stopping_rounds=10)
  params = {
      'n_estimators': trial.suggest_int('n_estimators', 50, 300),
      'max_depth': trial.suggest_int('max_depth', 2, 10),
      'learning_rate': trial.suggest_float('learning_rate', 0.01, 0.2),
      'subsample': trial.suggest_float('subsample', 0.4, 0.6),
      'colsample_bytree': trial.suggest_float('colsample_bytree', 0.4, 0.6),
      'min_child_weight': trial.suggest_int('min_child_weight',1, 10),
      'gamma': trial.suggest_float('gamma',0, 0.5)
  }
  cv = KFold(n_splits=5, shuffle=True, random_state=42)
  score = cross_val_score(xgbr, X, y, scoring='neg_mean_absolute_percentage_error', cv=cv)

  random_search = RandomizedSearchCV(
    estimator=xgbr,
    param_distributions=params,
    n_iter=50,
    cv=3,
    verbose=1,
    scoring=score,
    random_state=42
  )
```

```
  random_search.fit(X, y)
  print("Best XGBoost Parameters:", random_search.best_params_)

def find_hyperparameters(X_train, y_train, model_name):
  if model_name == 'Random Forest':
    rfr = RandomForestRegressor(random_state=42)

    rfr_param_dist = {
        'n_estimators': randint(50, 300),
        'max_depth': randint(2, 10),
        'min_samples_split': randint(2, 10),
        'min_samples_leaf': randint(1, 5),
        'max_features': ['sqrt', 'log2', None]
    }

    rfr_search = RandomizedSearchCV(
        estimator=rfr,
        param_distributions=rfr_param_dist,
        n_iter=50,
        cv=3,
        scoring='neg_mean_absolute_percentage_error',
        verbose=1,
        random_state=42,
        n_jobs=-1
    )

    rfr_search.fit(X_train, y_train)
    print("Best RFR Parameters:", rfr_search.best_params_)
    return rfr_search
  elif model_name == 'Gradient Boosting':
    gbr = GradientBoostingRegressor(random_state=42)
    gbr_param_dist = {
```

```
        'n_estimators': randint(50, 300),
        'learning_rate': [0.01, 0.05, 0.1, 0.2],
        'max_depth': randint(2, 10),
        'min_samples_split': randint(2, 10),
        'min_samples_leaf': randint(1, 5),
        'subsample': [0.6, 0.8, 1.0]
    }

    gbr_search = RandomizedSearchCV(
        estimator=gbr,
        param_distributions=gbr_param_dist,
        n_iter=50,
        cv=3,
        scoring='neg_mean_absolute_error',
        verbose=1,
        random_state=42,
        n_jobs=-1
    )

    gbr_search.fit(X_train, y_train)
    print("Best GBR Parameters:", gbr_search.best_params_)
    return gbr_search
elif model_name == 'XGBoost':
    # study = optuna.create_study(direction='maximize')
    # study.optimize(xgb_objective, n_trials=50)
    xgbr = XGBRegressor(random_state=42)
    params = {
        'n_estimators': randint(50, 300),
        'max_depth': randint(2, 10),
        'learning_rate': uniform(0.01, 0.2),
        'subsample': uniform(0.4, 0.6),
        'colsample_bytree': uniform(0.4, 0.6),
        'min_child_weight': randint(1, 10),
```

```
        'gamma': uniform(0, 0.5)
    }
    cv = KFold(n_splits=5, shuffle=True, random_state=42)

    xgbr_search = RandomizedSearchCV(
      estimator=xgbr,
      param_distributions=params,
      n_iter=50,
      cv=cv,
      verbose=1,
      scoring='neg_mean_absolute_percentage_error',
      random_state=42
    )

    xgbr_search.fit(X_train, y_train)
    print("Best XGBoost Parameters:", xgbr_search.best_params_)
    return xgbr_search
  else:
    print("Invalid model name")
    return None

def model_evaluation(models, X_train, X_test, y_train, y_test, name, model_names):
    # Containers for metrics
    precision_list = []
    recall_list = []
    f1_list = []
    auc_list = []
    roc_data = []
    feature_importance_list = []

    # Dynamic threshold based on training and test set
    threshold = ((y_train.min() + y_train.max()) / 2 + (y_test.min() + y_test.max()) / 2) / 2
    y_test_binary = (y_test >= threshold).astype(int)
```

```
for model, model_name in zip(models, model_names):
    model.fit(X_train, y_train)
    y_pred = model.predict(X_test)

    print(f'\nError of {model_name} model for {name} Data')
    MAE = mean_absolute_error(y_test, y_pred)
    MSE = mean_squared_error(y_test, y_pred)
    RMSE = np.sqrt(MSE)
    MAPE = mean_absolute_percentage_error(y_test, y_pred)
    R2 = r2_score(y_test, y_pred)
    print('MAE:', round(MAE, 5))
    print('MSE:', round(MSE, 5))
    print('RMSE:', round(RMSE, 5))
    print('MAPE:', round(MAPE, 5))
    print('R2:', round(R2, 5))

    y_pred_binary = (y_pred >= threshold).astype(int)

    precision = precision_score(y_test_binary, y_pred_binary)
    recall = recall_score(y_test_binary, y_pred_binary)
    f1 = f1_score(y_test_binary, y_pred_binary)
    roc_auc = roc_auc_score(y_test_binary, y_pred)
    fpr, tpr, _ = roc_curve(y_test_binary, y_pred)

    precision_list.append(precision)
    recall_list.append(recall)
    f1_list.append(f1)
    auc_list.append(roc_auc)
    roc_data.append((fpr, tpr, model_name))

    # Feature importances
    feature_importances = model.feature_importances_
```

```
    importance_df = pd.DataFrame(feature_importances, index=X_train.columns, columns=[model_name])
    feature_importance_list.append(importance_df)

# --- Plot Precision, Recall, F1 Score ---
x = np.arange(len(model_names))
width = 0.15

plt.figure(figsize=(10, 5))
plt.bar(x - width, precision_list, width, label='Precision', color='#4CAF50')
plt.bar(x, recall_list, width, label='Recall', color='#2196F3')
plt.bar(x + width, f1_list, width, label='F1 Score', color='#FF5722')

plt.xticks(x, model_names)
plt.ylim(0, 1)
plt.title('Precision, Recall, F1 Score Comparison')
plt.ylabel('Score')
plt.legend()
plt.grid(axis='y', linestyle='--', alpha=0.7)
plt.tight_layout()
plt.show()

# --- Plot ROC Curves ---
plt.figure(figsize=(7, 7))
for fpr, tpr, label in roc_data:
    plt.plot(fpr, tpr, label=f'{label} (AUC = {roc_auc_score(y_test_binary, model.predict(X_test)):.2f})')

plt.plot([0, 1], [0, 1], 'k--', label='Random Classifier')
plt.title('ROC Curve Comparison')
plt.xlabel('False Positive Rate')
plt.ylabel('True Positive Rate')
plt.legend(loc='lower right')
plt.grid(True)
plt.tight_layout()
```

```
    plt.show()

    # --- Feature Importance Heatmap ---
    feature_importance_all = pd.concat(feature_importance_list, axis=1)
    feature_importance_all = feature_importance_all.fillna(0)

    plt.figure(figsize=(10, len(feature_importance_all) * 0.4))
    sns.heatmap(feature_importance_all, annot=True, cmap='coolwarm', cbar=True)
    plt.title(f'Feature Importances Across Models - {name} Data')
    plt.tight_layout()
    plt.show()

    return feature_importance_all
```

Moderate-severe

```
import pandas as pd
import numpy as np

# 1. Filter the modified_data DataFrame to include only rows where the 'Severity' is 'mod-sev'.
modsev_data_filtered = modified_data[modified_data['injury_severity'] == 'moderate-severe'].copy()

# 2. Define the list of predictor columns and the outcome variable.
predictors_modelling = ['age_mean', 'sex_gender_ratio_men_males', 'study_quality', 'gender_ineq']
outcome = 'outcome'

# Combine predictors and outcome for NaN dropping
all_cols = predictors_modelling + [outcome]

# 3. Remove any rows that have missing values in any of the specified predictors or the outcome column.
modsev_data_filtered.dropna(subset=all_cols, inplace=True)

# 4. Separate the cleaned mild_data_filtered into features (X_mild) and the target variable (y_mild).
X_modsev = modsev_data_filtered[predictors_modelling].copy()
```

```
y_modsev = modsev_data_filtered[outcome].copy()

# 6. Convert all columns in X_modsev to a numeric (float) data type, coercing any non-numeric values to NaN.
for col in X_modsev.columns:
    X_modsev[col] = pd.to_numeric(X_modsev[col], errors='coerce')

# 7. Drop any rows from X_mild and y_mild that still contain NaN values after numeric conversion and one-hot encoding,
#    ensuring X_mild and y_mild maintain aligned indices.
# First, align indices before dropping NaNs in X_mild to ensure consistency with y_mild
X_modsev, y_modsev = X_modsev.align(y_modsev, join='inner', axis=0)

initial_len_X = len(X_modsev)
X_modsev.dropna(inplace=True)
y_modsev = y_modsev.loc[X_modsev.index] # Re-align y_mild after dropping rows from X_mild

if len(X_modsev) < initial_len_X:
    print(f"Dropped {initial_len_X - len(X_modsev)} rows due to NaNs after numeric conversion in X_modsev.")

print("Shape of X_modsev:", X_modsev.shape)
print("First 5 rows of X_modsev:\n", X_modsev.head())
print("Shape of y_modsev:", y_modsev.shape)
print("First 5 rows of y_modsev:\n", y_modsev.head())

from sklearn.model_selection import train_test_split from sklearn.ensemble import RandomForestRegressor
```

**1. Split the DataFrames into training and testing sets**

```
X_modsev_train, X_modsev_test, y_modsev_train, y_modsev_test = train_test_split( X_modsev, y_modsev, test_size=0.2,
random_state=52 )
print("Modsev Data - Train/Test Split Shapes:") print(f"X_train: {X_modsev_train.shape}, X_test: {X_modsev_test.shape}")
print(f"y_train: {y_modsev_train.shape}, y_test: {y_modsev_test.shape}")
```

**2. Call the find_hyperparameters function for each model**

```
print("\nFinding hyperparameters for Modsev data (Gradient Boosting)...") gbr_search_modsev =
find_hyperparameters(X_modsev_train, y_modsev_train, model_name='Gradient Boosting')
```

print("\nFinding hyperparameters for Modsev data (Random Forest)...") rfr_search_modsev = find_hyperparameters(X_modsev_train, y_modsev_train, model_name='Random Forest')
print("\nFinding hyperparameters for Modsev data (XGBoost)...") xgbr_search_modsev = find_hyperparameters(X_modsev_train, y_modsev_train, model_name='XGBoost')
**3. Instantiate models with the best hyperparameters**
gbr_modsev = GradientBoostingRegressor(**gbr_search_modsev.best_params_, random_state=42) rfr_modsev = RandomForestRegressor(**rfr_search_modsev.best_params_, random_state=42) xgbr_modsev = XGBRegressor(**xgbr_search_modsev.best_params_, random_state=42)
**4. Create a list named models**
models_modsev = [gbr_modsev, rfr_modsev, xgbr_modsev]
**5. Create a list named model_names**
model_names = ["Gradient Boosting", "Random Forest", "XGBoost"]
**6. Call the model_evaluation function**
model_evaluation( models=models_modsev, X_train=X_modsev_train, X_test=X_modsev_test, y_train=y_modsev_train, y_test=y_modsev_test, name='Moderate-Severe', model_names=model_names )

Severe
from sklearn.model_selection import train_test_split from sklearn.ensemble import RandomForestRegressor from xgboost import XGBRegressor import pandas as pd import numpy as np
**1. Filter the modified_data DataFrame to include only rows where the 'Severity' is 'Severe'.**
sev_data_filtered = modified_data[modified_data['injury_severity'] == 'Severe'].copy()
**Combine predictors and outcome for NaN dropping**
all_cols_sev = predictors_modelling + [outcome]
**2. Remove any rows that have missing values in any of the specified predictors or the outcome column.**
sev_data_filtered.dropna(subset=all_cols_sev, inplace=True)
**3. Separate the cleaned sev_data_filtered into features (X_sev) and the target variable (y_sev).**
X_sev = sev_data_filtered[predictors_modelling].copy() y_sev = sev_data_filtered[outcome].copy()
**4. Convert all columns in X_sev to a numeric (float) data type, coercing any non-numeric values to NaN.**
for col in X_sev.columns: X_sev[col] = pd.to_numeric(X_sev[col], errors='coerce')
**5. Drop any rows from X_sev and y_sev that still contain NaN values after numeric conversion, ensuring X_sev and y_sev maintain aligned indices.**
**First, align indices before dropping NaNs in X_sev to ensure consistency with y_sev**
X_sev, y_sev = X_sev.align(y_sev, join='inner', axis=0)

initial_len_X_sev = len(X_sev) X_sev.dropna(inplace=True) y_sev = y_sev.loc[X_sev.index] # Re-align y_sev after dropping rows from X_sev
if len(X_sev) < initial_len_X_sev: print(f"Dropped {initial_len_X_sev - len(X_sev)} rows due to NaNs after numeric conversion in X_sev.")
print("Shape of X_sev:", X_sev.shape) print("First 5 rows of X_sev:\n", X_sev.head()) print("Shape of y_sev:", y_sev.shape) print("First 5 rows of y_sev:\n", y_sev.head())

**Split the DataFrames into training and testing sets**

X_sev_train, X_sev_test, y_sev_train, y_sev_test = train_test_split( X_sev, y_sev, test_size=0.2, random_state=52 )
print("\nSevere Data - Train/Test Split Shapes:") print(f"X_train: {X_sev_train.shape}, X_test: {X_sev_test.shape}") print(f"y_train: {y_sev_train.shape}, y_test: {y_sev_test.shape}")

**Call the find_hyperparameters function for each model**

print("\nFinding hyperparameters for Severe data (Gradient Boosting)...") gbr_search_sev = find_hyperparameters(X_sev_train, y_sev_train, model_name='Gradient Boosting')
print("\nFinding hyperparameters for Severe data (Random Forest)...") rfr_search_sev = find_hyperparameters(X_sev_train, y_sev_train, model_name='Random Forest')
print("\nFinding hyperparameters for Severe data (XGBoost)...") xgbr_search_sev = find_hyperparameters(X_sev_train, y_sev_train, model_name='XGBoost')

**Instantiate models with the best hyperparameters**

gbr_sev = GradientBoostingRegressor(**gbr_search_sev.best_params_, random_state=42) rfr_sev = RandomForestRegressor(**rfr_search_sev.best_params_, random_state=42) xgbr_sev = XGBRegressor(**xgbr_search_sev.best_params_, random_state=42)

**Create a list named models_sev**

models_sev = [gbr_sev, rfr_sev, xgbr_sev]

**Create a list named model_names**

model_names = ["Gradient Boosting", "Random Forest", "XGBoost"]

**Call the model_evaluation function**

model_evaluation( models=models_sev, X_train=X_sev_train, X_test=X_sev_test, y_train=y_sev_train, y_test=y_sev_test, name='Severe', model_names=model_names )

**SHAP**

Moderate-severe

import shap

```
explainer = shap.TreeExplainer(rfr_modsev)
shap_values = explainer.shap_values(X_modsev_train)

shap.summary_plot(shap_values, X_modsev_train)

explainer = shap.TreeExplainer(gbr_modsev)
shap_values = explainer.shap_values(X_modsev_train)

shap.summary_plot(shap_values, X_modsev_train)

explainer = shap.TreeExplainer(xgbr_modsev)
shap_values = explainer.shap_values(X_modsev_train)

shap.summary_plot(shap_values, X_modsev_train)
```

Severe

```
import shap

explainer = shap.TreeExplainer(rfr_sev)
shap_values = explainer.shap_values(X_sev_train)

shap.summary_plot(shap_values, X_sev_train)

explainer = shap.TreeExplainer(gbr_sev)
shap_values = explainer.shap_values(X_sev_train)

shap.summary_plot(shap_values, X_sev_train)

explainer = shap.TreeExplainer(xgbr_sev)
shap_values = explainer.shap_values(X_sev_train)

shap.summary_plot(shap_values, X_sev_train)
```

**Sensitivity analysis**
Main + age SD
Moderate-severe

```
import pandas as pd
import numpy as np

# 1. Filter the modified_data DataFrame to include only rows where the 'Severity' is 'mod-sev'.
modsev_data_filtered = modified_data[modified_data['injury_severity'] == 'moderate-severe'].copy()

# 2. Define the list of predictor columns and the outcome variable.
predictors_sensone = ['age_mean', 'age_sd', 'sex_gender_ratio_men_males', 'study_quality', 'gender_ineq']
outcome = 'outcome'

# Combine predictors and outcome for NaN dropping
all_cols = predictors_sensone + [outcome]

# 3. Remove any rows that have missing values in any of the specified predictors or the outcome column.
modsev_data_filtered.dropna(subset=all_cols, inplace=True)

# 4. Separate the cleaned modsev_data_filtered into features (X_modsev) and the target variable (y_modsev).
X_modsev = modsev_data_filtered[predictors_sensone].copy()
y_modsev = modsev_data_filtered[outcome].copy()

# 6. Convert all columns in X_modsev to a numeric (float) data type, coercing any non-numeric values to NaN.
for col in X_modsev.columns:
    X_modsev[col] = pd.to_numeric(X_modsev[col], errors='coerce')

# 7. Drop any rows from X_modsev and y_modsev that still contain NaN values after numeric conversion and one-hot encoding,
#    ensuring X_modsev and y_modsev maintain aligned indices.
# First, align indices before dropping NaNs in X_modsev to ensure consistency with y_modsev
X_modsev, y_modsev = X_modsev.align(y_modsev, join='inner', axis=0)
```

```
initial_len_X = len(X_modsev)
X_modsev.dropna(inplace=True)
y_modsev = y_modsev.loc[X_modsev.index] # Re-align y_modsev after dropping rows from X_modsev

if len(X_modsev) < initial_len_X:
    print(f"Dropped {initial_len_X - len(X_modsev)} rows due to NaNs after numeric conversion in X_modsev.")

print("Shape of X_modsev:", X_modsev.shape)
print("First 5 rows of X_modsev:\n", X_modsev.head())
print("Shape of y_modsev:", y_modsev.shape)
print("First 5 rows of y_modsev:\n", y_modsev.head())

from sklearn.model_selection import train_test_split from sklearn.ensemble import RandomForestRegressor
```

**1. Split the DataFrames into training and testing sets**

```
X_modsev_train, X_modsev_test, y_modsev_train, y_modsev_test = train_test_split( X_modsev, y_modsev, test_size=0.2,
random_state=52 )
print("Modsev Data - Train/Test Split Shapes:") print(f"X_train: {X_modsev_train.shape}, X_test: {X_modsev_test.shape}")
print(f"y_train: {y_modsev_train.shape}, y_test: {y_modsev_test.shape}")
```

**2. Call the find_hyperparameters function for each model**

```
print("\nFinding hyperparameters for Modsev data (Gradient Boosting)...") gbr_search_modsev =
find_hyperparameters(X_modsev_train, y_modsev_train, model_name='Gradient Boosting')
print("\nFinding hyperparameters for Modsev data (Random Forest)...") rfr_search_modsev = find_hyperparameters(X_modsev_train,
y_modsev_train, model_name='Random Forest')
print("\nFinding hyperparameters for Modsev data (XGBoost)...") xgbr_search_modsev = find_hyperparameters(X_modsev_train,
y_modsev_train, model_name='XGBoost')
```

**3. Instantiate models with the best hyperparameters**

```
gbr_modsev = GradientBoostingRegressor(**gbr_search_modsev.best_params_, random_state=42) rfr_modsev =
RandomForestRegressor(**rfr_search_modsev.best_params_, random_state=42) xgbr_modsev =
XGBRegressor(**xgbr_search_modsev.best_params_, random_state=42)
```

**4. Create a list named models**

```
models_modsev = [gbr_modsev, rfr_modsev, xgbr_modsev]
```

**5. Create a list named model_names**
model_names = ["Gradient Boosting", "Random Forest", "XGBoost"]
**6. Call the model_evaluation function**
model_evaluation( models=models_modsev, X_train=X_modsev_train, X_test=X_modsev_test, y_train=y_modsev_train, y_test=y_modsev_test, name='Moderate-Severe', model_names=model_names )
import shap

explainer = shap.TreeExplainer(rfr_modsev)
shap_values = explainer.shap_values(X_modsev_train)

shap.summary_plot(shap_values, X_modsev_train)

import shap

explainer = shap.TreeExplainer(gbr_modsev)
shap_values = explainer.shap_values(X_modsev_train)

shap.summary_plot(shap_values, X_modsev_train)

import shap

explainer = shap.TreeExplainer(xgbr_modsev)
shap_values = explainer.shap_values(X_modsev_train)

shap.summary_plot(shap_values, X_modsev_train)

Severe
from sklearn.model_selection import train_test_split from sklearn.ensemble import RandomForestRegressor from xgboost import XGBRegressor import pandas as pd import numpy as np
**1. Filter the modified_data DataFrame to include only rows where the 'Severity' is 'Severe'.**
sev_data_filtered = modified_data[modified_data['injury_severity'] == 'Severe'].copy()

**Combine predictors and outcome for NaN dropping**
all_cols_sev = predictors_sensone + [outcome]
**2. Remove any rows that have missing values in any of the specified predictors or the outcome column.**
sev_data_filtered.dropna(subset=all_cols_sev, inplace=True)
**3. Separate the cleaned sev_data_filtered into features (X_sev) and the target variable (y_sev).**
X_sev = sev_data_filtered[predictors_sensone].copy() y_sev = sev_data_filtered[outcome].copy()
**4. Convert all columns in X_sev to a numeric (float) data type, coercing any non-numeric values to NaN.**
for col in X_sev.columns: X_sev[col] = pd.to_numeric(X_sev[col], errors='coerce')
**5. Drop any rows from X_sev and y_sev that still contain NaN values after numeric conversion, ensuring X_sev and y_sev maintain aligned indices.**
**First, align indices before dropping NaNs in X_sev to ensure consistency with y_sev**
X_sev, y_sev = X_sev.align(y_sev, join='inner', axis=0)
initial_len_X_sev = len(X_sev) X_sev.dropna(inplace=True) y_sev = y_sev.loc[X_sev.index] # Re-align y_sev after dropping rows from X_sev
if len(X_sev) < initial_len_X_sev: print(f"Dropped {initial_len_X_sev - len(X_sev)} rows due to NaNs after numeric conversion in X_sev.")
print("Shape of X_sev:", X_sev.shape) print("First 5 rows of X_sev:\n", X_sev.head()) print("Shape of y_sev:", y_sev.shape) print("First 5 rows of y_sev:\n", y_sev.head())
**Split the DataFrames into training and testing sets**
X_sev_train, X_sev_test, y_sev_train, y_sev_test = train_test_split( X_sev, y_sev, test_size=0.2, random_state=52 )
print("\nSevere Data - Train/Test Split Shapes:") print(f"X_train: {X_sev_train.shape}, X_test: {X_sev_test.shape}") print(f"y_train: {y_sev_train.shape}, y_test: {y_sev_test.shape}")
**Call the find_hyperparameters function for each model**
print("\nFinding hyperparameters for Severe data (Gradient Boosting)...") gbr_search_sev = find_hyperparameters(X_sev_train, y_sev_train, model_name='Gradient Boosting')
print("\nFinding hyperparameters for Severe data (Random Forest)...") rfr_search_sev = find_hyperparameters(X_sev_train, y_sev_train, model_name='Random Forest')
print("\nFinding hyperparameters for Severe data (XGBoost)...") xgbr_search_sev = find_hyperparameters(X_sev_train, y_sev_train, model_name='XGBoost')
**Instantiate models with the best hyperparameters**
gbr_sev = GradientBoostingRegressor(**gbr_search_sev.best_params_, random_state=42) rfr_sev = RandomForestRegressor(**rfr_search_sev.best_params_, random_state=42) xgbr_sev = XGBRegressor(**xgbr_search_sev.best_params_, random_state=42)

**Create a list named models_sev**

```
models_sev = [gbr_sev, rfr_sev, xgbr_sev]
```

**Create a list named model_names**

```
model_names = ["Gradient Boosting", "Random Forest", "XGBoost"]
```

**Call the model_evaluation function**

```
model_evaluation( models=models_sev, X_train=X_sev_train, X_test=X_sev_test, y_train=y_sev_train, y_test=y_sev_test, name='Severe', model_names=model_names )
import shap

explainer = shap.TreeExplainer(rfr_sev)
shap_values = explainer.shap_values(X_sev_train)

shap.summary_plot(shap_values, X_sev_train)

import shap

explainer = shap.TreeExplainer(gbr_sev)
shap_values = explainer.shap_values(X_sev_train)

shap.summary_plot(shap_values, X_sev_train)

import shap

explainer = shap.TreeExplainer(xgbr_sev)
shap_values = explainer.shap_values(X_sev_train)

shap.summary_plot(shap_values, X_sev_train)
```

Main + region

Moderate-severe

```
region = ['North America', 'Europe', 'Asia/Pacific']
```

```
import pandas as pd
import numpy as np

# 1. Filter the modified_data DataFrame to include only rows where the 'Severity' is 'mod-sev'.
modsev_data_filtered = modified_data[modified_data['injury_severity'] == 'moderate-severe'].copy()

# 2. Define the list of predictor columns and the outcome variable.
predictors_senstwo = ['age_mean', 'sex_gender_ratio_men_males', 'study_quality', 'gender_ineq'] + region
outcome = 'outcome'

# Combine predictors and outcome for NaN dropping
all_cols = predictors_senstwo + [outcome]

# 3. Remove any rows that have missing values in any of the specified predictors or the outcome column.
modsev_data_filtered.dropna(subset=all_cols, inplace=True)

# 4. Separate the cleaned modsev_data_filtered into features (X_modsev) and the target variable (y_modsev).
X_modsev = modsev_data_filtered[predictors_senstwo].copy()
y_modsev = modsev_data_filtered[outcome].copy()

# 6. Convert all columns in X_modsev to a numeric (float) data type, coercing any non-numeric values to NaN.
for col in X_modsev.columns:
    X_modsev[col] = pd.to_numeric(X_modsev[col], errors='coerce')

# 7. Drop any rows from X_modsev and y_modsev that still contain NaN values after numeric conversion and one-hot encoding,
#    ensuring X_modsev and y_modsev maintain aligned indices.
# First, align indices before dropping NaNs in X_modsev to ensure consistency with y_modsev
X_modsev, y_modsev = X_modsev.align(y_modsev, join='inner', axis=0)

initial_len_X = len(X_modsev)
X_modsev.dropna(inplace=True)
y_modsev = y_modsev.loc[X_modsev.index] # Re-align y_modsev after dropping rows from X_modsev
```

```
if len(X_modsev) < initial_len_X:
    print(f"Dropped {initial_len_X - len(X_modsev)} rows due to NaNs after numeric conversion in X_modsev.")

print("Shape of X_modsev:", X_modsev.shape)
print("First 5 rows of X_modsev:\n", X_modsev.head())
print("Shape of y_modsev:", y_modsev.shape)
print("First 5 rows of y_modsev:\n", y_modsev.head())

from sklearn.model_selection import train_test_split from sklearn.ensemble import RandomForestRegressor
```

**1. Split the DataFrames into training and testing sets**

X_modsev_train, X_modsev_test, y_modsev_train, y_modsev_test = train_test_split( X_modsev, y_modsev, test_size=0.2, random_state=52 )
print("Modsev Data - Train/Test Split Shapes:") print(f"X_train: {X_modsev_train.shape}, X_test: {X_modsev_test.shape}")
print(f"y_train: {y_modsev_train.shape}, y_test: {y_modsev_test.shape}")

**2. Call the find_hyperparameters function for each model**

print("\nFinding hyperparameters for Modsev data (Gradient Boosting)...") gbr_search_modsev = find_hyperparameters(X_modsev_train, y_modsev_train, model_name='Gradient Boosting')
print("\nFinding hyperparameters for Modsev data (Random Forest)...") rfr_search_modsev = find_hyperparameters(X_modsev_train, y_modsev_train, model_name='Random Forest')
print("\nFinding hyperparameters for Modsev data (XGBoost)...") xgbr_search_modsev = find_hyperparameters(X_modsev_train, y_modsev_train, model_name='XGBoost')

**3. Instantiate models with the best hyperparameters**

gbr_modsev = GradientBoostingRegressor(**gbr_search_modsev.best_params_, random_state=42) rfr_modsev = RandomForestRegressor(**rfr_search_modsev.best_params_, random_state=42) xgbr_modsev = XGBRegressor(**xgbr_search_modsev.best_params_, random_state=42)

**4. Create a list named models**

models_modsev = [gbr_modsev, rfr_modsev, xgbr_modsev]

**5. Create a list named model_names**

model_names = ["Gradient Boosting", "Random Forest", "XGBoost"]

**6. Call the model_evaluation function**

model_evaluation( models=models_modsev, X_train=X_modsev_train, X_test=X_modsev_test, y_train=y_modsev_train, y_test=y_modsev_test, name='Moderate-Severe with region', model_names=model_names )
import shap

```
explainer = shap.TreeExplainer(rfr_modsev)
shap_values = explainer.shap_values(X_modsev_train)

shap.summary_plot(shap_values, X_modsev_train)

import shap

explainer = shap.TreeExplainer(gbr_modsev)
shap_values = explainer.shap_values(X_modsev_train)

shap.summary_plot(shap_values, X_modsev_train)

import shap

explainer = shap.TreeExplainer(xgbr_modsev)
shap_values = explainer.shap_values(X_modsev_train)

shap.summary_plot(shap_values, X_modsev_train)
```

Severe

from sklearn.model_selection import train_test_split from sklearn.ensemble import RandomForestRegressor from xgboost import XGBRegressor import pandas as pd import numpy as np

**1. Filter the modified_data DataFrame to include only rows where the 'Severity' is 'Severe'.**

sev_data_filtered = modified_data[modified_data['injury_severity'] == 'Severe'].copy()

**Combine predictors and outcome for NaN dropping**

all_cols_sev = predictors_senstwo + [outcome]

**2. Remove any rows that have missing values in any of the specified predictors or the outcome column.**

sev_data_filtered.dropna(subset=all_cols_sev, inplace=True)

**3. Separate the cleaned sev_data_filtered into features (X_sev) and the target variable (y_sev).**

X_sev = sev_data_filtered[predictors_senstwo].copy() y_sev = sev_data_filtered[outcome].copy()

**4. Convert all columns in X_sev to a numeric (float) data type, coercing any non-numeric values to NaN.**

for col in X_sev.columns: X_sev[col] = pd.to_numeric(X_sev[col], errors='coerce')

**5. Drop any rows from X_sev and y_sev that still contain NaN values after numeric conversion, ensuring X_sev and y_sev maintain aligned indices.**

**First, align indices before dropping NaNs in X_sev to ensure consistency with y_sev**

X_sev, y_sev = X_sev.align(y_sev, join='inner', axis=0)

initial_len_X_sev = len(X_sev) X_sev.dropna(inplace=True) y_sev = y_sev.loc[X_sev.index] # Re-align y_sev after dropping rows from X_sev

if len(X_sev) < initial_len_X_sev: print(f"Dropped {initial_len_X_sev - len(X_sev)} rows due to NaNs after numeric conversion in X_sev.")

print("Shape of X_sev:", X_sev.shape) print("First 5 rows of X_sev:\n", X_sev.head()) print("Shape of y_sev:", y_sev.shape) print("First 5 rows of y_sev:\n", y_sev.head())

**Split the DataFrames into training and testing sets**

X_sev_train, X_sev_test, y_sev_train, y_sev_test = train_test_split( X_sev, y_sev, test_size=0.2, random_state=52 )

print("\nSevere Data - Train/Test Split Shapes:") print(f"X_train: {X_sev_train.shape}, X_test: {X_sev_test.shape}") print(f"y_train: {y_sev_train.shape}, y_test: {y_sev_test.shape}")

**Call the find_hyperparameters function for each model**

print("\nFinding hyperparameters for Severe data (Gradient Boosting)...") gbr_search_sev = find_hyperparameters(X_sev_train, y_sev_train, model_name='Gradient Boosting')

print("\nFinding hyperparameters for Severe data (Random Forest)...") rfr_search_sev = find_hyperparameters(X_sev_train, y_sev_train, model_name='Random Forest')

print("\nFinding hyperparameters for Severe data (XGBoost)...") xgbr_search_sev = find_hyperparameters(X_sev_train, y_sev_train, model_name='XGBoost')

**Instantiate models with the best hyperparameters**

gbr_sev = GradientBoostingRegressor(**gbr_search_sev.best_params_, random_state=42) rfr_sev = RandomForestRegressor(**rfr_search_sev.best_params_, random_state=42) xgbr_sev = XGBRegressor(**xgbr_search_sev.best_params_, random_state=42)

**Create a list named models_sev**

models_sev = [gbr_sev, rfr_sev, xgbr_sev]

**Create a list named model_names**

model_names = ["Gradient Boosting", "Random Forest", "XGBoost"]

**Call the model_evaluation function**

```
model_evaluation( models=models_sev, X_train=X_sev_train, X_test=X_sev_test, y_train=y_sev_train, y_test=y_sev_test, name='Severe', model_names=model_names )
import shap

explainer = shap.TreeExplainer(rfr_sev)
shap_values = explainer.shap_values(X_sev_train)

shap.summary_plot(shap_values, X_sev_train)

explainer = shap.TreeExplainer(gbr_sev)
shap_values = explainer.shap_values(X_sev_train)

shap.summary_plot(shap_values, X_sev_train)

explainer = shap.TreeExplainer(xgbr_sev)
shap_values = explainer.shap_values(X_sev_train)

shap.summary_plot(shap_values, X_sev_train)
```

Main + n

Moderate-severe

```
import pandas as pd
import numpy as np

# 1. Filter the modified_data DataFrame to include only rows where the 'Severity' is 'mod-sev'.
modsev_data_filtered = modified_data[modified_data['injury_severity'] == 'moderate-severe'].copy()

# 2. Define the list of predictor columns and the outcome variable.
predictors_sensthree = ['age_mean', 'cohort_size', 'sex_gender_ratio_men_males', 'study_quality', 'gender_ineq']
outcome = 'outcome'
```

```
# Combine predictors and outcome for NaN dropping
all_cols = predictors_sensthree + [outcome]

# 3. Remove any rows that have missing values in any of the specified predictors or the outcome column.
modsev_data_filtered.dropna(subset=all_cols, inplace=True)

# 4. Separate the cleaned modsev_data_filtered into features (X_modsev) and the target variable (y_modsev).
X_modsev = modsev_data_filtered[predictors_sensthree].copy()
y_modsev = modsev_data_filtered[outcome].copy()

# 6. Convert all columns in X_modsev to a numeric (float) data type, coercing any non-numeric values to NaN.
for col in X_modsev.columns:
    X_modsev[col] = pd.to_numeric(X_modsev[col], errors='coerce')

# 7. Drop any rows from X_modsev and y_modsev that still contain NaN values after numeric conversion and one-hot encoding,
#    ensuring X_modsev and y_modsev maintain aligned indices.
# First, align indices before dropping NaNs in X_modsev to ensure consistency with y_modsev
X_modsev, y_modsev = X_modsev.align(y_modsev, join='inner', axis=0)

initial_len_X = len(X_modsev)
X_modsev.dropna(inplace=True)
y_modsev = y_modsev.loc[X_modsev.index] # Re-align y_modsev after dropping rows from X_modsev

if len(X_modsev) < initial_len_X:
    print(f"Dropped {initial_len_X - len(X_modsev)} rows due to NaNs after numeric conversion in X_modsev.")

print("Shape of X_modsev:", X_modsev.shape)
print("First 5 rows of X_modsev:\n", X_modsev.head())
print("Shape of y_modsev:", y_modsev.shape)
print("First 5 rows of y_modsev:\n", y_modsev.head())

from sklearn.model_selection import train_test_split from sklearn.ensemble import RandomForestRegressor
```

**1. Split the DataFrames into training and testing sets**

X_modsev_train, X_modsev_test, y_modsev_train, y_modsev_test = train_test_split( X_modsev, y_modsev, test_size=0.2, random_state=52 )
print("Modsev Data - Train/Test Split Shapes:") print(f"X_train: {X_modsev_train.shape}, X_test: {X_modsev_test.shape}")
print(f"y_train: {y_modsev_train.shape}, y_test: {y_modsev_test.shape}")

**2. Call the find_hyperparameters function for each model**

print("\nFinding hyperparameters for Modsev data (Gradient Boosting)...") gbr_search_modsev = find_hyperparameters(X_modsev_train, y_modsev_train, model_name='Gradient Boosting')
print("\nFinding hyperparameters for Modsev data (Random Forest)...") rfr_search_modsev = find_hyperparameters(X_modsev_train, y_modsev_train, model_name='Random Forest')
print("\nFinding hyperparameters for Modsev data (XGBoost)...") xgbr_search_modsev = find_hyperparameters(X_modsev_train, y_modsev_train, model_name='XGBoost')

**3. Instantiate models with the best hyperparameters**

gbr_modsev = GradientBoostingRegressor(**gbr_search_modsev.best_params_, random_state=42) rfr_modsev = RandomForestRegressor(**rfr_search_modsev.best_params_, random_state=42) xgbr_modsev = XGBRegressor(**xgbr_search_modsev.best_params_, random_state=42)

**4. Create a list named models**

models_modsev = [gbr_modsev, rfr_modsev, xgbr_modsev]

**5. Create a list named model_names**

model_names = ["Gradient Boosting", "Random Forest", "XGBoost"]

**6. Call the model_evaluation function**

model_evaluation( models=models_modsev, X_train=X_modsev_train, X_test=X_modsev_test, y_train=y_modsev_train, y_test=y_modsev_test, name='Moderate-Severe with cohort size', model_names=model_names )
explainer = shap.TreeExplainer(rfr_modsev)
shap_values = explainer.shap_values(X_modsev_train)

shap.summary_plot(shap_values, X_modsev_train)

explainer = shap.TreeExplainer(gbr_modsev)
shap_values = explainer.shap_values(X_modsev_train)

shap.summary_plot(shap_values, X_modsev_train)

explainer = shap.TreeExplainer(xgbr_modsev)
shap_values = explainer.shap_values(X_modsev_train)

shap.summary_plot(shap_values, X_modsev_train)

Severe
from sklearn.model_selection import train_test_split from sklearn.ensemble import RandomForestRegressor from xgboost import XGBRegressor import pandas as pd import numpy as np
**1. Filter the modified_data DataFrame to include only rows where the 'Severity' is 'Severe'.**
sev_data_filtered = modified_data[modified_data['injury_severity'] == 'Severe'].copy()
**Combine predictors and outcome for NaN dropping**
all_cols_sev = predictors_sensthree + [outcome]
**2. Remove any rows that have missing values in any of the specified predictors or the outcome column.**
sev_data_filtered.dropna(subset=all_cols_sev, inplace=True)
**3. Separate the cleaned sev_data_filtered into features (X_sev) and the target variable (y_sev).**
X_sev = sev_data_filtered[predictors_sensthree].copy() y_sev = sev_data_filtered[outcome].copy()
**4. Convert all columns in X_sev to a numeric (float) data type, coercing any non-numeric values to NaN.**
for col in X_sev.columns: X_sev[col] = pd.to_numeric(X_sev[col], errors='coerce')
**5. Drop any rows from X_sev and y_sev that still contain NaN values after numeric conversion, ensuring X_sev and y_sev maintain aligned indices.**
**First, align indices before dropping NaNs in X_sev to ensure consistency with y_sev**
X_sev, y_sev = X_sev.align(y_sev, join='inner', axis=0)
initial_len_X_sev = len(X_sev) X_sev.dropna(inplace=True) y_sev = y_sev.loc[X_sev.index] # Re-align y_sev after dropping rows from X_sev
if len(X_sev) < initial_len_X_sev: print(f"Dropped {initial_len_X_sev - len(X_sev)} rows due to NaNs after numeric conversion in X_sev.")
print("Shape of X_sev:", X_sev.shape) print("First 5 rows of X_sev:\n", X_sev.head()) print("Shape of y_sev:", y_sev.shape) print("First 5 rows of y_sev:\n", y_sev.head())
**Split the DataFrames into training and testing sets**
X_sev_train, X_sev_test, y_sev_train, y_sev_test = train_test_split( X_sev, y_sev, test_size=0.2, random_state=52 )
print("\nSevere Data - Train/Test Split Shapes:") print(f"X_train: {X_sev_train.shape}, X_test: {X_sev_test.shape}") print(f"y_train: {y_sev_train.shape}, y_test: {y_sev_test.shape}")

**Call the find_hyperparameters function for each model**
print("\nFinding hyperparameters for Severe data (Gradient Boosting)...") gbr_search_sev = find_hyperparameters(X_sev_train, y_sev_train, model_name='Gradient Boosting')
print("\nFinding hyperparameters for Severe data (Random Forest)...") rfr_search_sev = find_hyperparameters(X_sev_train, y_sev_train, model_name='Random Forest')
print("\nFinding hyperparameters for Severe data (XGBoost)...") xgbr_search_sev = find_hyperparameters(X_sev_train, y_sev_train, model_name='XGBoost')
**Instantiate models with the best hyperparameters**
gbr_sev = GradientBoostingRegressor(**gbr_search_sev.best_params_, random_state=42) rfr_sev = RandomForestRegressor(**rfr_search_sev.best_params_, random_state=42) xgbr_sev = XGBRegressor(**xgbr_search_sev.best_params_, random_state=42)
**Create a list named models_sev**
models_sev = [gbr_sev, rfr_sev, xgbr_sev]
**Create a list named model_names**
model_names = ["Gradient Boosting", "Random Forest", "XGBoost"]
**Call the model_evaluation function**
model_evaluation( models=models_sev, X_train=X_sev_train, X_test=X_sev_test, y_train=y_sev_train, y_test=y_sev_test, name='Severe', model_names=model_names )

```
explainer = shap.TreeExplainer(rfr_sev)
shap_values = explainer.shap_values(X_sev_train)

shap.summary_plot(shap_values, X_sev_train)

explainer = shap.TreeExplainer(gbr_sev)
shap_values = explainer.shap_values(X_sev_train)

shap.summary_plot(shap_values, X_sev_train)

explainer = shap.TreeExplainer(xgbr_sev)
shap_values = explainer.shap_values(X_sev_train)

shap.summary_plot(shap_values, X_sev_train)
```
